## Supplementary Material for "Informing pandemic response in the face of uncertainty. *An evaluation of the U.S. COVID-19 Scenario Modeling Hub*"

Emily Howerton (The Pennsylvania State University (PSU)), Lucie Contamin (University of Pittsburgh), Luke C Mullany (Johns Hopkins University Applied Physics Lab (JHU/APL)), Michelle Qin (Harvard University), Nicholas G. Reich (University of Massachusetts Amherst), Samantha Bents (National Institutes of Health Fogarty International Center (NIH)), Rebecca K. Borchering (PSU, Centers for Disease Control and Prevention (CDC)), Sung-mok Jung (University of North Carolina (UNC)) Sara L. Loo (Johns Hopkins University Infectious Disease Dynamics (JHU-IDD)) Claire P. Smith (JHU-IDD), John Levander (University of Pittsburgh (UPitt)), Jessica Kerr (UPitt), J. Espino (UPitt), Willem G. van Panhuis (National Institute of Allergy and Infectious Diseases), Harry Hochheiser (UPitt), Marta Galanti (Columbia University (CU)), Teresa Yamana (CU), Sen Pei (CU), Jeffrey Shaman (CU), Kaitlin Rainwater-Lovett (JHU/APL), Matt Kinsey (JHU/APL), Kate Tallaksen (JHU/APL), Shelby Wilson (JHU/APL), Lauren Shin (JHU/APL), Joseph C. Lemaitre (UNC), Joshua Kaminsky (JHU-IDD), Juan Dent Hulse (JHU-IDD), Elizabeth C. Lee (JHU-IDD), Clif McKee (JHU-IDD), Alison Hill (JHU-IDD), Dean Karlen (University of Victoria (UVic)), Matteo Chinazzi (Northeastern University (NEU)), Jessica T. Davis (NEU), Kunpeng Mu (NEU), Xinyue Xiong (NEU), Ana Pastore y Piontti (NEU), Alessandro Vespignani (NEU), Erik T. Rosenstrom (North Carolina State University (NCSU)), Julie S. Ivy (NCSU), Maria E. Mayorga (NCSU), Julie L. Swann (NCSU), Guido España (University of Notre Dame (UND)), Sean Cavany (UND), Sean Moore (UND), Alex Perkins (UND), Thomas Hladish (University of Florida (UF)), Alexander Pillai (UF), Kok Ben Toh (Northwestern University), Ira Longini Jr. (UF), Shi Chen (University of North Carolina at Charlotte (UNCC)), Rajib Paul (UNCC), Daniel Janies (UNCC), Jean-Claude Thill (UNCC), Anass Bouchnita (University of Texas at El Paso (UTEP)), Kaiming Bi (University of Texas at Austin (UTA)), Michael Lachmann (Santa Fe Institute (SFI)), Spencer Fox (University of Georgia (UGA)), Lauren Ancel Meyers (UTA), UT COVID-19 Modeling Consortium, Ajitesh Srivastava (University of Southern California), Przemyslaw Porebski (University of Virginia (UVA)), Srini Venkatramanan (UVA), Aniruddha Adiga (UVA), Bryan Lewis (UVA), Brian Klahn (UVA), Joseph Outten (UVA), Benjamin Hurt (UVA), Jiangzhuo Chen (UVA), Henning Mortveit (UVA), Amanda Wilson (UVA), Madhav Marathe (UVA), Stefan Hoops (UVA), Parantapa Bhattacharya (UVA), Dustin Machi (UVA), Betsy L. Cadwell (CDC), Jessica M. Healy (CDC), Rachel B. Slayton (CDC), Michael A. Johansson (CDC), Matthew Biggerstaff (CDC), Shaun Truelove (JHU-IDD), Michael C. Runge (U.S. Geological Survey), Katriona Shea (PSU), Cécile Viboud* (NIH), Justin Lessler* (UNC)
*co-senior authors

### Table of Contents

S1. Projections evaluated
Figure S1
Table S1-Table S2
S2. Evaluating scenario plausibility
Figure S2-Figure S6
Table S3
S3. Comparative models
Figure S7-Figure S22
S4. Trend classification
Figure S23-Figure S44
S5. Additional performance results
Figure S45-Figure S60
S6. References

### S1. Projections evaluated

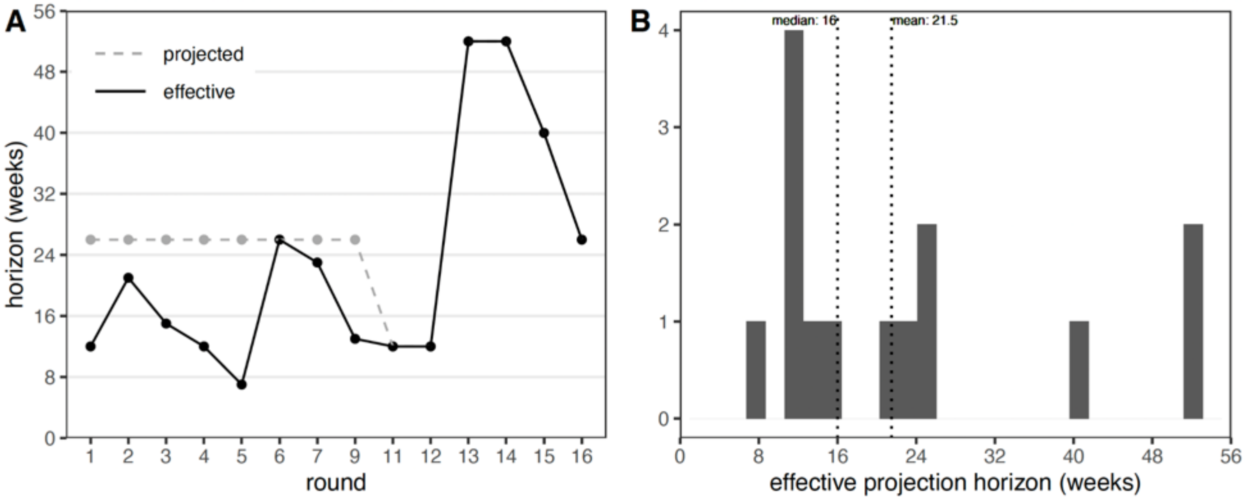


**Figure S1: Projection horizon versus effective horizon.** Effective projection horizon is defined as the number of weeks projected before the emergence of an unanticipated SARS-CoV-2 variant. (A) Effective projection horizon for each round (black line) compared to horizon projected (gray dashed line). (B) Histogram of effective projection horizon across 14 public rounds, including median and mean (dotted vertical lines).

**Table S1:** Summary of individual models that submitted projections in the first sixteen rounds of the U.S. COVID-19 Scenario Modeling Hub (SMH).

| **CU-AGE-ST**<br>Columbia University |
|---|
| The CU-AGE-ST uses a combination of two models for producing age-stratified, state level projections of SARS-COV-2 in the United States: a metapopulation, non-age-stratified model for a single strain and a state level compartmental model, population and age-stratified. Specifically, the inference step is performed with a metapopulation model that reproduces transmission within and between the 3,142 counties in the United States and assimilates daily county cases to estimate the county-level distribution of parameters and variables. Inter-county mobility is modeled using commuting patterns from U.S. Census Bureau, adjusted with Safegraph mobility |

observations from 2020. The inferred parameters and initial conditions are aggregated at the state level and combined with state-level data on population structure, age and state-specific seroprevalence estimates and published estimates of age-specific reporting rates, hospitalization rates, and death rates. The Centers for Disease Control and Prevention's (CDC's) Nationwide Commercial Laboratory Seroprevalence Survey estimates are used to assess the relative difference in age-specific susceptibility and age-specific incidence. Projections are produced with a separate compartmental model run in isolation for each of the 50 U.S. states and the District of Columbia (DC). This second model is stratified in 12 age and population-specific groups to reproduce different patterns of disease severity, different impact of non-pharmaceutical interventions (NPIs), and vaccine prioritization. The model imposes a seasonal forcing on the real time reproductive number.

**JHUAPL-Bucky**
Johns Hopkins Applied Physics Laboratory

The JHUAPL-Bucky model is an age-stratified, county-level susceptible-exposed-infected-recovered (SEIR) model. The Bucky model estimates cases, hospitalizations, and deaths due to COVID-19 across all counties and territories in the United States.

Geographic spatial considerations are incorporated via public mobility data which accounts for inter-county mobility. Contact matrices derived from Prem *et al.* (*1*) are used to account for interactions between age groups within a given county/territory. In addition to static data sources related to the demographic distribution of individuals, dynamic input to the model consists primarily of incident case, hospitalization and death data from the two weeks preceding a simulation. These input data are smoothed/approximated with a generalized additive model.

Both static and dynamic input data sources are used to direct model parameter estimation methods. Parameters are selected via a joint optimization of all the priors via a Bayesian optimization procedure to maximize the coverage of our confidence intervals on the historical input data. Parameters (case reporting rates, doubling times, initial conditions, etc.) are estimated locally at the county or state level in order to account for differences in disease and transmission properties due to variation in population demographics and variant prevalence.

To characterize uncertainty associated with the output of a given simulation, two-thousand Monte Carlo runs are performed over multiplicative distributions of the individual parameter estimates. Predictive quantiles are computed from the outputs of these simulations.

**JHU_IDD-CovidSP**
Johns Hopkins University

The Flexible Epidemic Modeling Pipeline (flepiMoP, formerly covidSP) models transmission of SARS-CoV-2 in the United States at the state level using a compartmental metapopulation structure, where U.S. states are connected through human mobility informed by commuting census data. The model compartments have varied over the course of the COVID-19 pandemic and between Scenario Modeling Hub rounds, depending on round specifications. In round 12, our model represented the population though 300 compartments per state, built from all the possible combinations of five disease stages (Susceptible, Exposed, Infectious, Recovered, and recovered with Waned immunity; SEIRW), five vaccination statuses (unvaccinated, vaccinated with 1 dose, vaccinated with 2 doses or boosted, vaccinated with waned vaccine-derived immunity, and unvaccinated with prior infection), four variants (Wild, Alpha, Delta, Omicron), and three age-strata (below 18, 18 to 64, above 65).
Transmission is simulated from January 1st, 2020, and the early infections of each variant are seeded into the different states according to their first observations. The transitions between the compartments are simulated by integrating the governing coupled ordinary differential equations. Transition rates are modified by state- and time-specific non-pharmaceutical interventions, state-specific seasonality of SARS-CoV-2 transmission, and local variation in overall transmissibility by state. The incidence of infection from the dynamical model is passed to an observation model that computes observed cases, hospitalizations, and deaths, where the case detection probability varies in time and space, and the infection fatality ratio (IFR) varies by state, age group, vaccination status, and variant.
While many parameters are derived from the literature (e.g., vaccine-induced protection against infection or death), other parameters are fitted (infection to case ratio, impact of non-pharmaceutical interventions, seasonality, and local transmissibility). We calibrate the model to CSSE data for death and cases in each state along with the variant proportions from CoVariants.org (GISAID) through our custom inference algorithm. This algorithm enables Bayesian inference on large-scale dynamic models through multi-chain Markov Chain Monte Carlo (MCMC), leveraging parallel computing resources with many short chains to handle large-scale epidemic dynamics and

high-dimensional parameter space.

**Karlen-pypm**
University of Victoria

Models built with the python population modeling framework (www.pypm.ca) use finite time difference equations to model a homogeneous population that reproduces the cases, hospitalizations, and deaths time-series data for the jurisdictions under study. The infection rate is proportional to the product of transmission rate, susceptible fraction, and number of circulating contagious individuals. The transmission rate is piecewise constant, adjusted to match case rates, typically constant over a period of 2 months. Adjustable time delays for partial propagations from infected to symptomatic to reported cases to hospitalization and to death are fit to match data. Reporting fraction is adjusted to match seroprevalence data. Multiple infection cycles are included to model variants, using a common susceptible population. An additional susceptible population is available to variants that partially escape from immunity in the population. Selection coefficients estimated from genomic data are used to set transmission rates for new variants as they emerge. Vaccination implemented with a fraction of those vaccinated reducing the susceptible population. Waning of vaccine and natural immunity are included with adjustable time delays to repopulate the susceptible population. The models can produce time series of expectation values and time series of simulated stochastic data. The former is used to produce point estimates for future rates and the latter to produce intervals.

**MOBS_NEU-GLEAM_COVID**
Northeastern University

The model is a U.S. specific metapopulation compartmental model that refines our Global and Epidemic modeling approach. The GLEAM model is a stochastic, spatial, age-structured metapopulation model. Previously this model was used to characterize the early stage of the COVID-19 epidemic (*2*). Each subpopulation is defined as the catchment area around major transportation hubs. The airline transportation data encompass daily travel data in the origin–destination format from the Official Aviation Guide database50 reflecting actual traffic changes that occurred during the pandemic. Ground mobility and commuting flows are derived from the analysis and modeling of data collected from the statistics offices of 30 countries on 5 continents. The model accounts for travel restrictions and government-issued policies. Furthermore, the model accounts for the reduction of internal, country-wide mobility and changes in contact patterns in each country and state. The U.S. local epidemic and mobility model (LEAM) is enhanced to consider as a single subpopulation each one of the 3,142 counties (or its statistical equivalent) for each one of the 50 U.S. states. Population size and county-specific age distributions reflect Census' annual resident population estimates for year 2019. Commuting flows between counties are obtained from the 2011-2015 5-Year ACS Commuting Flows survey and properly adjusted to account for differences in population totals since the creation of the dataset. Contact matrices, age-specific traveling probabilities, and air traffic flows are in common with GLEAM and they are properly mapped at LEAM-US county-level resolution. Google's COVID-19 Community Mobility Reports data collected at the county-level resolution are used to model mobility and the effects of NPIs on individual behavior. The transmission dynamics take place within each subpopulation and assume a classic compartmentalization scheme for disease progression similar to those used in several large-scale models of SARS-CoV-2 transmission. Each individual, at any given point in time, is assigned to a compartment corresponding to their particular disease-related state. This state also controls the individual's ability to travel. Individuals transition between compartments through stochastic chain binomial processes. The compartmental structure includes vaccination status and has been extended during the different projections rounds to accommodate new variants through an explicit multi-strain compartmental structure The model calibration is performed for each state through an Approximate Bayesian computation rejection algorithm using as evidence the weekly ground truth data for deaths, hospitalizations and cases.

**NCSU-COVSIM**
North Carolina State University

The COVSIM team uses a stochastic agent-based simulation to model disease spread in North Carolina. Agents are assigned age, race/ethnicity, and high risk health condition attributes to represent the underlying population. Agents have time-varying interactions in households, peer groups (school or workplace), and the community which serve as the driver for disease spread. We use a force of infection model to generate the time until the next infection occurs and select agents to be infected. Once an agent is selected they progress through an extended set of SEIR disease states, which is dependent on the agent's attributes, behaviors, and variants. We model masks, vaccines and booster uptake. We include waning immunity from recovered agents back to the susceptible state. Our model is manually calibrated using disease parameters from literature tailored to North Carolina.

| NotreDame-FRED<br>Notre Dame University |
| --- |
| The Notre Dame team uses an agent-based model based on a modified COVID-19 version of the Framework for Reproducing Epidemic Dynamics (FRED). The model simulates daily interactions among agents in specific locations, such as households, schools, workplaces, or neighborhood areas. The synthetic population is representative of the demographic and geographic characteristics of the simulated States. An agent's health is represented as one of the following states, Susceptible - Exposed - Infectious - Recovered. The model includes circulating variants of SARS-CoV-2 with different characteristics of transmissibility and immunity scape. Recovered individuals are only protected against reinfections with the same variant, and partially protected against reinfection with a different variant for 8 months on average. NPIs are included in the model. Vaccination is included in the model with a prioritization strategy based on age. Three components of the vaccine are modeled, protection against infection, disease given infection, and protection against severe disease given symptoms. The model is calibrated using data on deaths and dominance of circulating variants. |
| **UF-ABM**<br>University of Florida |
| Stochastic, discrete-time agent-based model (ABM) of SARS-CoV-2 transmission in the state of Florida derived from our existing ABM for dengue transmission (*3*, *4*). The model synthetic population includes 20.6 million people and 11.2 million locations (i.e., households, workplaces, schools, long-term care facilities, and hospitals). Transmission occurs via a daily cycle of location-based interactions, (e.g., within household, employee-customer interactions, or between households via social interactions). Individual infections may progress through SEIRD states (with multiple levels of severity, and hospitalization), using existing literature to parameterize infectious outcome probabilities and disease state durations. Probability of infection is modified by location-type-specific infection hazards, infecting viral strain, seasonality, and time-varying personal-protective behaviors. Disease severity depends on the age and health status (i.e. has comorbidity or not) of the agent. A time-varying reporting model serves to replicate empirical dynamic detection and reporting processes. The model represents vaccination through the first 3 doses using a generalized mRNA vaccine and delivers doses in an age-structured way that reflects empirical vaccine delivery in Florida. |
| **UNCC-hierbin**<br>University of North Carolina at Charlotte |
| The UNCC team builds a relatively simple statistical model to retrospectively fit COVID-19 spread. The underlying mechanism is an SEIR-type compartment model and we focus on projecting the cumulative case numbers. In the first few rounds (up to round 9), we fit an exponential model for the cumulative case for each state as well as at nation-level. Then we project the model to the scenario-specific time periods (e.g., 12, 26, or 52 weeks horizon). Hospitalization and death are modeled as a binomial outcome of cases with lags identified from literature. Starting from round 10, we have applied a multivariate long short term memory (LSMT) model to address both long-term and short-term dynamics of the COVID-19 dynamics. Multivariate LSMT also overcomes potential issues of arbitrarily assigned lags between case and hospitalization and death. |
| **USC-SIkJalpha**<br>University of Southern California |
| We proposed the SIkJalpha model as a discrete-time compartmental model. The central idea is that new infections are created through interactions between the currently susceptible population and previously infected population with a rate depending on the time since infection. This is discretized into k bins of size J, i.e., past k periods, each of J units of time, determine k types of infected populations with k different infection rates. The learning of the parameters is done through a weighted linear regression where the weight of old observation decays as a power of alpha. The approach has evolved with time, depending on dataset availability, variables and decisions under consideration that define the scenarios, and new factors that we believed to have a significant impact. True infections were estimated using seroprevalence data until Round 13, after which wastewater data were incorporated. We used a sigmoid curve to fit and extrapolate vaccine adoption until Round 3, and a contagion model after Round 3. The model supported as many vaccine rounds as given in the United States. In Round 14, the model was updated to support an arbitrary number of vaccines. We assumed the vaccine and natural immunity model to be "all-or-nothing" until Round 7, after which a waning immunity model was used. Two variants were supported starting in Round 3 until Round 5, after which an arbitrary number of variants were allowed. We |

disaggregated the new infection time-series into multiple time-series variants using prevalence estimates. Each of the time-series is fitted competing for a common susceptible population to estimate variant-specific transmission rates. Since Round 10, the model supported immune escape variants as well. Additionally, we expanded the model to track all possible vaccines and natural immunity states to estimate the level of susceptibility in the population. The model supports arbitrary age structure.

**UTA-ImmunoSEIRS**
University of Texas at Austin

UTA-ImmunoSEIRS team uses an age-structured COVID-19 SEIRS compartment model that tracks changes in the level of protection acquired from past infection and vaccination. They describe the changes in population-wide immunity resulting from three sources: Delta infections, Omicron infections, and vaccination. The level of each source of protection is explicitly modeled through a state variable. Natural infections increase the infection-acquired protection variables and primary and booster vaccines increase the vaccine-acquired protection variable. The levels of immunity wane at different speeds that are based on published estimates. The variables are used to reduce disease susceptibility and severity by inhibiting infections, symptomatic disease, hospitalizations, and deaths. The efficacy of each form of immunity depends on the relative prevalence of the circulating variants.

**UVA-adaptive**
University of Virginia

UVA-adaptive (as of round 12) is a discrete time SEIR based model with explicit tracking of multiple tiers of vaccine induced immunity, and the latest variant of infection. The overall infection curve produced by the model was converted to reported cases using dynamic case ascertainment and calibrated to match observed ground truth at the state level. Case ascertainment over time was informed by CDC seroprevalence surveys (until Feb 2022), and then coarsely adjusted based on wastewater surveillance and rates of at-home testing. Growth of variants were obtained either from scenario specification (via seeding), or enforced through a prevalence curve based on the growth advantage. Past and ongoing vaccinations are obtained at the state level and appropriately assigned to eligible individuals according to immune stratification. Vaccine efficacy against infection/disease are assumed to be the same, and overall coverage (when relevant) are obtained from scenario specifications. Hospitalizations and death outcomes are obtained by using an age-stratified adjustment over three age groups (0-18, 19-64 and 65+) with scaling factors to match the respective ground truths. Uncertainty bounds are obtained by using an experiment design over other model parameters such as infectious/incubation periods, reporting delay and bounds on ascertainment rates.

**UVA-EpiHiper**
University of Virginia

The UVA-EpiHiper model is an agent-based, individual level networked model. It computes stochastic transmissions of a disease in a synthetic contact network between individuals and stochastic state transitions within each individual following a disease model. Our disease model is an SEIR model expanded with asymptomatic, vaccinated, hospitalized, ventilated, and deceased states, and stratified by age group. Over the rounds, we have extended our disease model to represent multiple variants, waning immunity, and immune escape. The immune waning is modeled as transition from states with natural/vaccinal immunity to a partially susceptible state, and the time to transition is sampled independently for each individual.

Our model is initialized with (i) county level data of prior infections (part of which have waned immunity) and recent confirmed case counts and (ii) state level data of prior vaccinations (part of which have waned immunity). Prior infections are derived from confirmed cases using age stratified ascertainment rates. We have modeled the following NPIs: (i) generic social distancing, the compliance to which changes over rounds; (ii) school closure during winter and summer breaks, and mask mandate in schools; (iii) voluntary home isolation of symptomatic people. Vaccines are applied to eligible individuals according to state level vaccine administration data and projection of future coverage as specified by scenarios. Our model is calibrated at state level targeting the estimated effective reproduction number at the beginning of the projection period.

We run simulations for each state and combine the output to get results of the whole United States. The simulations produce daily infections, hospitalizations, and deaths; and each simulation runs for multiple replicates. We aggregate daily data to get weekly data and compute quantiles for each target from the multiple replicates.

**Table S2: Projections included and excluded from performance analysis.** For projections that are excluded, the percent of all projections from SMH rounds 1-16 is shown in parenthesis. Weeks are defined on Saturdays (and defined here as a range to summarize). Twenty-four percent (24%) of all projections were excluded, primarily in internal training rounds 8 and 10. Some projections could have been excluded for multiple reasons; in these cases, duplicates are included in each percentage calculation. FIPS = Federal Information Processing System codes for U.S. states, CDF = cumulative distribution function.

| Field | Inclusion/Exclusion | Values |
|---|---|---|
| Round | Included | 1, 2, 3, 4, 5, 6, 7, 9, 11, 12, 13, 14, 15, 16 |
| | Excluded: internal training rounds | 8 (7.4% of all projections), 10 (13.3%) |
| Location name (FIPS) | Included | United States (US), Alabama (01), Alaska (02), Arizona (04), Arkansas (05), California (06), Colorado (08), Connecticut (09), Delaware (10), District of Columbia (11), Florida (12), Georgia (13), Hawaii (15), Idaho (16), Illinois (17), Indiana (18), Iowa (19), Kansas (20), Kentucky (21), Louisiana (22), Maine (23),Maryland (24), Massachusetts (25), Michigan (26), Minnesota (27), Mississippi (28), Missouri (29), Montana (30), Nebraska (31), Nevada (32), New Hampshire (33), New Jersey (34), New Mexico (35), New York (36), North Carolina (37), North Dakota (38), Ohio (39), Oklahoma (40), Oregon (41), Pennsylvania (42), Rhode Island (44), South Carolina (45), South Dakota (46), Tennessee (47), Texas (48), Utah (49), Vermont (50), Virginia (51), Washington (53), West Virginia (54), Wisconsin (55), Wyoming (56) |
| | Excluded: territories | American Samoa (60, 0.3%), Guam (66, 0.4%), Northern Mariana Islands (69, 0.4%), Puerto Rico (72, 0.7%), Virgin Islands (78, 0.5%) |
| Weeks | Included | Round 1: 2021-01-09 – 03-27-2021<br>Round 2: 2021-01-30 – 06-19-2021<br>Round 3: 2021-03-13 – 2021-06-19<br>Round 4: 2021-04-03 – 2021-06-19<br>Round 5: 2021-05-08 – 2021-06-19<br>Round 6: 2021-06-05 – 2021-11-27<br>Round 7: 2021-07-10 – 2021-12-18<br>Round 9: 2021-09-18 – 2021-12-18<br>Round 11: 2021-12-25 – 2022-03-12<br>Round 12: 2022-01-15 – 2022-04-02<br>Round 13: 2022-03-19 – 2023-03-11<br>Round 14: 2022-06-11 – 2023-05-20<br>Round 15: 2022-08-06 – 2023-05-06<br>Round 16: 2022-11-05 – 2023-04-29 |
| | Excluded: ground truth data no longer available | Round 13: 2021-03-04 – 2023-03-11 (0.2%)<br>Round 14: 2021-03-04 – 2023-05-20 (2.4%)<br>Round 15: 2021-03-04 – 2023-05-06 (1.5%)<br>Round 16: 2021-03-04 – 2023-04-29 (1.3%) |
| Models | Included | CU-AGE-ST, Ensemble_LOP_untrimmed, Ensemble_LOP, Ensemble, JHU_IDD-CovidSP, JHUAPL-Bucky, Karlen-pypm, MOBS_NEU-GLEAM_COVID, UNCC-hierbin, USC-SIkJalpha, UTA-ImmunoSEIRS, UVA-adaptive, UVA-EpiHiper |
| | Excluded: projections for < 10 locations | NCSU-COVSIM, Rounds 13-15 (<0.1%)<br>NotreDame-FRED, Rounds 11-12 (0.3%)<br>UTA-ImmunoSEIRS, Rounds 12-13 (0.2%)<br>UF-ABM, Round 11 (<0.1%) |

| | | |
|---|---|---|
| | Excluded: point estimates only | OliverWyman-Navigator (<0.1%) |
| | Excluded: projections for < 4 scenarios | IHME-IHME_COVID_model_deaths_unscaled (0.3%) |
| | Excluded: non-increasing CDF | USC-SIkJalpha (Round 1, 0.6%)<br>UNCC-hierbin (Round 5, <0.1%) |
| | Excluded: outside of projection period | JHU_IDD-CovidSP (Rounds 1, 2, 11, 0.1%) |

### S2. Evaluating scenario plausibility

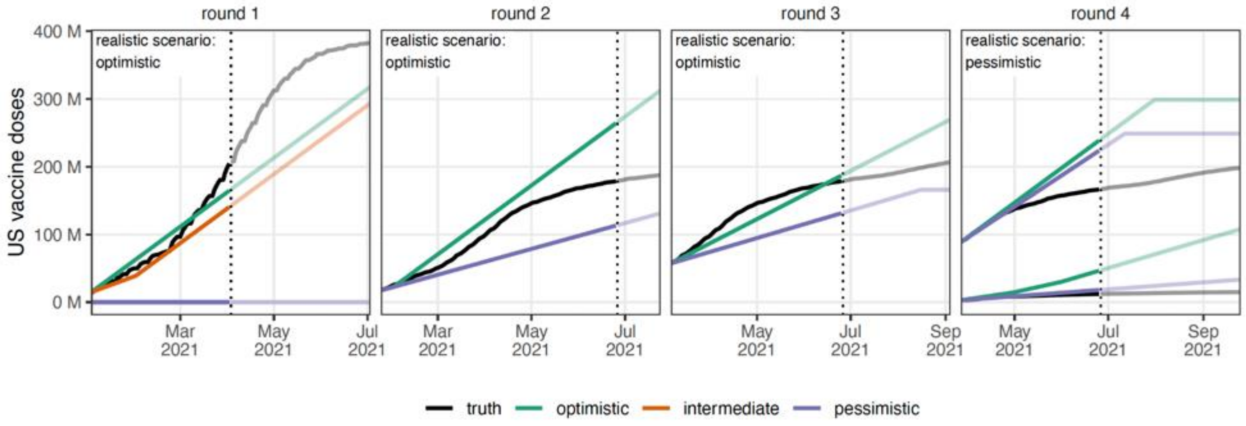


**Figure S2: Comparison of vaccination scenario specifications to observed vaccine uptake, Rounds 1-4.** For rounds with scenarios specifying vaccine distribution or administration, we compared the optimistic (green) and pessimistic (purple) scenarios with the observed (black) U.S. national vaccine coverage. Round 1 also included an intermediate vaccination scenario (orange). Rounds specified vaccination according to different metrics: total distributed doses (Rounds 1 and 2), administered first doses (Round 3), and administered first doses of Moderna & Pfizer and J&J separately (Round 4; J&J is the lower set of 3 lines). Week of truncation is indicated by a dotted vertical line (Round 1 truncated after emergence of the alpha variant, Rounds 2-4 truncated after emergence of the delta variant).

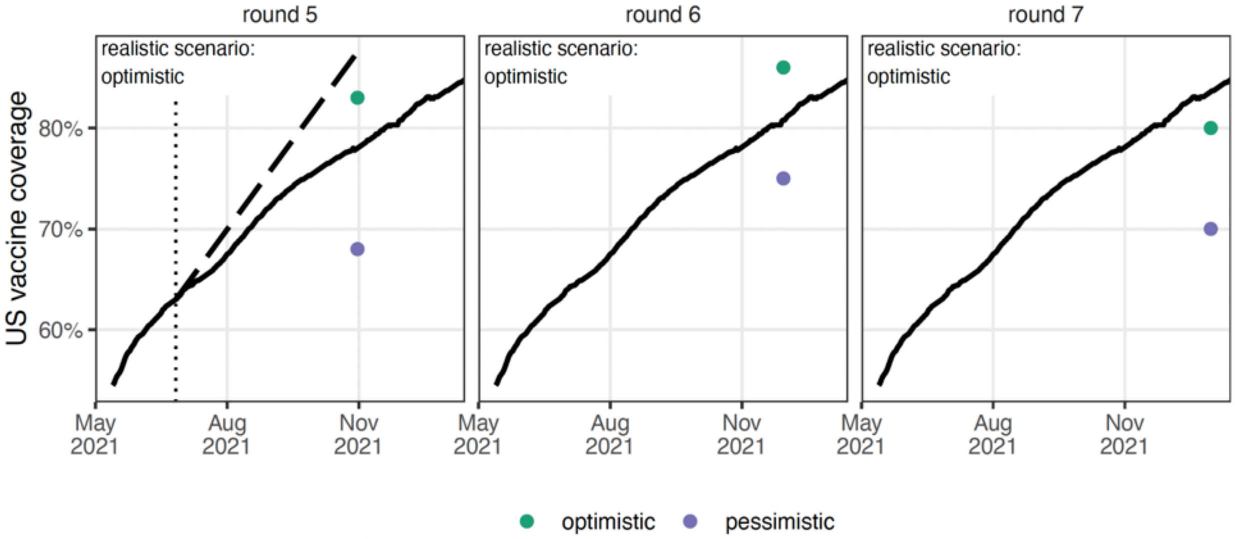


**Figure S3: Comparison of vaccination scenario assumptions to observed vaccine uptake, Rounds 5-7.** For rounds with scenarios specifying vaccine coverage at the end of the projection period, we compared the optimistic (green) and pessimistic (purple) scenarios with the observed (black) US national vaccine coverage. Coverage was specified at the end of the projection period (location of point on x-axis; Round 5: October 31, 2021, Round 6: November 30, 2021, Round 7: December 31, 2021). Round 5 was truncated on June 26, 2021 (vertical dotted line), so we also extended the cumulative coverage gains during the projection period, 0.2% per day, for the rest of the projection period (dashed line). Both methods indicate the optimistic coverage scenario is most realistic for Round 5.

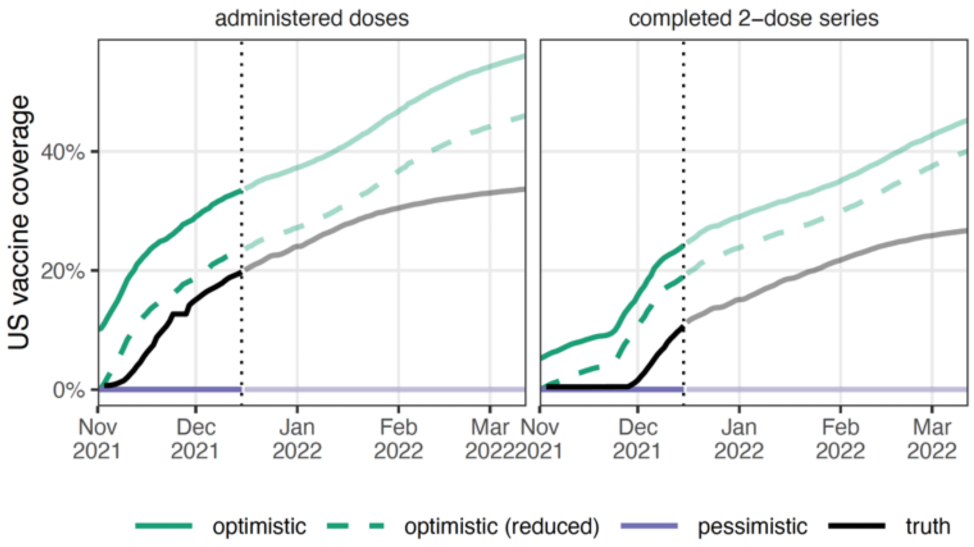


**Figure S4: Comparison of vaccination scenario specifications to observed vaccine uptake, Round 9.** Optimistic (green) scenario assumed vaccination coverage in 5-11 year olds would be equal to uptake in 12-17 year olds, and pessimistic (purple) scenario assumed vaccination would not be approved in 5-11 year olds. Vaccinations were approved on 2 Nov 2021 for 5-11 year olds, and actual uptake in 5-11 year olds is shown (black). Approximately 2.5M vaccines were administered in individuals 12-17 years of age before 13 May 2021 (Pfizer BioNTech was approved for 16+ at the outset). The green lines show coverage including this 2.5M (1.3M completed 2-dose series) and the dashed green lines show coverage since 13 May 2021. Vertical dotted line indicates the timing of the emergence of the omicron variant.

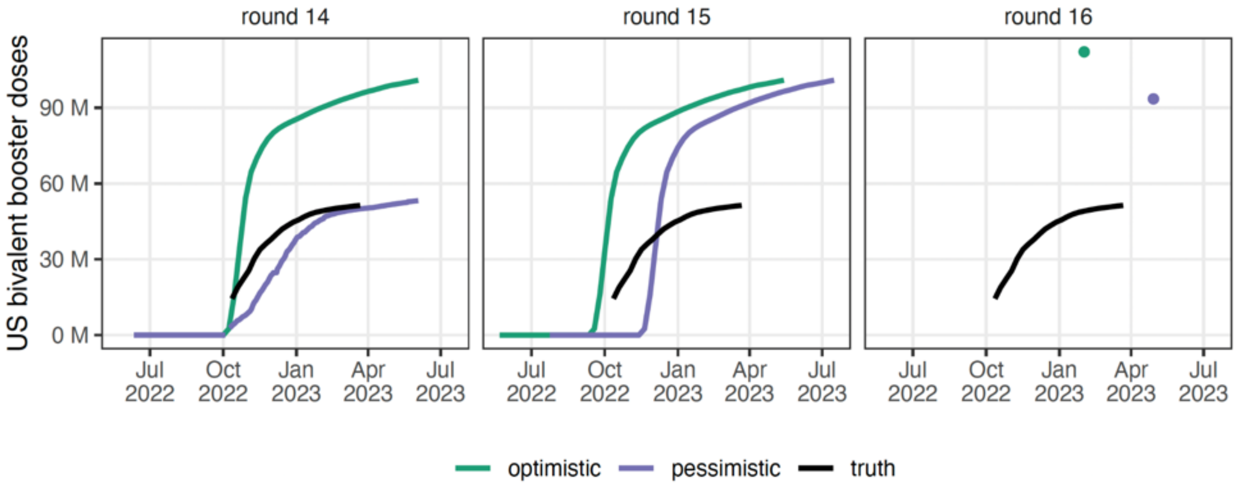


**Figure S5: Comparison of vaccination scenario assumptions to observed vaccine uptake, Rounds 14-16.** For rounds scenarios specifying weekly vaccine bivalent booster coverage (Round 14,15) and those specifying bivalent booster coverage at the end of the projection period (Round 16), we compared the optimistic (green) and pessimistic (purple) scenarios with the observed (black) US national bivalent booster coverage.

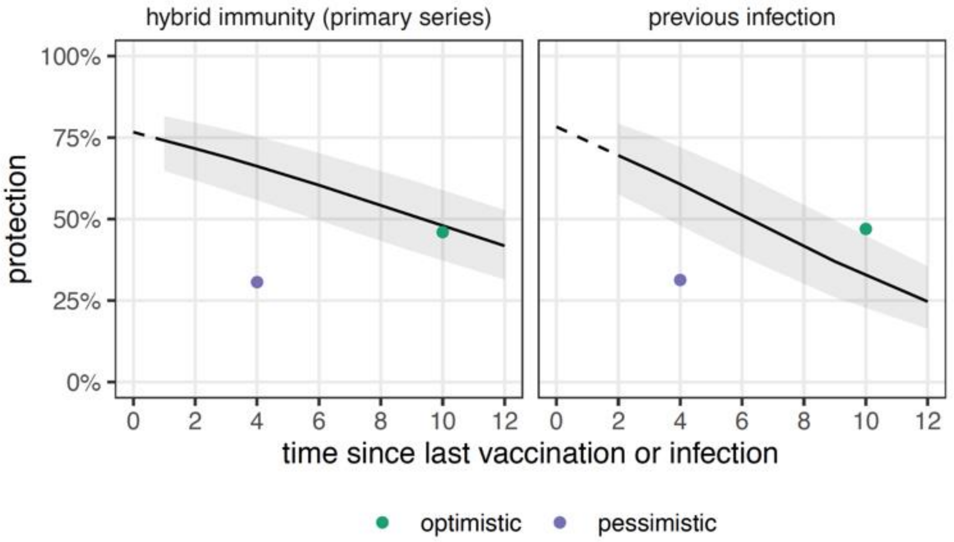


**Figure S6: Comparison of waning scenario assumptions to meta-analysis estimates, Round 13.** Scenario assumptions (green and purple dots) stipulated a median time in weeks to loss of immunity (x-axis) and a level at which protection against symptomatic disease stabilized after immunity loss (y-axis). Scenario assumptions did not distinguish between hybrid immunity (immunity from natural infection and vaccination) and natural immunity, but based on these graphs, the optimistic assumption was most realistic in both situations.

**Table S3: Scenarios by round.** For each round, the major assumptions of each scenario (A-D) and the observed values corresponding to each major assumption are specified. Scenarios retrospectively determined to be "most realistic" are denoted with an asterisk (*). If a single most likely scenario could not be determined (here, due to insufficient data on non-pharmaceutical interventions (NPIs) for scenarios in Rounds 3-5), multiple most likely scenarios were considered. Projection period is represented weekly, where weeks are defined on Saturdays. For a full list of scenario specifications by round, see https://github.com/midas-network/covid19-scenario-modeling-hub/tree/master/previous-rounds.

| **Round 1**<br>Jan 9 to Jul 3, 2021 (alpha variant truncation on Apr 3, 2021) |
| --- |
| **Scenario A: optimistic***<br>*Social distancing:* NPIs continue for six weeks from their start date, interventions step down from baseline to lowest levels seen since September 2020 over two one-month steps<br>Vaccination: 50 million total doses distributed per month (~292 million total first doses distributed, ~140 million total doses distributed before truncation); 95% vaccine efficacy after two doses, 50% after one dose |
| **Scenario B: moderate**<br>*Social distancing:* NPIs continue for three weeks from their start date, interventions step down from baseline to lowest levels seen since May 2020 over two one-month steps<br>*Vaccination:* 25 million total doses distributed in January 2021, 50 million for all other months (~273 million first doses distributed, ~122 million total doses distributed before truncation); 70% vaccine efficacy after two doses, 20% after one dose |
| **Scenario C: fatigue**<br>*Social distancing:* NPIs continue for three weeks from their start date, interventions step down from baseline to 5% below lowest levels seen since May 2020 over two one-month steps<br>*Vaccination:* 25 million total doses distributed in January 2021, 50 million for all other months, no more than 50% of any priority group accepts the vaccine (~273 million first doses distributed, ~122 million total doses distributed before truncation); 95% vaccine efficacy after two doses, 50% after one dose |

**Scenario D: counterfactual**

*Social distancing:* NPIs continue for three weeks from their start date, interventions step down from baseline to lowest levels seen since May. 2020 over two one-month steps
*Vaccination:* no vaccines distributed (0 total doses distributed)

**Observed**

*Social distancing:* despite the availability on mobility and policy data, the specificity of these data does not match that of our NPI scenarios; therefore we retain both moderate and low NPI scenarios as likely
*Vaccination:* ~361 million total doses distributed; ~186 million first doses distributed before truncation; 50% coverage achieved in 65+ by May 5, 2021; 80-90% vaccine efficacy against disease after two doses (*5*), 30% after one dose(*6*)

**Round 2**

Jan 30 to Jul 25, 2021 (delta variant truncation on Jun 26, 2021)

**Scenario A: optimistic, no variant**

*Social distancing:* NPIs continue for six weeks from their start date, interventions step down from baseline to lowest levels seen since Sept. 2020 over two one-month steps
*Vaccination:* 25 million doses administered per month after January 2021 (~291 million first doses administered, ~244 million first doses administered before truncation); 95% vaccine efficacy after two doses, 50% after one dose
*Variant:* no new variant

**Scenario B: optimistic, variant***

*Social distancing:* NPIs continue for six weeks from their start date, interventions step down from baseline to lowest levels seen since Sept. 2020 over two one-month steps
*Vaccination:* 25 million doses administered per month after January 2021 (~291 million first doses administered, ~244 million first doses administered before truncation); 95% vaccine efficacy after two doses, 50% after one dose
*Variant:* new variant is 1.5 times more transmissible than current strains; 50% dominant by Mar 15, 2020 and 100% dominant by May 1, 2020

**Scenario C: fatigue, no variant**

*Social distancing:* NPIs continue for three weeks from their start date, interventions step down from baseline to 5% below lowest levels seen since May 2020 over two one-month steps
*Vaccination:* monthly administration follows rates seen in January 2021; no more than 50% of any priority group accepts the vaccine (~124 million first doses administered, ~104 million first doses administered before truncation); 95% vaccine efficacy after two doses, 50% after one dose
*Variant:* no new variant

**Scenario D: fatigue, variant**

*Social distancing:* NPIs continue for three weeks from their start date, interventions step down from baseline to 5% below lowest levels seen since May 2020 over two one-month steps
*Vaccination:* monthly administration follows rates seen in January 2021; no more than 50% of any priority group accepts the vaccine (~114 million first doses administered, ~96 million first doses administered before truncation); 95% vaccine efficacy after two doses, 50% after one dose
*Variant:* new variant is 1.5 times more transmissible than current strains; 50% dominant by Mar 15 2020 and 100% dominant by May 1 2020

**Observed**

*Social distancing:* despite the availability on mobility and policy data, the specificity of these data does not match that of our NPI scenarios; therefore we retain both moderate and low NPI scenarios as likely
*Vaccine availability:* ~170 million first doses administered; ~161 million first doses administered before truncation; 50% coverage achieved in 65+ by May 5, 2021;
*Variant:* transmissibility increases estimated to be 24% (*7*), 40-50% (*8*), and 43-90% (*9*); alpha variant reaches 50% in US on Mar 30, 2021 (*10*); vaccine efficacy against disease after two doses (*5*), 30% after one dose (*6*)

**Round 3**

Mar 13 to Sep 4, 2021 (delta variant truncation on Jun 26, 2021)

**Scenario A: high vaccination, moderate NPI***

*Non-pharmaceutical interventions (NPIs):* NPIs decline gradually over a period of 5 months ending in August 2021 at 50% of the effectiveness observed in February 2021
*Vaccination:* 35 million first doses administered per month, no more than 90% of any population group receives the vaccine (~212 million first doses administered, ~130 million first doses administered before truncation); 95% vaccine efficacy against disease after two doses, 90% after one dose

**Scenario B: high vaccination, low NPI***

*NPIs:* NPIs decline gradually over a period of 5 months ending in August 2021 at 20% of the effectiveness observed in February 2021
*Vaccination:* 35 million first doses administered per month, no more than 90% of any population group receives the vaccine (~212 million first doses administered, ~130 million first doses administered before truncation); 95% vaccine efficacy against disease after two doses, 90% after one dose

**Scenario C: low vaccination, moderate NPI**

*NPIs:* NPIs decline gradually over a period of 5 months ending in August 2021 at 50% of the effectiveness observed in February 2021
*Vaccination:* 20 million first doses administered per month, no more than 50% of any population group receives the vaccine (~108 million total doses administered, ~75 million total doses administered before truncation); 75% vaccine efficacy against disease after two doses, 50% after one dose

**Scenario D: low vaccination, low NPI**

*NPIs:* NPIs decline gradually over a period of 5 months ending in August 2021 at 20% of the effectiveness observed in February 2021
*Vaccination:* 20 million first doses administered per month, no more than 50% of any population group receives the vaccine (~108 million total doses administered, ~75 million total doses administered before truncation); 75% vaccine efficacy against disease after two doses, 50% after one dose

**Observed**

*NPIs:* despite the availability on mobility and policy data, the specificity of these data does not match that of our NPI scenarios; therefore we retain both moderate and low NPI scenarios as likely
*Vaccination:* ~150 million first doses administered, ~122 million total first doses administered before truncation (excluding J&J, which was not considered in scenarios); 50% coverage is reached nationally on May 13, 2021, 90% coverage is never reached; 80-90% vaccine efficacy against disease after two doses (*5*), 30% after one dose (*6*)

**Round 4**

Apr 3 to Sep 25, 2021 (delta variant truncation on Jun 26, 2021)

**Scenario A: high vaccination, moderate NPI**

*Non-pharmaceutical interventions (NPIs):* NPIs decline gradually over a period of 6 months ending in September 2021 at 50% of the effectiveness observed in March 2021
*Vaccination:* 50 million mRNA (Moderna/Pfizer) first doses administered monthly; 10 million Johnson& Johnson first doses administered in April, 15 million in May, and 20 million month from June-September; no more than 90% of any population group receives the vaccine (~210 million first doses administered of Moderna/Pfizer, and ~105 million doses administered of Johnson & Johnson, 152 million first doses administered of Moderna/Pfizer, and ~44 million doses administered of Johnson & Johnson before truncation)
95% mRNA vaccine efficacy against disease after 2 doses, 75% after one dose; 70% J&J vaccine efficacy against symptoms after one dose

**Scenario B: high vaccination, low NPI**

*NPIs:* NPIs decline gradually over a period of 6 months ending in September 2021 at 20% of the effectiveness observed in March 2021
*Vaccination:* 50 million mRNA first doses administered monthly; 10 million Johnson & Johnson first doses administered in April, 15 million in May, and 20 million month from June-September; no more than 90% of any population group receives the vaccine (~210 million first doses administered of Moderna/Pfizer, and ~105 million doses administered of Johnson & Johnson, 152 million first doses administered of Moderna/Pfizer, and ~44 million doses administered of Johnson & Johnson before truncation)
95% mRNA vaccine efficacy against disease after 2 doses, 75% after one dose; 70% J&J vaccine efficacy against symptoms after one dose

| |
|---|
| **Scenario C: low vaccination, moderate NPI***<br>*NPIs:* NPIs decline gradually over a period of 6 months ending in September 2021 at 50% of the effectiveness observed in March 2021<br>*Vaccination: Vaccination:* 45 million mRNA first doses administered monthly; 5 million Johnson & Johnson first doses administered monthly; no more than 75% of any population group receives the vaccine (~160 million first doses administered of Moderna/Pfizer, and ~30 million doses administered of Johnson & Johnson, 136 million first doses administered of Moderna/Pfizer, and ~15 million doses administered of Johnson & Johnson before truncation)<br>85% mRNA vaccine efficacy against disease after 2 doses, 50% after one dose; 60% J&J vaccine efficacy against symptoms after one dose |
| **Scenario D: low vaccination, low NPI***<br>*NPIs:* NPIs decline gradually over a period of 6 months ending in September 2021 at 20% of the effectiveness observed in March 2021<br>*Vaccination: Vaccination:* 45 million mRNA first doses administered monthly; 5 million Johnson & Johnson first doses administered monthly; no more than 75% of any population group receives the vaccine (~160 million first doses administered of Moderna/Pfizer, and ~30 million doses administered of Johnson & Johnson, 136 million first doses administered of Moderna/Pfizer, and ~15 million doses administered of Johnson & Johnson before truncation)<br>85% mRNA vaccine efficacy against disease after 2 doses, 50% after one dose; 60% J&J vaccine efficacy against symptoms after one dose |
| **Observed**<br>*NPIs:* despite the availability on mobility and policy data, the specificity of these data does not match that of our NPI scenarios; therefore we retain both moderate and low NPI scenarios as likely<br>*Vaccination:* ~110 million mRNA first doses administered, and ~12 million first doses administered of J&J, ~78 million mRNA first doses administered and 9 million J&J first doses administered before truncation; 75% coverage is reached nationally on September 25, 2021, 90% coverage is never reached; 80-90% vaccine efficacy against disease after two doses (*5*), 30% after one dose (*6*) |
| **Round 5**<br>May 08 to Oct 30, 2021 (delta variant truncation on Jun 26, 2021) |
| **Scenario A: high vaccination, moderate NPI***<br>*Non-pharmaceutical interventions (NPIs):* NPIs decline gradually over a period of 6 months ending in October 2021 at 50% of the effectiveness observed in April 2021<br>*Vaccination:* national saturation at 83% of vaccine-eligible population by October 31, 2021 |
| **Scenario B: high vaccination, low NPI***<br>*NPIs:* NPIs decline gradually over a period of 6 months ending in October 2021 at 80% of the effectiveness observed in April 2021<br>*Vaccination:* national saturation at 83% of vaccine-eligible population by October 31, 2021 |
| **Scenario C: low vaccination, moderate NPI**<br>*NPIs:* NPIs decline gradually over a period of 6 months ending in October 2021 at 50% of the effectiveness observed in April 2021<br>*Vaccination:* national saturation at 68% of vaccine-eligible population by October 31, 2021 |
| **Scenario D: low vaccination, low NPI**<br>*NPIs:* NPIs decline gradually over a period of 6 months ending in October 2021 at 80% of the effectiveness observed in April 2021<br>*Vaccination:* national saturation at 68% of vaccine-eligible population by October 31, 2021 |
| **Observed**<br>*NPIs:* despite the availability on mobility and policy data, the specificity of these data does not match that of our NPI scenarios; therefore we retain both moderate and low NPI scenarios as likely<br>*Vaccination:* national saturation at 78% of vaccine-eligible population by October 31, 2021 (note: because this round is truncated early in the projection period, we also consider the cumulative coverage gains during the projection period, 0.2% per day; if we extend this for the duration of the projection period, we have an expected coverage of 88%) |

**Round 6**
Jun 5 to Nov 27, 2021 (not truncated)

**Scenario A: high vaccination, low variant impact**
*Variant:* B.1.617 is 1.2 times more transmissible than B.1.1.7
*Vaccination:* national saturation at 86% of vaccine-eligible population by November 30, 2021; 90% vaccine efficacy against disease after two doses, 50% after one dose

**Scenario B: high vaccination, high variant impact***
*Variant:* B.1.617 is 1.6 times more transmissible than B.1.1.7
*Vaccination:* national saturation at 86% of vaccine-eligible population by November 30, 2021; 90% vaccine efficacy against disease after two doses, 50% after one dose

**Scenario C: low vaccination, low variant impact**
*Variant:* B.1.617 is 1.2 times more transmissible than B.1.1.7
*Vaccination:* national saturation at 75% of vaccine-eligible population by November 30, 2021; 90% vaccine efficacy against disease after two doses, 50% after one dose

**Scenario D: low vaccination, high variant impact**
*Variant:* B.1.617 is 1.6 times more transmissible than B.1.1.7
*Vaccination:* national saturation at 75% of vaccine-eligible population by November 30, 2021; 90% vaccine efficacy against disease after two doses, 50% after one dose

**Observed**
*Variant:* B.1.617 is estimated in a meta-analysis to be approximately 1.5 times more transmissible than B.1.1.7(*11*), though estimates from Earnest *et al.* (*12*) are higher (1.63-2.67)
*Vaccination:* national saturation at 81% of vaccine-eligible population on November 30, 2021; 80-90% vaccine efficacy against disease after two doses (*5*), 30% after one dose (*6*)

**Round 7**
Jul 10, 2021 to Jan 1, 2022 (omicron variant truncation on Dec 25, 2021)

**Scenario A: high vaccination, low variant impact**
*Variant:* B.1.617 is 1.4 times more transmissible than B.1.1.7
*Vaccination:* national saturation at 80% of vaccine-eligible population by December 31, 2021

**Scenario B: high vaccination, high variant impact***
*Variant:* B.1.617 is 1.6 times more transmissible than B.1.1.7
*Vaccination:* national saturation at 80% of vaccine-eligible population by December 31, 2021

**Scenario C: low vaccination, low variant impact**
*Variant:* B.1.617 is 1.4 times more transmissible than B.1.1.7
*Vaccination:* national saturation at 70% of vaccine-eligible population by December 31, 2021

**Scenario D: low vaccination, high variant impact**
*Variant* B.1.617 is 1.6 times more transmissible than B.1.1.7
*Vaccination:* national saturation at 70% of vaccine-eligible population by December 31, 2021

**Observed**
*Variant:* B.1.617 is estimated in a meta-analysis to be approximately 1.5 times more transmissible than B.1.1.7(*11*), though estimates from Earnest *et al.* (*12*) are higher (1.63-2.67)
*Vaccination:* national saturation at 84% of vaccine-eligible population by December 31, 2021 (83% on December 25, 2021 when projections are truncated)

**Round 9**
Sep 18, 2021 to Mar 12, 2022 (omicron variant truncation on Dec 25, 2021)

**Scenario A: Childhood vaccination, no variant***
*Variant:* the same mix of variants continues to circulate

*Childhood vaccination:* vaccination among 5-11 year old children is approved and begins on Nov. 1, 2021. State-level uptake reflects that observed in the 12-17 year old age group since distribution to this group began on May 13, 2021. This is equivalent to 46% national coverage of full series at end of projection period; 27% national coverage of full series before truncation. Excluding 2.5M individuals from 12-17 that were vaccinated before approval on 13 May 2021 (Pfizer BioNTech was approved for 16+ at the outset), 41% national coverage at the end of the projection period, and 22% national coverage before truncation.

**Scenario B: Childhood vaccination, variant**

*Variant:* a variant 1.5 times more transmissible emerges, comprising 1% of circulating viruses on Nov. 15, 2021
*Childhood vaccination:* vaccination among 5-11 year old children is approved and begins on Nov. 1, 2021. State-level uptake reflects that observed in the 12-17 year old age group since distribution to this group began on May 13, 2021. This is equivalent to 46% national coverage of full series at end of projection period; 27% national coverage of full series before truncation. Excluding 2.5M individuals from 12-17 that were vaccinated before approval on 13 May 2021 (Pfizer BioNTech was approved for 16+ at the outset), 41% national coverage at the end of the projection period, and 22% national coverage before truncation.

**Scenario C: No childhood vaccination, no variant**

*Variant:* the same mix of variants continues to circulate
*Childhood vaccination:* vaccination among 5-11 year old children is not approved

**Scenario D: No childhood vaccination, variant**

*Variant:* a variant 1.5 times more transmissible emerges, comprising 1% of circulating viruses on Nov. 15, 2021
*Childhood vaccination:* vaccination among 5-11 year old children is not approved

**Observed**

*Variant:* no variant emergence before omicron
*Childhood vaccination:* Vaccination was approved in 5-11 year old children on Nov 2, 2021(*13*). 27% national of full series for 5-11 year olds at the end of the projection period, and 13% national coverage before truncation.

**Round 11**

Dec 25, 2021 to Mar 12, 2022 (not truncated)

**Scenario A: optimistic severity, high immune escape**

*Variant transmissibility/immune escape:* Omicron has the same seasonally-adjusted R0 as Delta (R0=6); 80% of previously immune are susceptible to infection
*Variant severity:* Among naive, Omicron has a 50% less severe infection than Delta; among previously infected or vaccinated, residual protection from hospitalization is 85% and from death is 95%

**Scenario B: optimistic severity, low immune escape***

*Variant transmissibility/immune escape:* Omicron has 1.6 times the seasonally-adjusted R0 as Delta (R0=10); 50% of previously immune are susceptible to infection
*Variant severity:* Among naive, Omicron has a 50% less severe infection than Delta; among previously infected or vaccinated, residual protection from hospitalization is 85% and from death is 95%

**Scenario C: pessimistic severity, high immune escape**

*Variant transmissibility/immune escape:* Omicron has the same seasonally-adjusted R0 as Delta (R0=6); 80% of previously immune are susceptible to infection
*Variant severity:* Among naive, there is no change in severity of infection compared to Delta; among previously infected or vaccinated, residual protection from hospitalization is 70% and from death is 85%

**Scenario D: pessimistic severity, low immune escape**

*Variant transmissibility/immune escape:* Omicron has 1.6 times the seasonally-adjusted R0 as Delta (R0=10); 50% of previously immune are susceptible to infection
*Variant severity:* Among naive, there is no change in severity of infection compared to Delta; among previously infected or vaccinated, residual protection from hospitalization is 70% and from death is 85%

**Observed**

*Variant transmissibility/immune escape:* Omicron R0 is more than twice that of Delta; immune escape is less than 50% (*14*)
*Variant severity:* Severity is 66% lower for Omicron compared to Delta based on UK deaths and 50-80% lower for

| |
|---|
| a range of severe outcomes in the US(*15*, *16*). Vaccine effectiveness against Omicron hospitalization is 60-91% for 2 doses and 85-95% for 3 doses. Vaccine effectiveness against Omicron death is 80% for 2 doses (mRNA vaccines) and 88-94% for 3 doses(*17*) |
| **Round 12**<br>Jan 15 to Apr 2, 2022 (not truncated) |
| **Scenario A: optimistic severity, high immune escape**<br>*Variant immune escape:* 80% of previously immune are susceptible to infection<br>*Variant severity:* Omicron has a 70% less severe infection than Delta among all immune classes |
| **Scenario B: optimistic severity, low immune escape***<br>*Variant immune escape:* 50% of previously immune are susceptible to infection<br>*Variant severity:* Omicron has a 70% less severe infection than Delta among all immune classes |
| **C: pessimistic severity, high immune escape**<br>*Variant immune escape:* 80% of previously immune are susceptible to infection<br>*Variant severity:* Omicron has a 30% less severe infection than Delta among all immune classes |
| **D: pessimistic severity, low immune escape**<br>*Variant immune escape:* 50% of previously immune are susceptible to infection<br>*Variant severity:* Omicron has a 30% less severe infection than Delta among all immune classes |
| **Observed**<br>*Variant immune escape:* Omicron immune escape is less than 50%(*14*)<br>*Variant severity:* Severity is 66% lower for Omicron compared to Delta based on UK deaths and 50-80% lower for a range of severe outcomes in the US(*15*, *16*). Vaccine effectiveness against Omicron hospitalization is 60-91% for 2 doses and 85-95% for 3 doses. Vaccine effectiveness against Omicron death is 80% for 2 doses (mRNA vaccines) and 88-94% for 3 doses(*17*) |
| **Round 13**<br>Mar 13, 2022 to Mar 11, 2023 (not truncated) |
| **Scenario A: optimistic immunity waning, no new immune escape variant***<br>*Immunity waning:* median transition time to partially immune state is 10 months. In the partially immune state, there is a 40% reduction in protection from levels reported immediately after exposure (vaccination or infection).<br>*Variant:* current mix of strains continue to circulate for the duration of the projection period. |
| **Scenario B: optimistic immunity waning, new immune escape variant***<br>*Immunity waning:* median transition time to partially immune state is 10 months. In the partially immune state, there is a 40% reduction in protection from levels reported immediately after exposure (vaccination or infection).<br>*Variant:* new variant emerges on May 1, 2022 that has 30% immune escape and the same intrinsic transmissibility and severity as Omicron. There is a continuous influx of 50 weekly infections of this variant for the following 16 weeks. |
| **Scenario C: pessimistic immunity waning, no new immune escape variant**<br>*Immunity waning:* median transition time to partially immune state is 4 months. In the partially immune state, there is a 60% reduction in protection from levels reported immediately after exposure (vaccination or infection).<br>*Variant:* current mix of strains continue to circulate for the duration of the projection period |
| **Scenario D: pessimistic immunity waning, new immune escape variant**<br>*Immunity waning:* median transition time to partially immune state is 4 months. In the partially immune state, there is a 60% reduction in protection from levels reported immediately after exposure (vaccination or infection).<br>*Variant:* new variant emerges on May 1, 2022 that has 30% immune escape and the same intrinsic transmissibility and severity as Omicron. There is a continuous influx of 50 weekly infections of this variant for the following 16 weeks. |
| **Observed**<br>*Immunity waning:* Meta-analysis estimates(*18*) indicate reduction from levels reported immediately after exposure is 22% after 4 months and 58% after 10 months for immunity from previous infection. Estimates are 14% after 4 |

| |
|---|
| months and 37% after 10 months for hybrid immunity (infection and vaccination). Protection levels at 10 months were linearly interpolated between estimates provided at 9 and 12 months.<br>*Variant:* due to lack of data on introduction and immune escape of different Omicron sublineages, we retain both immune escape variant scenarios as likely |
| **Round 14**<br>Jun 5, 2022 to Jun 3, 2023 (not truncated) |
| **Scenario A: age-restricted booster recommendations, no new immune escape variant***<br>*Booster:* a third, reformulated booster is recommended on October 1, 2022 for adults 50+ and those with chronic conditions. Booster uptake among previously vaccinated is 15% lower than uptake of the first booster (~58 million doses by June 3, 2023, ~50 million by March 15, 2023). Recommended time between booster doses is maintained.<br>*Variant:* current mix of strains continue to circulate for the duration of the projection period. |
| **Scenario B: age-restricted booster recommendations, new immune escape variant***<br>*Booster:* a third, reformulated booster is recommended on October 1, 2022 for adults 50+ and those with chronic conditions. Booster uptake among previously vaccinated is 15% lower than uptake of the first booster (~58 million doses by June 3, 2023, ~50 million by March 15, 2023). Recommended time between booster doses is maintained.<br>*Variant:* new variant emerges on Sep 4, 2022 that has 40% immune escape against infection, a 20% increased risk of hospitalization and death relative to Omicron. There is a continuous influx of 50 weekly infections of this variant from Sep 4, 2022- Dec 24, 2022. |
| **Scenario C: broad booster recommendations, no new immune escape variant**<br>*Booster:* a third, reformulated booster is recommended on October 1, 2022 for all adults 18+. Booster uptake among previously vaccinated is 10% lower than uptake of the 2021-2022 flu vaccine coverage (~94 million doses by June 3, 2023, ~94 million by March 15, 2023). Boosters are recommended regardless of time since previous receipt of a booster.<br>*Variant:* current mix of strains continue to circulate for the duration of the projection period. |
| **Scenario D: broad booster recommendations, new immune escape variant**<br>*Booster:* a third, reformulated booster is recommended on October 1, 2022 for all adults 18+. Booster uptake among previously vaccinated is 10% lower than uptake of the 2021-2022 flu vaccine coverage (~101 million doses by June 3, 2023, ~94 million by March 15, 2023). Boosters are recommended regardless of time since previous receipt of a booster.<br>*Variant:* new variant emerges on Sep 4, 2022 that has 40% immune escape against infection, a 20% increased risk of hospitalization and death relative to Omicron. There is a continuous influx of 50 weekly infections of this variant from Sep 4, 2022- Dec 24, 2022. |
| **Observed**<br>*Booster:* Third, reformulated booster was recommended by ACIP on September 1, 2022 for all individuals 12+ (Pfizer) or 18+ (Moderna)(*19*). As of March 20, 2023, ~51 million bivalent booster doses were administered. The most plausible scenario was selected based on vaccine uptake.<br>*Variant:* due to lack of data on introduction and immune escape of different Omicron sublineages, we retain both immune escape variant scenarios as likely. |
| **Round 15**<br>Jul 31, 2022 to May 6, 2022 (not truncated) |
| **Scenario A: early boosters, no new immune escape variant***<br>*Booster:* Reformulated boosters are available to all adults on Sep 11, 2022. Uptake of reformulated boosters among previously vaccinated is 10% lower than uptake of the 2021-2022 flu vaccine coverage (~101 million doses). Boosters are recommended regardless of time since previous receipt of a booster.<br>*Variant:* current mix of strains continue to circulate for the duration of the projection period. |
| **Scenario B: early boosters, new immune escape variant***<br>*Booster:* Reformulated boosters are available to all adults on Sep 11, 2022. Uptake of reformulated boosters among previously vaccinated is 10% lower than uptake of the 2021-2022 flu vaccine coverage (~101 million doses). Boosters are recommended regardless of time since previous receipt of a booster. |

*Variant:* new variant emerges on Sep 4, 2022 that has 40% immune escape against infection, a 20% increased risk of hospitalization and death relative to Omicron. There is a continuous influx of 50 weekly infections of this variant from Sep 4, 2022- Dec 24, 2022.

**Scenario C: late boosters, no new immune escape variant**

*Booster:* Reformulated boosters are available to all adults on Nov 13, 2022. Uptake of reformulated boosters among previously vaccinated is 10% lower than uptake of the 2021-2022 flu vaccine coverage (~101 million doses). Boosters are recommended regardless of time since previous receipt of a booster.
*Variant:* current mix of strains continue to circulate for the duration of the projection period.

**Scenario D: late boosters, new immune escape variant**

*Booster:* Reformulated boosters are available to all adults on Nov 13, 2022. Uptake of reformulated boosters among previously vaccinated is 10% lower than uptake of the 2021-2022 flu vaccine coverage (~101 million doses). Boosters are recommended regardless of time since previous receipt of a booster.
*Variant:* new variant emerges on Sep 4, 2022 that has 40% immune escape against infection, a 20% increased risk of hospitalization and death relative to Omicron. There is a continuous influx of 50 weekly infections of this variant from Sep 4, 2022- Dec 24, 2022.

**Observed**

*Booster*: Third, reformulated booster was recommended by ACIP on September 1, 2022 for all individuals 12+ (Pfizer) or 18+ (Moderna)(*19*). As of March 20, 2023, ~51 million bivalent booster doses were administered.
*Variant:* due to lack of data on introduction and immune escape of different Omicron sublineages, we retain both immune escape variant scenarios as likely.

**Round 16**

Oct 30, 2022 to Apr 29, 2023 (not truncated)

**Scenario A: high booster uptake, moderate immune escape variant**

*Booster:* Reformulated booster coverage plateaus at 90% of flu vaccination levels by Feb 1, 2022 (~93 million doses).
*Variants:* Level 5 variants have 25% immune escape from BA.5.2, with seeding based on combined observed prevalence of Level 5 variants at the start of the projection period. No change in severity given symptomatic infection.

**Scenario B: high booster uptake, high immune escape variant**

*Booster:* Reformulated booster coverage plateaus at 90% of flu vaccination levels by Feb 1, 2022 (~93 million doses).
*Variants:* Level 6/7 variants have 50% immune escape from BA.5.2, with seeding based on combined observed prevalence of Level 6/7 variants at the start of the projection period. No change in severity given symptomatic infection.

**C: low booster uptake, moderate immune escape variant***

*Booster:* Reformulated booster uptake plateaus at levels mirroring first booster uptake by April 29, 2023 (112 million doses).
*Variants:* Level 5 variants have 25% immune escape from BA.5.2, with seeding based on combined observed prevalence of Level 5 variants at the start of the projection period. No change in severity given symptomatic infection.

**D: low booster uptake, high immune escape variant***

*Booster:* Reformulated booster uptake plateaus at levels mirroring first booster uptake by April 29, 2023 (~112 million doses).
*Variants:* Level 6/7 variants have 50% immune escape from BA.5.2, with seeding based on combined observed prevalence of Level 6/7 variants at the start of the projection period. No change in severity given symptomatic infection.

**Observed**

*Booster:* ~49 million bivalent booster doses had been administered on February 1, 2023, and ~51 million doses by March 20, 2023.
*Variants:* due to lack of data on introduction and immune escape of different Omicron sublineages, we retain both immune escape variant scenarios as likely.

### S3. Comparative models

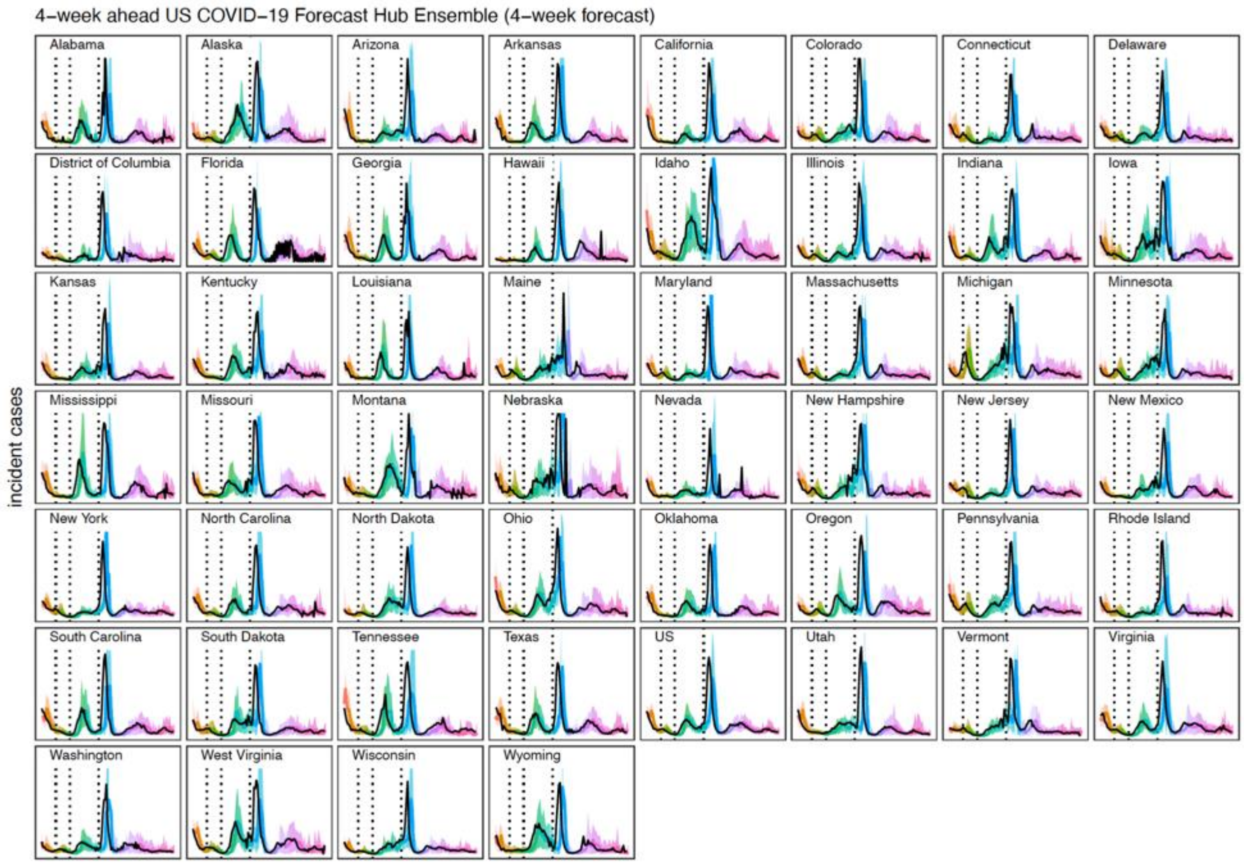


**Figure S7: 4-week forecast comparative model projections for incident cases.** Across locations, median (line) and 95% prediction interval (ribbon) for each round is shown with a different color. Black curve represents observations, and dotted black vertical lines represent the truncation dates for Alpha (2021-04-03), Delta (2021-06-26), and Omicron (2021-12-25).

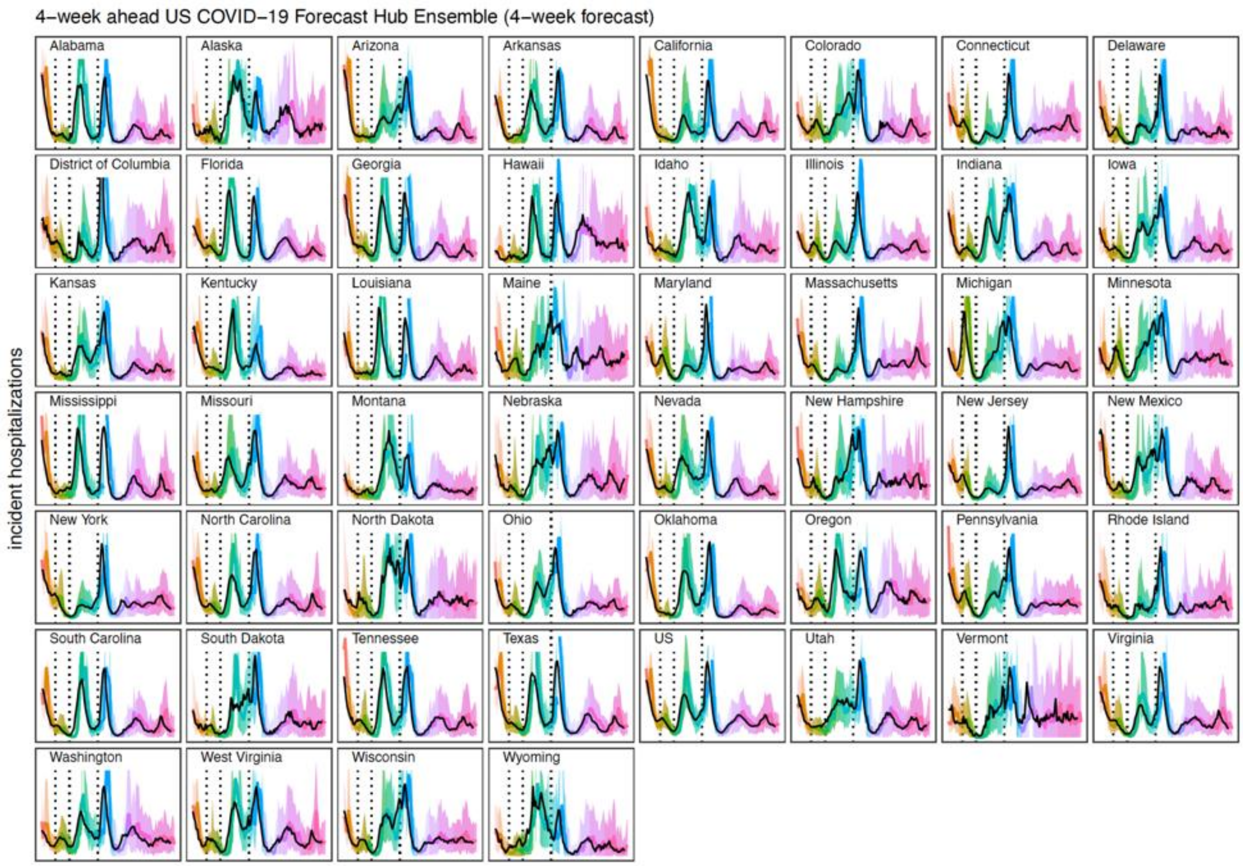


**Figure S8: Highly-informed comparative model projections for incident hospitalizations.** Across locations, median (line) and 95% projection interval (ribbon) for each round is shown with a different color. Black curve represents observations, and dotted black vertical lines represent the truncation dates for Alpha (2021-04-03), Delta (2021-06-26), and Omicron (2021-12-25).

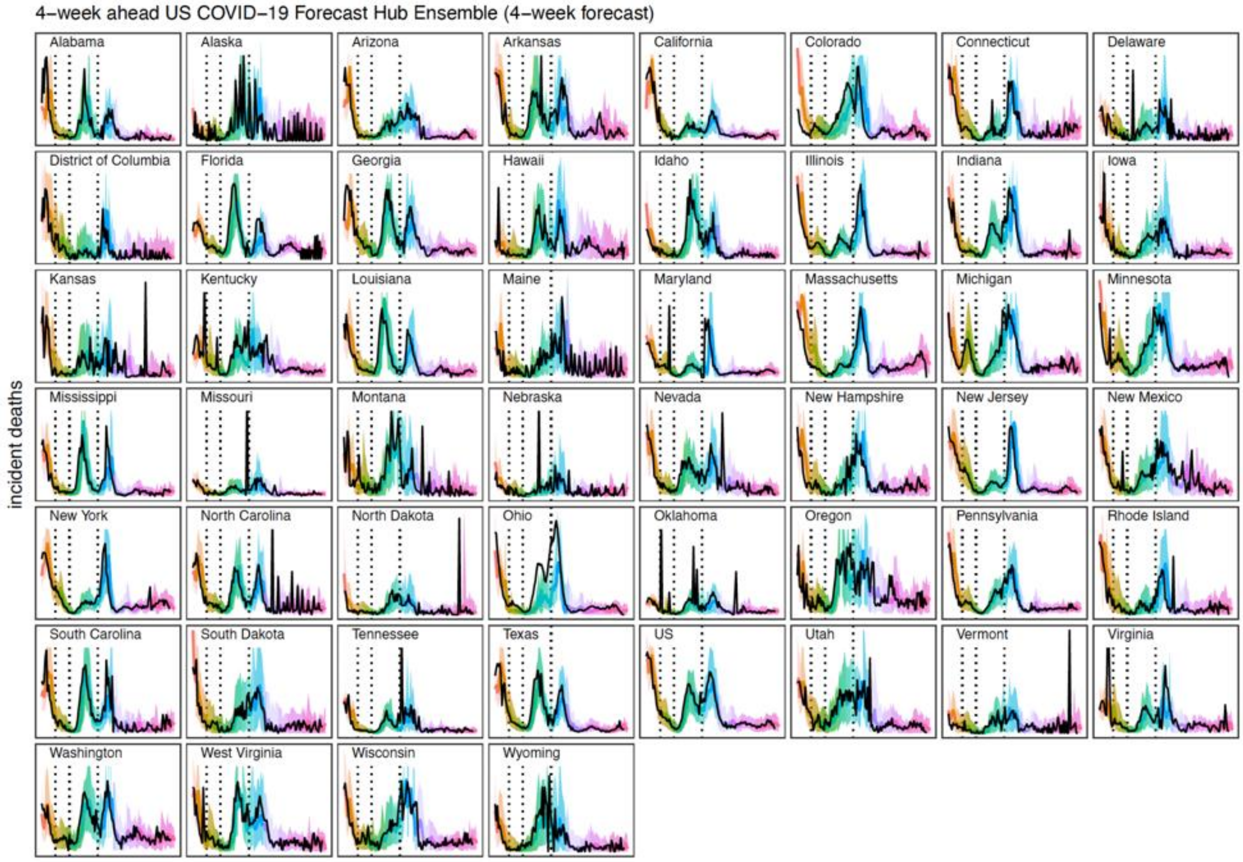


**Figure S9: 4-week forecast comparative model projections for incident deaths.** Across locations, median (line) and 95% projection interval (ribbon) for each round is shown with a different color. Black curve represents observations, and dotted black vertical lines represent the truncation dates for Alpha (2021-04-03), Delta (2021-06-26), and Omicron (2021-12-25).

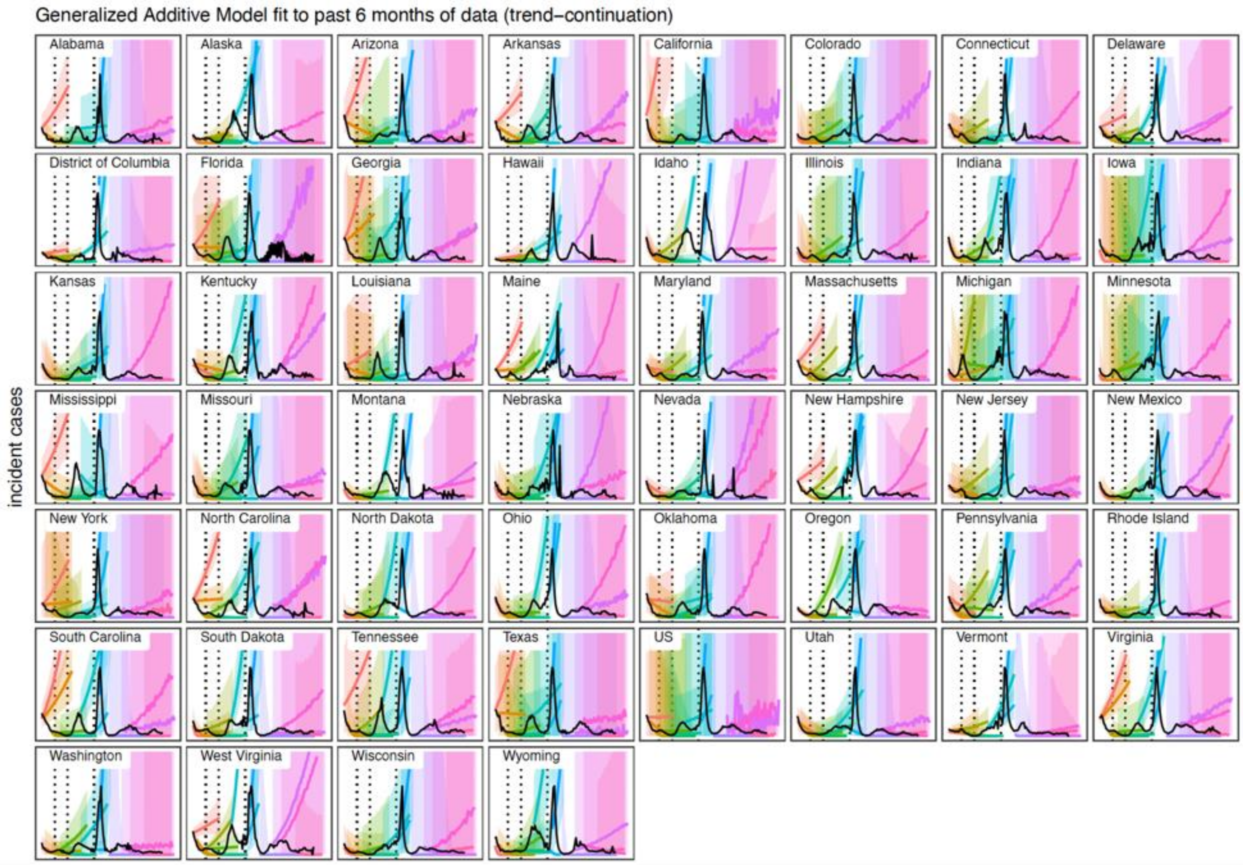


**Figure S10: Trend-continuation comparative model projections for incident cases.** Across locations, median (line) and 95% projection interval (ribbon) for each round is shown with a different color. Black curve represents observations, and dotted black vertical lines represent the truncation dates for Alpha (2021-04-03), Delta (2021-06-26), and Omicron (2021-12-25).

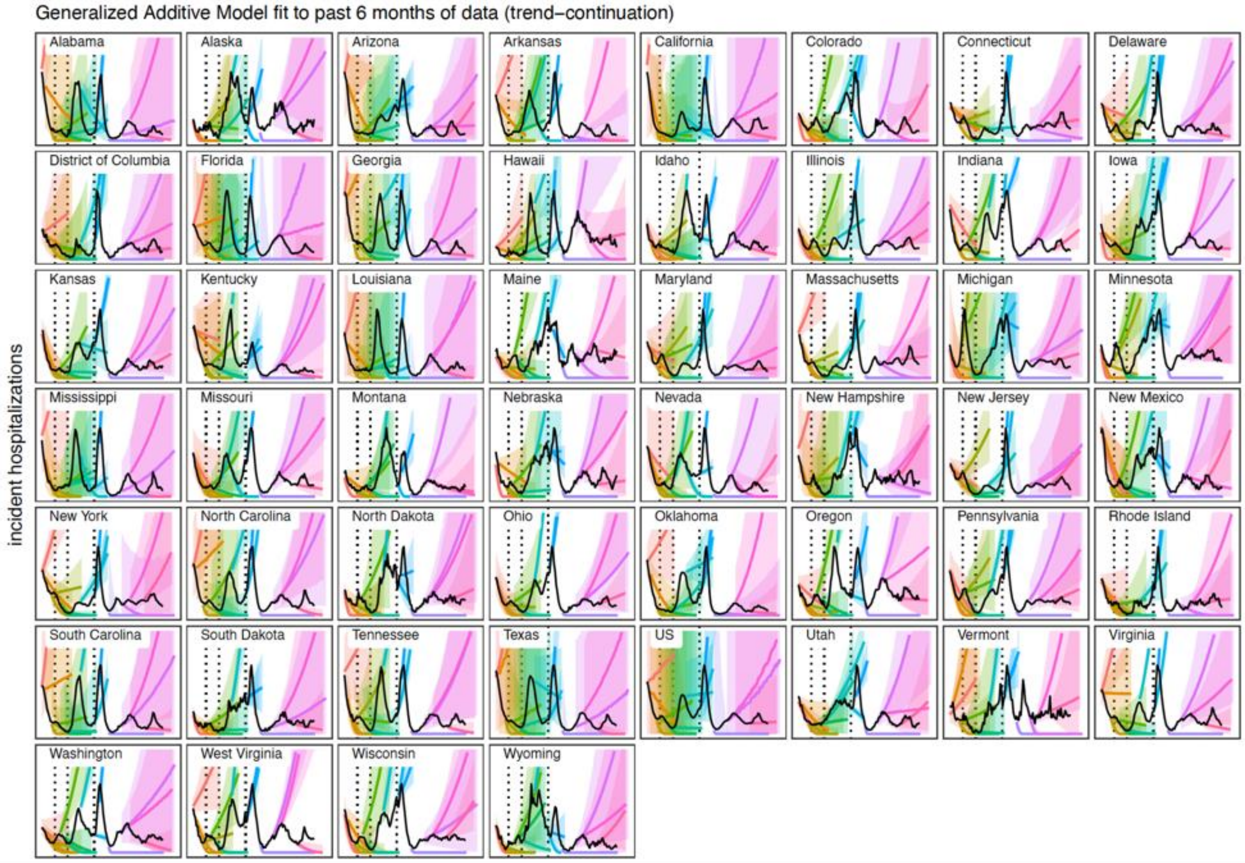


**Figure S11: Trend-continuation comparative model projections for incident hospitalizations.** Across locations, median (line) and 95% projection interval (ribbon) for each round is shown with a different color. Black curve represents observations, and dotted black vertical lines represent the truncation dates for Alpha (2021-04-03), Delta (2021-06-26), and Omicron (2021-12-25).

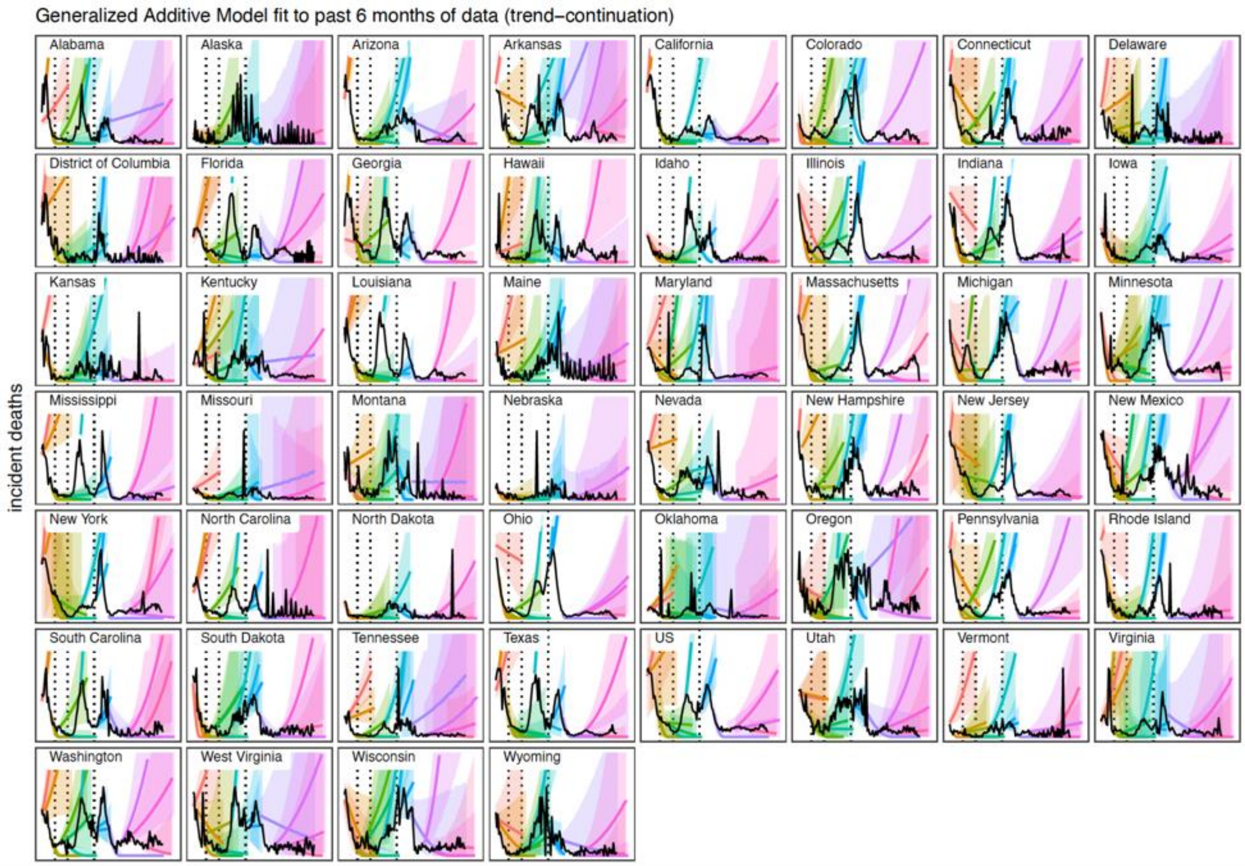


**Figure S12: Trend-continuation comparative model projections for incident deaths.** Across locations, median (line) and 95% projection interval (ribbon) for each round is shown with a different color. Black curve represents observations, and dotted black vertical lines represent the truncation dates for Alpha (2021-04-03), Delta (2021-06-26), and Omicron (2021-12-25).

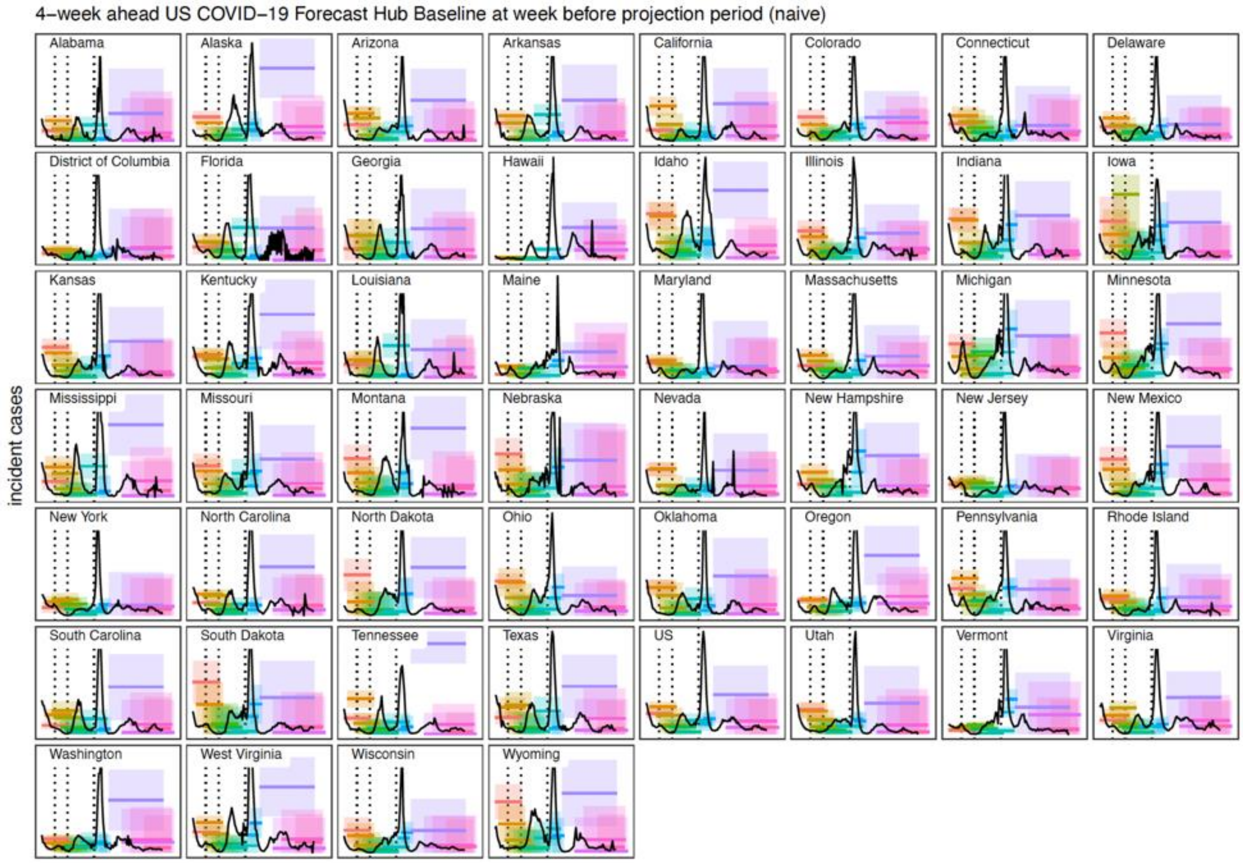


**Figure S13: Naive comparative model projections for incident cases.** Across locations, median (line) and 95% projection interval (ribbon) for each round is shown with a different color. Black curve represents observations, and dotted black vertical lines represent the truncation dates for Alpha (2021-04-03), Delta (2021-06-26), and Omicron (2021-12-25).

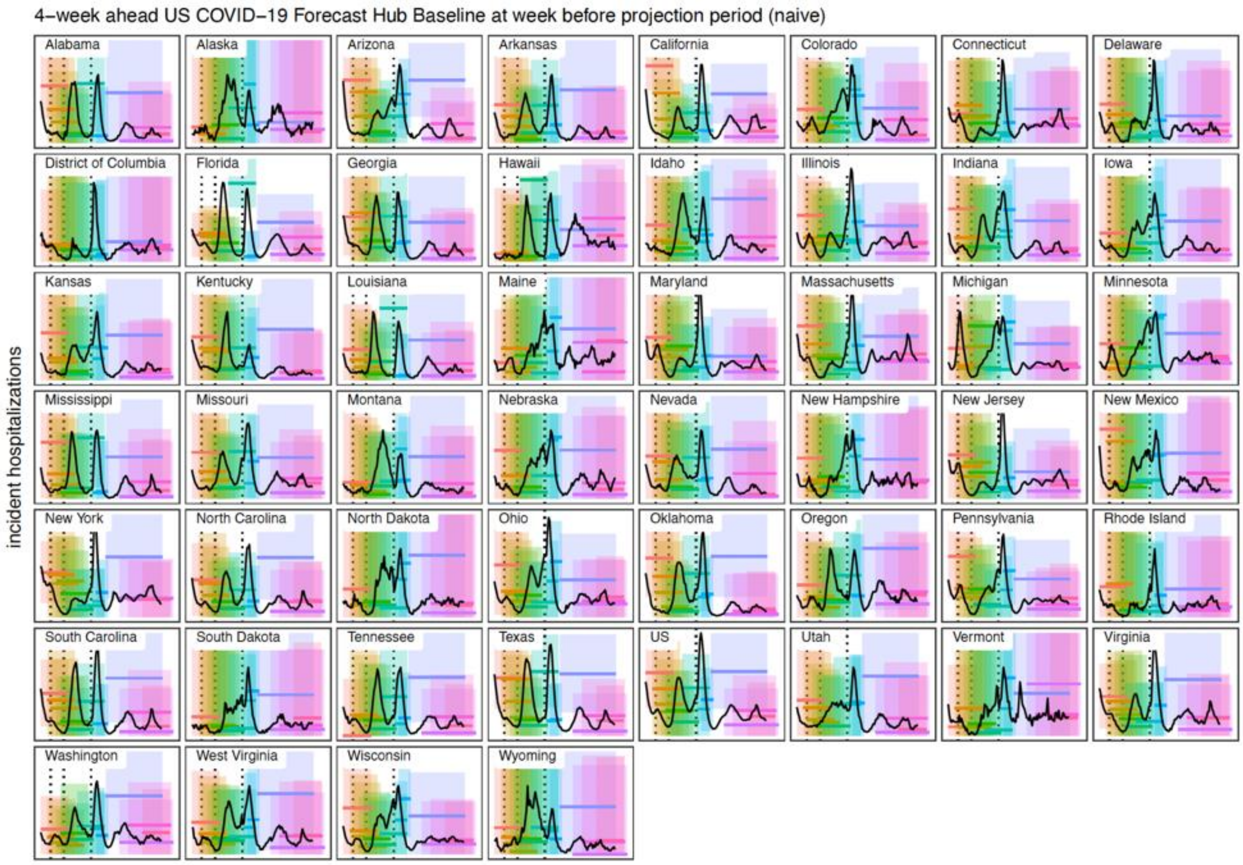


**Figure S14: Naive comparative model projections for incident hospitalizations.** Across locations, median (line) and 95% projection interval (ribbon) for each round is shown with a different color. Black curve represents observations, and dotted black vertical lines represent the truncation dates for Alpha (2021-04-03), Delta (2021-06-26), and Omicron (2021-12-25).

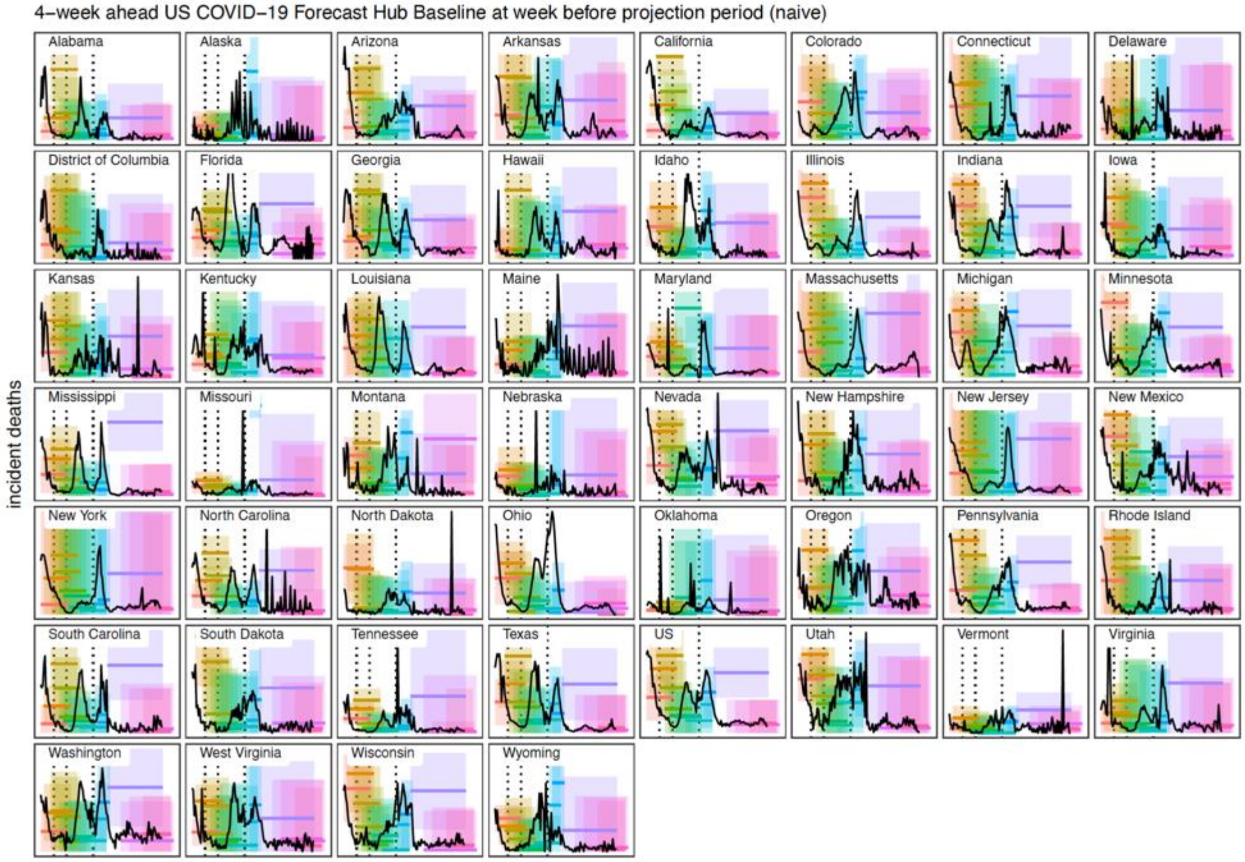


**Figure S15: Naive comparative model projections for incident deaths.** Across locations, median (line) and 95% projection interval (ribbon) for each round is shown with a different color. Black curve represents observations, and dotted black vertical lines represent the truncation dates for Alpha (2021-04-03), Delta (2021-06-26), and Omicron (2021-12-25).

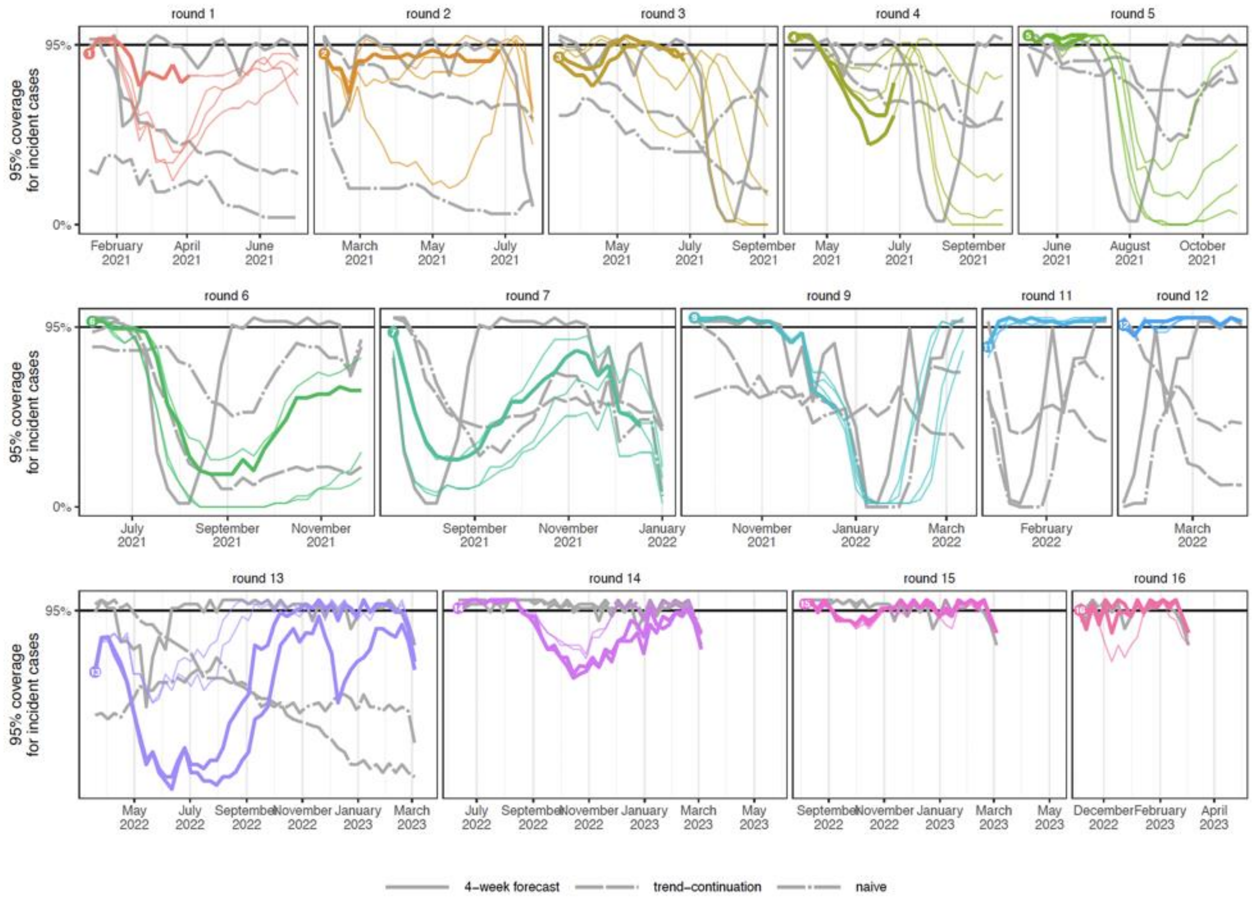


**Figure S16: Ninety-five percent (95%) prediction interval coverage of SMH ensemble and comparative models for incident cases.** Across all rounds, coverage is shown for SMH trimmed-LOP ensemble (color), with the most realistic scenario bolded. Coverage is compared to a highly-informed model (solid gray line), a trend-continuation model (dashed gray line), and a naive model (dot-dashed gray line). Ideal coverage of 95% is indicated by a black horizontal line. Each round is shown in a different panel, with only the projection period for that round included. Ground truth data was only produced through 4 March 2023, so our evaluation of Rounds 13-16 ends at this date. Note, jagged coverage in Rounds 13-16 is due to increased number of reporting anomalies in later weeks (e.g., Florida).

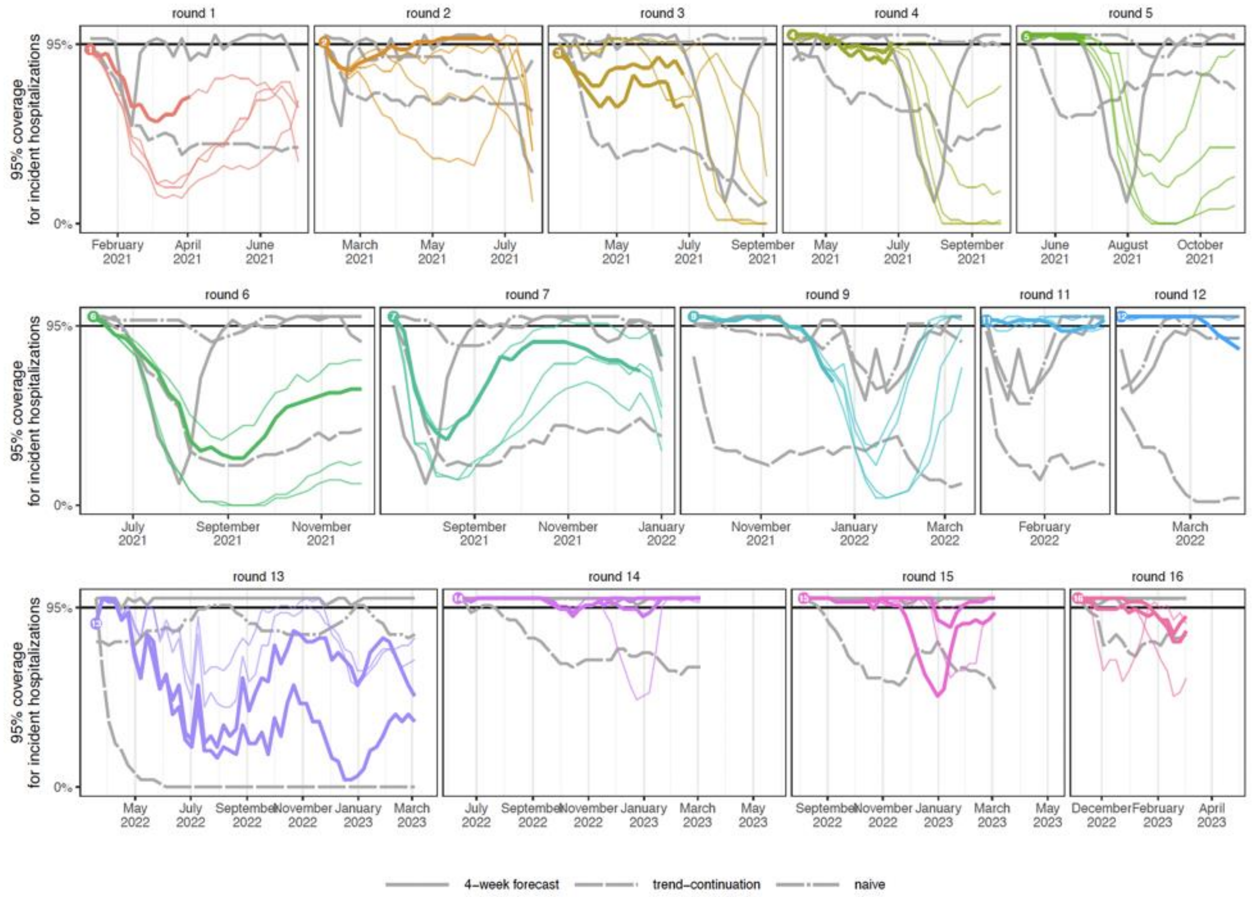


**Figure S17: Ninety-five percent (95%) prediction interval coverage of SMH ensemble and comparative models for incident hospitalizations.** Across all rounds, coverage is shown for SMH trimmed-LOP ensemble (color), with the most realistic scenario bolded. Coverage is compared to a 4-week forecast model (solid gray line), a trend-continuation model (dashed gray line), and a naive model (dot-dashed gray line). Ideal coverage of 95% is indicated by a black horizontal line. Each round is shown in a different panel, with only the projection period for that round included. Ground truth data was only produced through 4 March 2023, so our evaluation of Rounds 13-16 ends at this date.

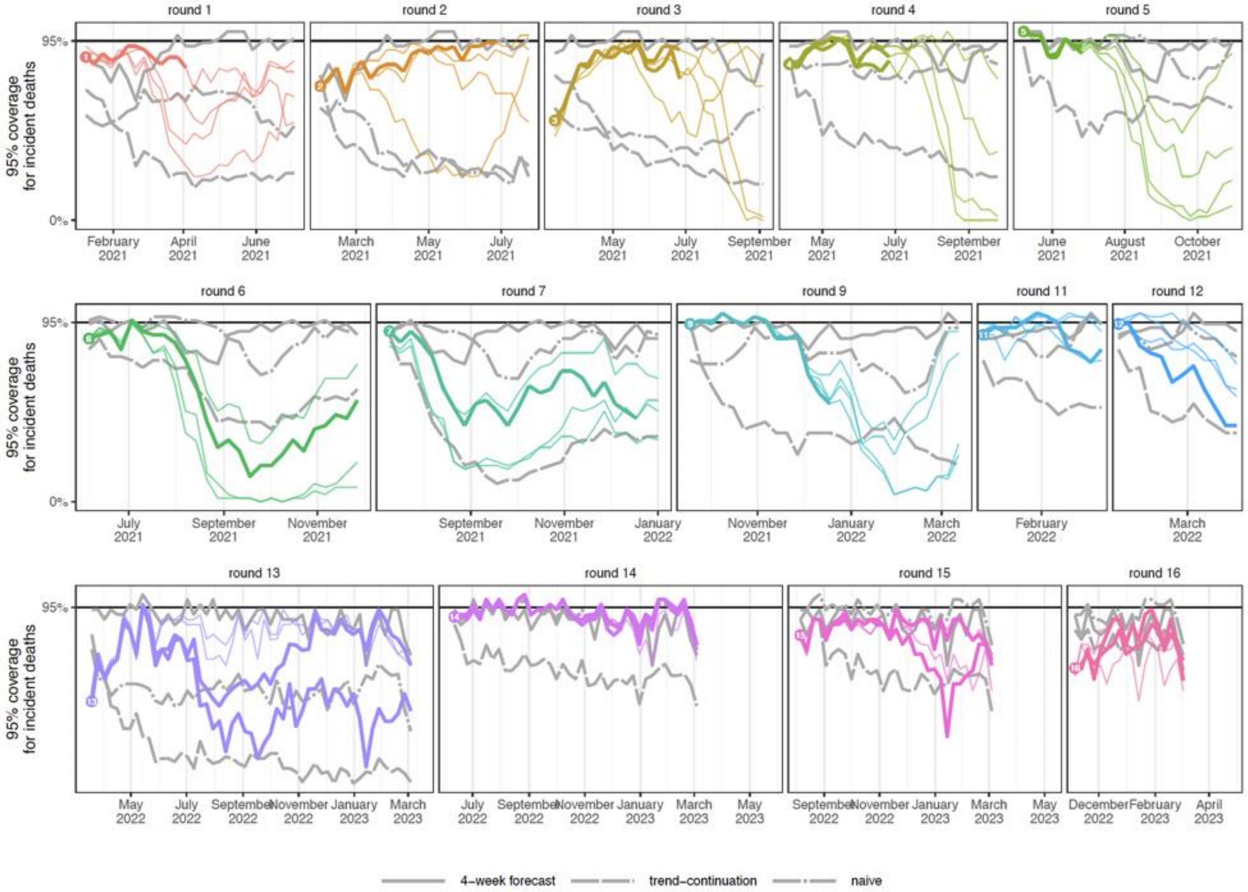


**Figure S18: Ninety-five percent (95%) prediction interval coverage of SMH ensemble and comparative models for incident deaths.** Across all rounds, coverage is shown for SMH trimmed-LOP ensemble (color), with the most realistic scenario bolded. Coverage is compared to a highly-informed model (solid gray line), a trend-continuation model (dashed gray line), and a naive model (dot-dashed gray line). Ideal coverage of 95% is indicated by a black horizontal line. Each round is shown in a different panel, with only the projection period for that round included. Ground truth data was only produced through 4 March 2023, so our evaluation of Rounds 13-16 ends at this date. Note, jagged coverage in Rounds 13-16 is due to increased number of reporting anomalies in later weeks (e.g., Florida).

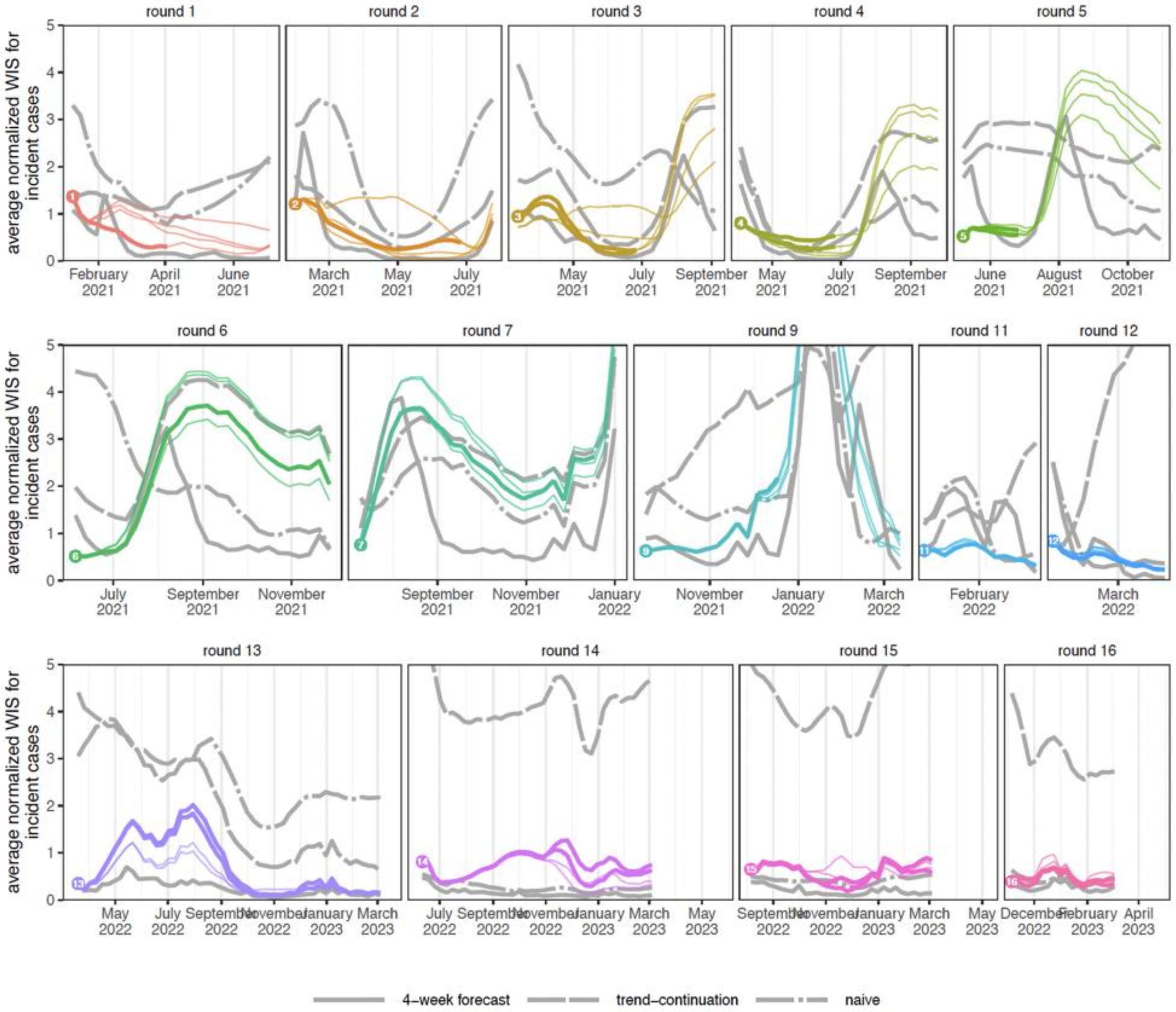


**Figure S19: Average normalized WIS of SMH ensemble and comparative models for incident cases.** Across all rounds, average normalized WIS is shown for SMH trimmed-LOP ensemble (color), with the most realistic scenario bolded. Average normalized WIS is compared to a 4-week forecast model (solid gray line), a trend-continuation model (dashed gray line), and a naive model (dot-dashed gray line). Each round is shown in a different panel, with only the projection period for that round included. Normalized WIS is calculated by relativizing WIS by the standard deviation of WIS across all scenarios and models for a given week, target, and round. This yields a scale-free value, and we average normalized WIS across all locations for a given projection week (and scenario in the case of the SMH ensemble). Ground truth data was only produced through 4 March 2023, so our evaluation of Rounds 13-16 ends at this date.

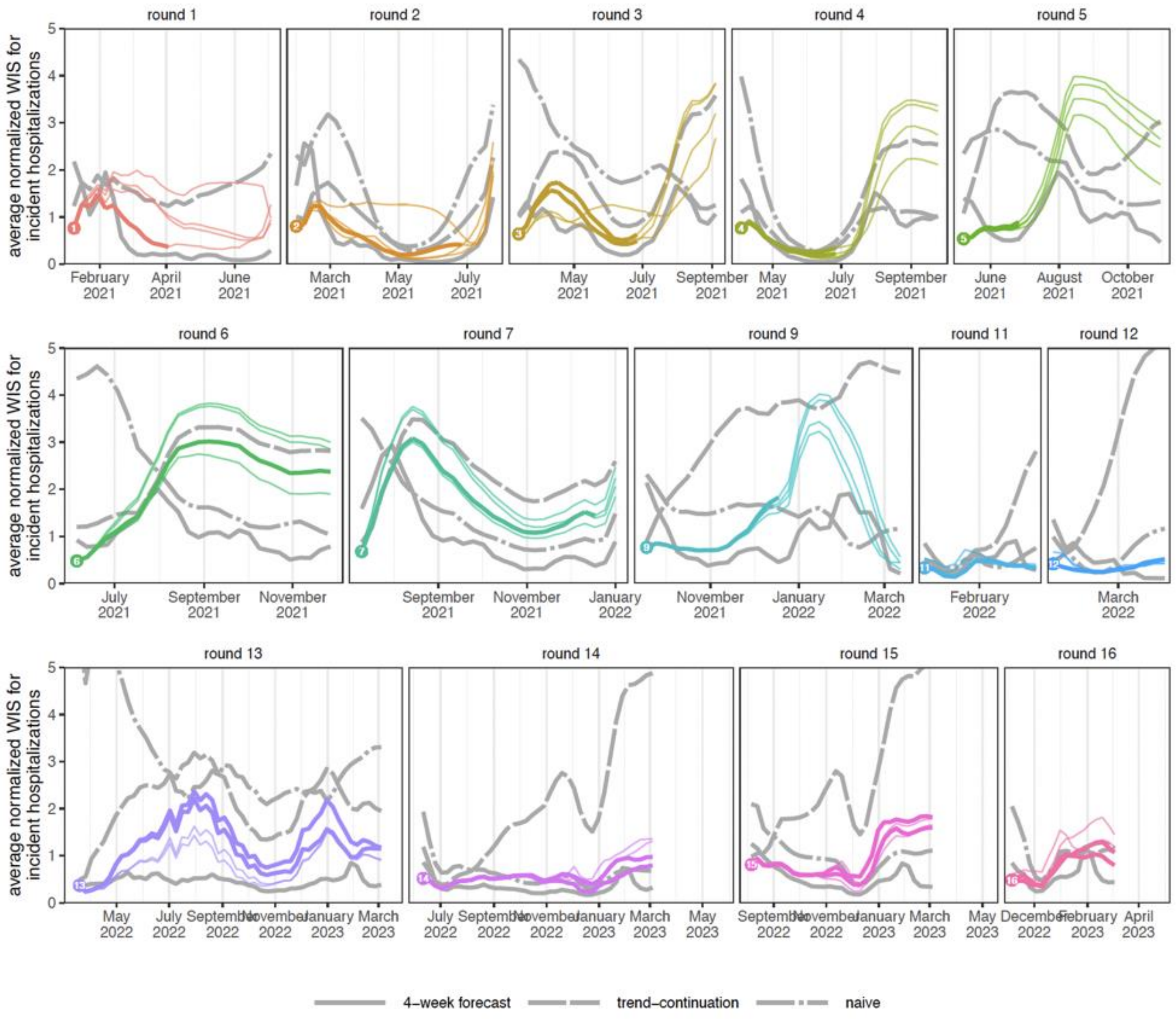


**Figure S20: Average normalized WIS of SMH ensemble and comparative models for incident hospitalizations.** Across all rounds, average normalized WIS is shown for SMH trimmed-LOP ensemble (color), with the most realistic scenario bolded. Average normalized WIS is compared to a highly-informed model (solid gray line), a trend-continuation model (dashed gray line), and a naive model (dot-dashed gray line). Each round is shown in a different panel, with only the projection period for that round included. Normalized WIS is calculated by relativizing WIS by the standard deviation of WIS across all scenarios and models for a given week, target, and round. This yields a scale-free value, and we average normalized WIS across all locations for a given projection week (and scenario in the case of the SMH ensemble). Ground truth data was only produced through 4 March 2023, so our evaluation of Rounds 13-16 ends at this date.

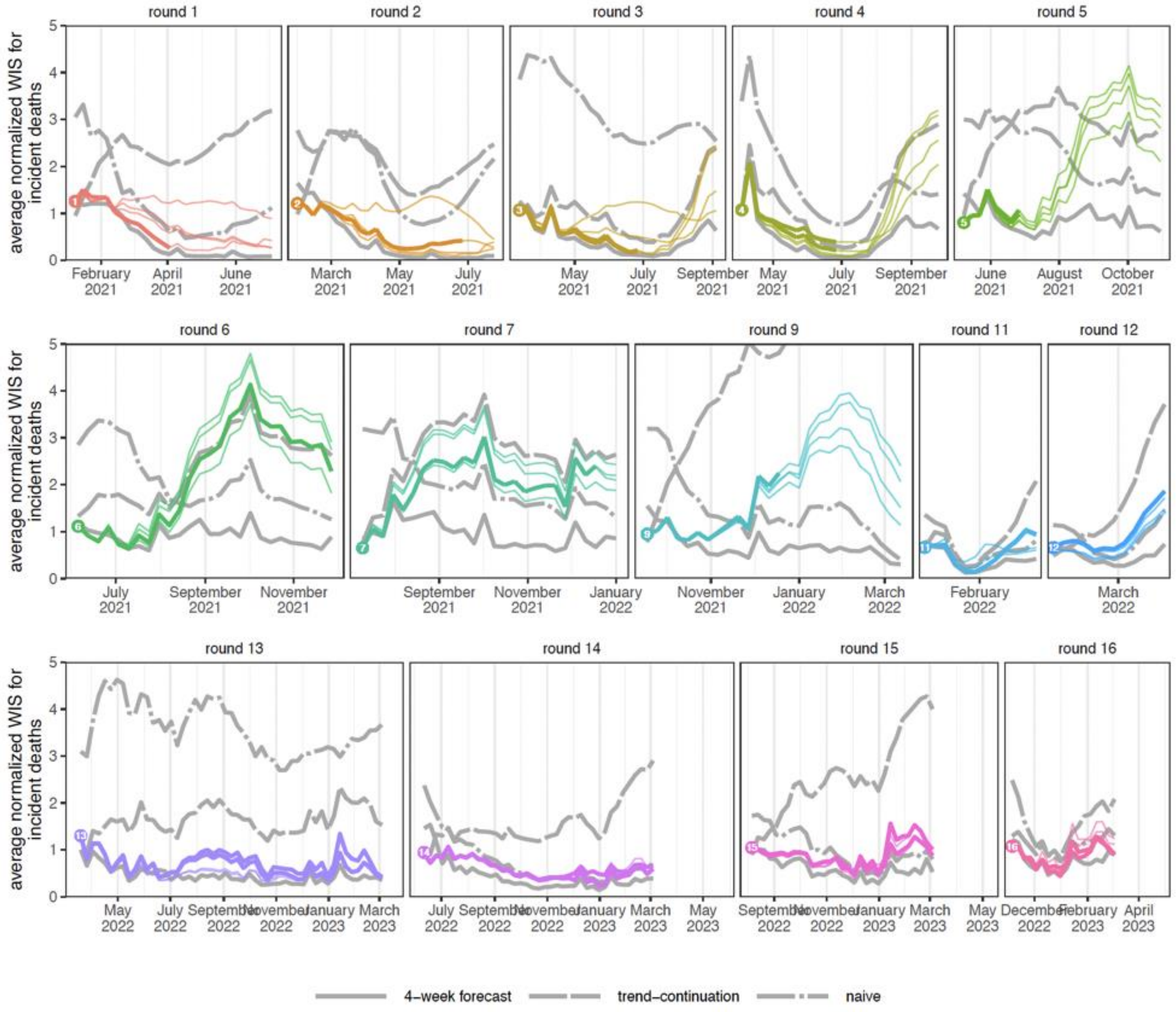


**Figure S21: Average normalized WIS of SMH ensemble and comparative models for incident deaths.** Across all rounds, average normalized WIS is shown for SMH trimmed-LOP ensemble (color), with the most realistic scenario bolded. Average normalized WIS is compared to a 4-week forecast model (solid gray line), a trend-continuation model (dashed gray line), and a naive model (dot-dashed gray line). Each round is shown in a different panel, with only the projection period for that round included. Normalized WIS is calculated by relativizing WIS by the standard deviation of WIS across all scenarios and models for a given week, target, and round. This yields a scale-free value, and we average normalized WIS across all locations for a given projection week (and scenario in the case of the SMH ensemble). Ground truth data was only produced through 4 March 2023, so our evaluation of Rounds 13-16 ends at this date.

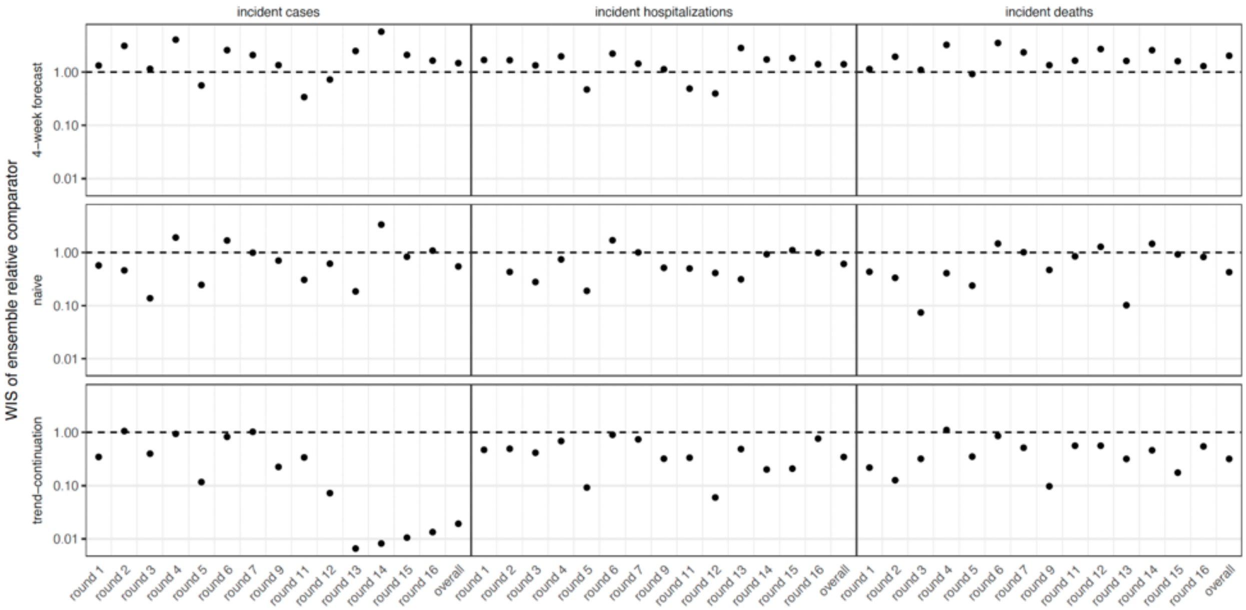


**Figure S22: WIS ratio of SMH ensemble compared to each comparative model for incident cases, hospitalizations, and deaths.** WIS for SMH ensemble and each comparative model is averaged across all locations and weeks in a given round (considering only realistic scenarios), or across all locations, weeks and rounds, for "overall". WIS ratio is the average WIS for the SMH ensemble divided by that for each comparative model. Values below 1 (dashed horizontal line) indicates that SMH ensemble performance is better than that of the comparative model.

### S4. Trend classification

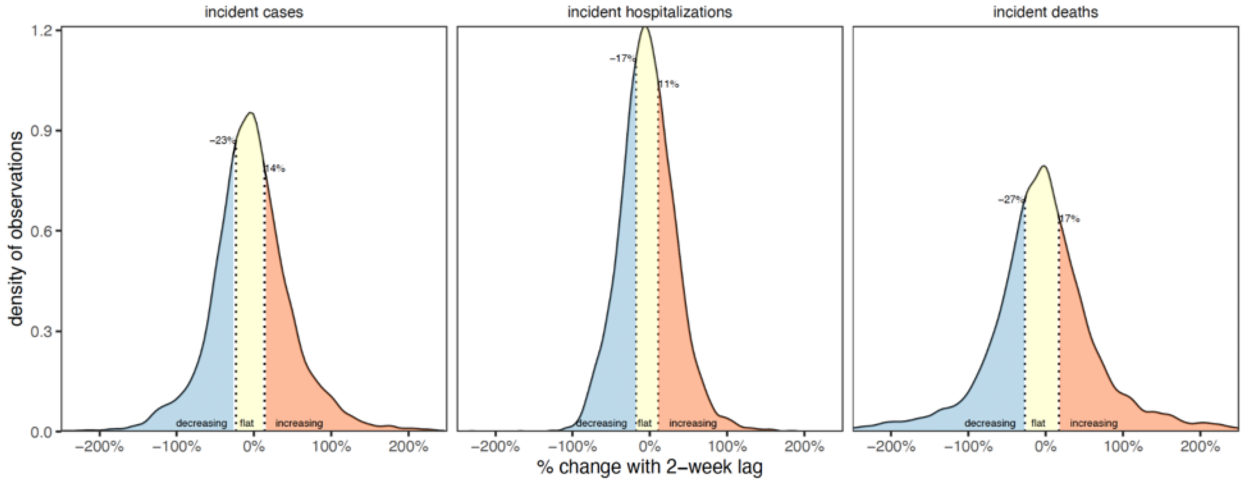


**Figure S23: Distribution of percent change values (logarithmic scale) for 2-week lag and 33% of observations flat.** For a given location, we calculated the percentage change over a two-week interval, i.e., % change = $\log(o_t)$ - $\log(o_{t-2})$. From the distribution of percent change values across all targets, locations, and weeks (26 December 2020 - 4 March 2023), we identify the upper and lower threshold for classifying a time-point as decreasing (blue, below Q33) or increasing (orange, above Q66). The central values are classified as flat (yellow, between Q33 and Q66). The x-axis has been truncated to -250%-250%, though > 0.1% of % change values are outside of this interval.

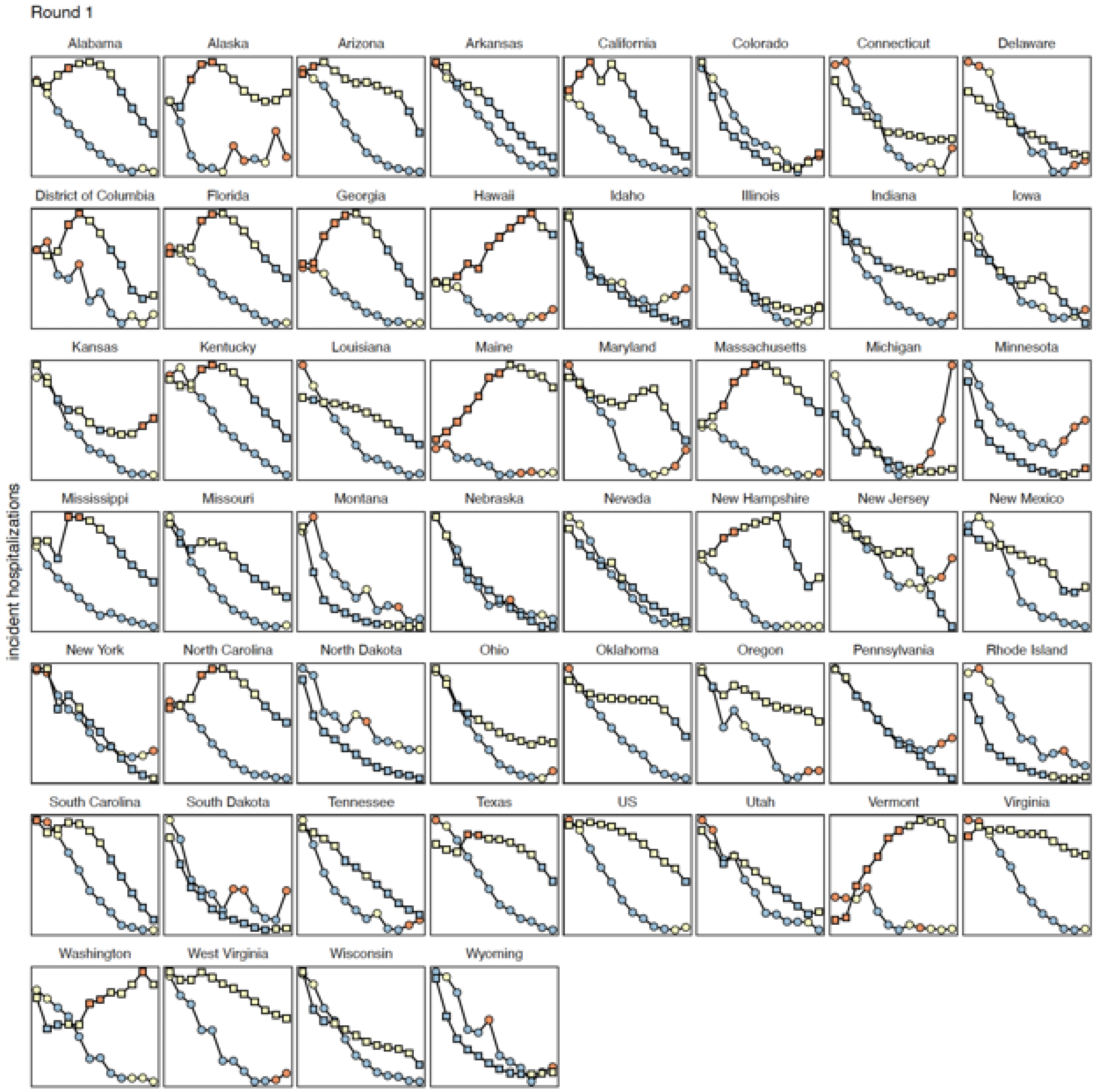


**Figure S24: Classifications of observations and projections as decreasing (blue), flat (yellow), or increasing (orange) for Round 1.** Observed (circles) and projected (squares, SMH ensemble median) incident hospitalizations are shown from 26 Dec 2020 through 27 Mar 2021. Observed incident hospitalizations in the two weeks prior to the start of the projection period (gray) are used to classify the first two weeks and shown for reference. For simplicity, we show the most plausible scenario over the plausible time window (i.e., after truncation). Note, the scale of the y-axis is not consistent across panels.

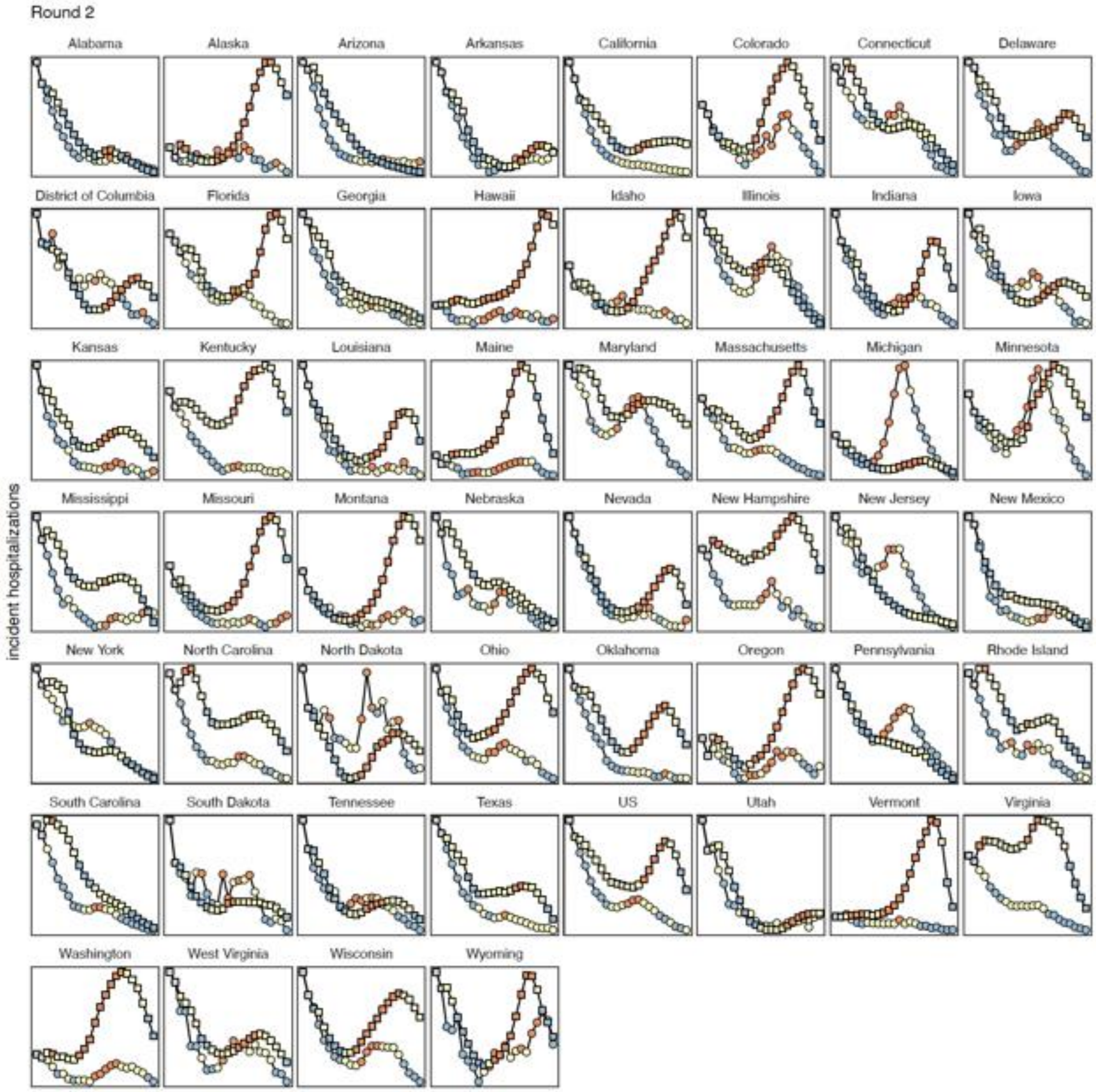


**Figure S25: Classifications of observations and projections as decreasing (blue), flat (yellow), or increasing (orange) for Round 2.** Observed (circles) and projected (squares, SMH ensemble median) incident hospitalizations are shown from 16 Jan 2021 through 19 Jun 2021. Observed incident hospitalizations in the two weeks prior to the start of the projection period (gray) are used to classify the first two weeks and shown for reference. For simplicity, we show the most plausible scenario over the plausible time window (i.e., after truncation). Note, the scale of the y-axis is not consistent across panels.

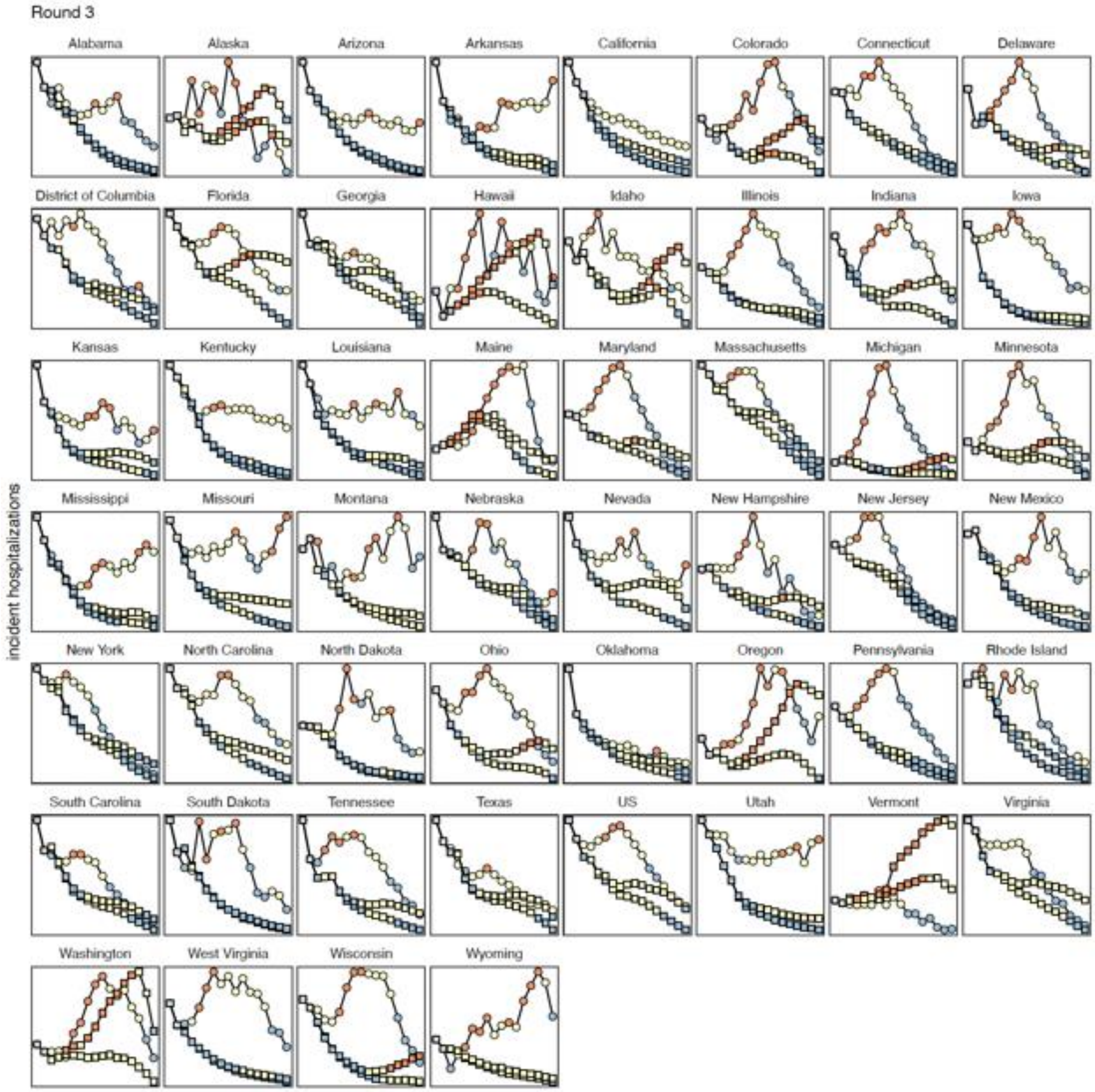


**Figure S26: Classifications of observations and projections as decreasing (blue), flat (yellow), or increasing (orange) for Round 3.** Observed (circles) and projected (squares, SMH ensemble median) incident hospitalizations are shown from 27 Feb 2021 through 19 Jun 2021. Observed incident hospitalizations in the two weeks prior to the start of the projection period (gray) are used to classify the first two weeks and shown for reference. For simplicity, we show the two most plausible scenarios for this round over the plausible time window (i.e., after truncation). Note, the scale of the y-axis is not consistent across panels.

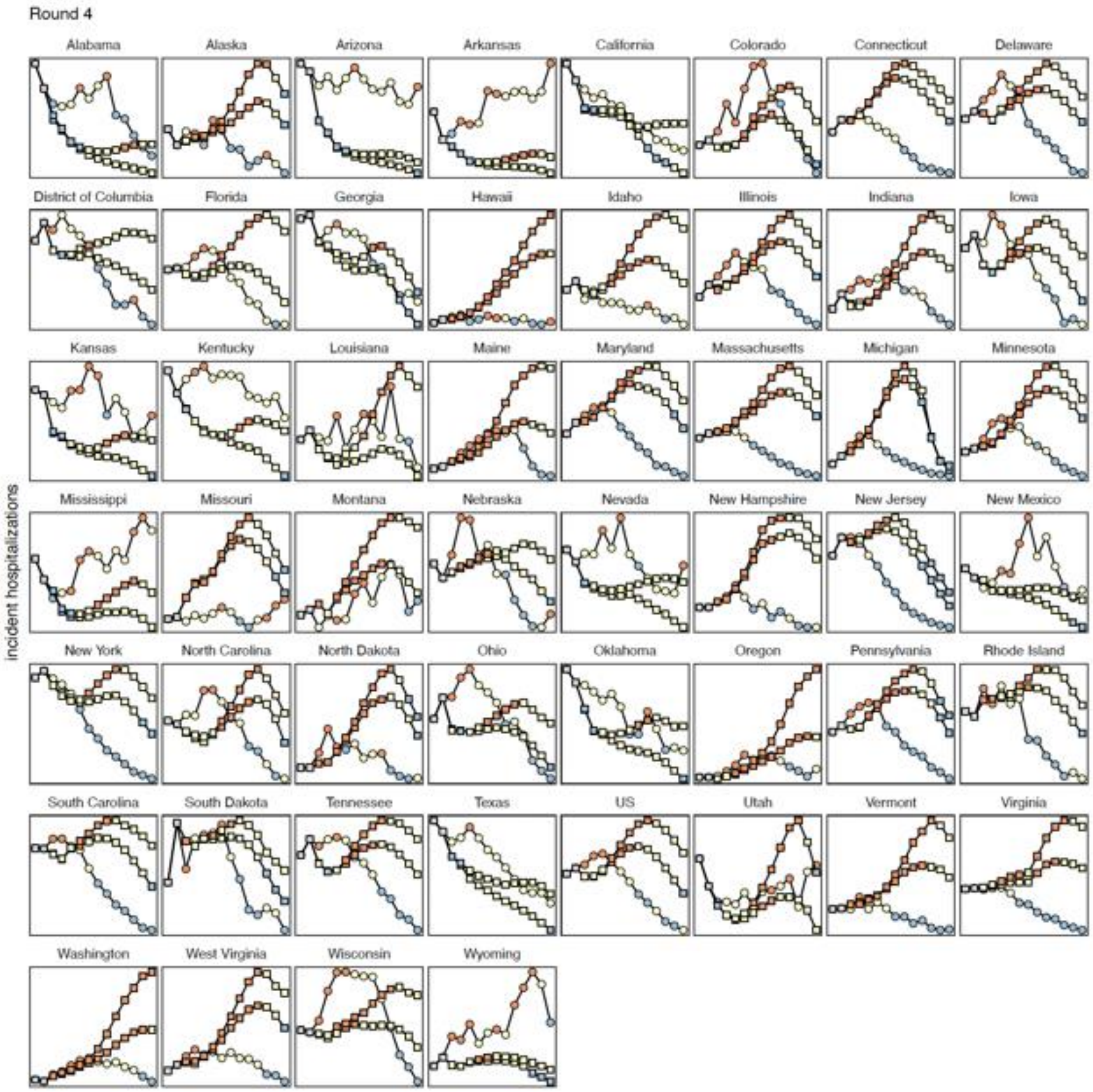


**Figure S27: Classifications of observations and projections as decreasing (blue), flat (yellow), or increasing (orange) for Round 4.** Observed (circles) and projected (squares, SMH ensemble median) incident hospitalizations are shown from 20 Mar 2021 through 19 Jun 2021. Observed incident hospitalizations in the two weeks prior to the start of the projection period (gray) are used to classify the first two weeks and shown for reference. For simplicity, we show the two most plausible scenarios for this round over the plausible time window (i.e., after truncation). Note, the scale of the y-axis is not consistent across panels.

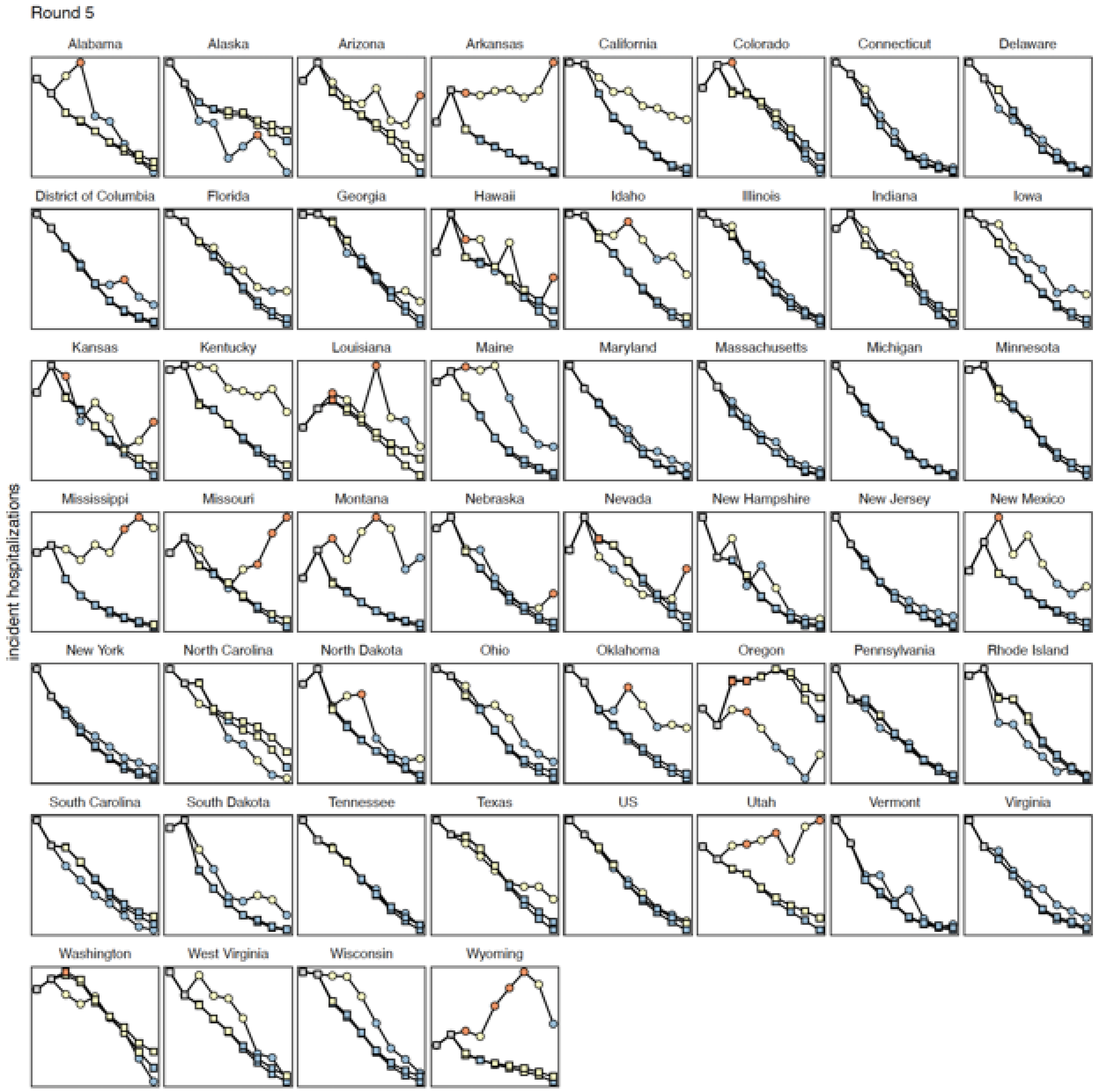


**Figure S28: Classifications of observations and projections as decreasing (blue), flat (yellow), or increasing (orange) for Round 5.** Observed (circles) and projected (squares, SMH ensemble median) incident hospitalizations are shown from 24 Arp 2021 through 19 Jun 2021. Observed incident hospitalizations in the two weeks prior to the start of the projection period (gray) are used to classify the first two weeks and shown for reference. For simplicity, we show the most plausible scenario over the plausible time window (i.e., after truncation). Note, the scale of the y-axis is not consistent across panels.

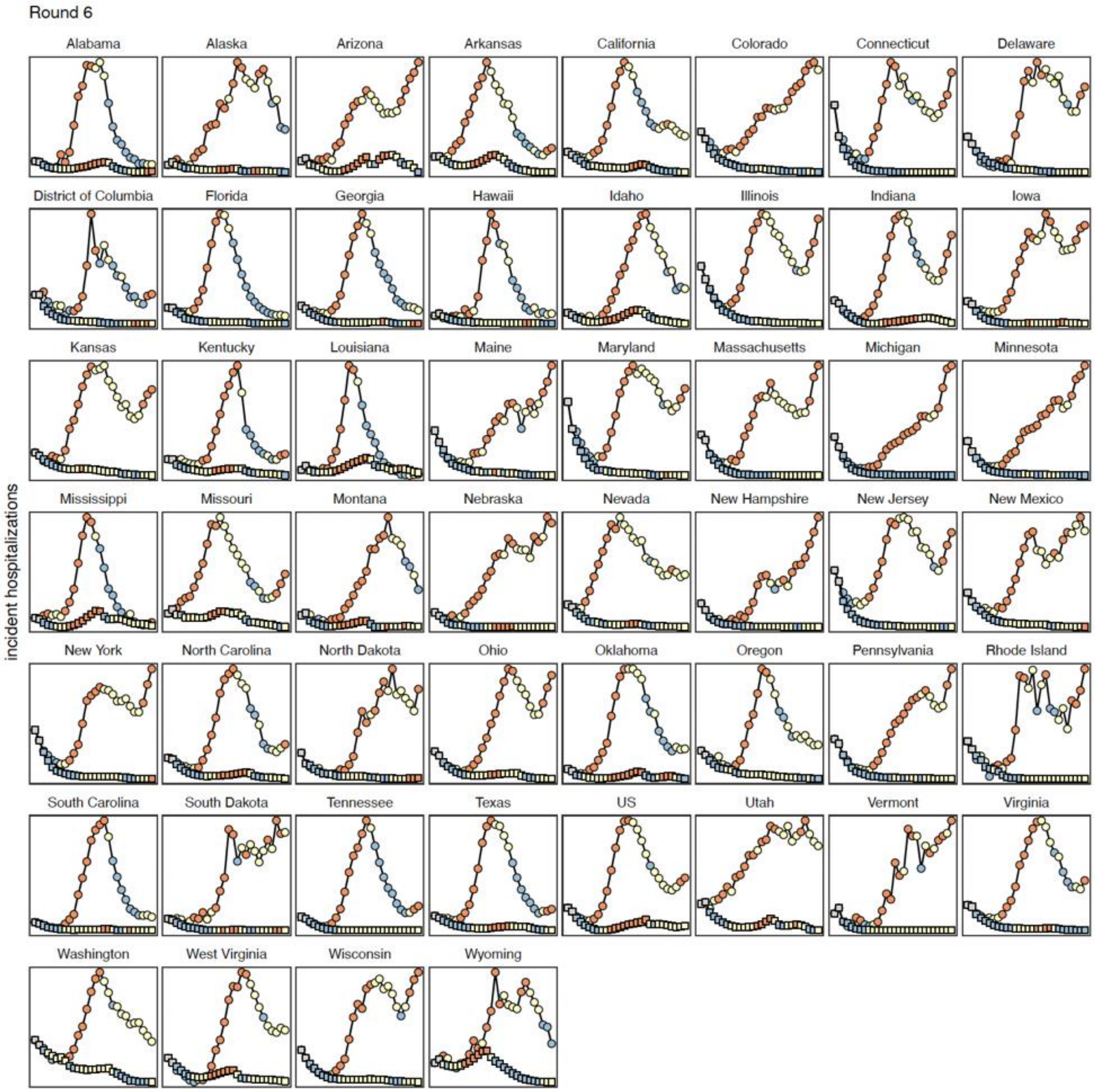


**Figure S29: Classifications of observations and projections as decreasing (blue), flat (yellow), or increasing (orange) for Round 6.** Observed (circles) and projected (squares, SMH ensemble median) incident hospitalizations are shown from 22 May 2021 through 27 Nov 2021. Observed incident hospitalizations in the two weeks prior to the start of the projection period (gray) are used to classify the first two weeks and shown for reference. For simplicity, we show the most plausible scenario over the plausible time window (i.e., after truncation). Note, the scale of the y-axis is not consistent across panels.

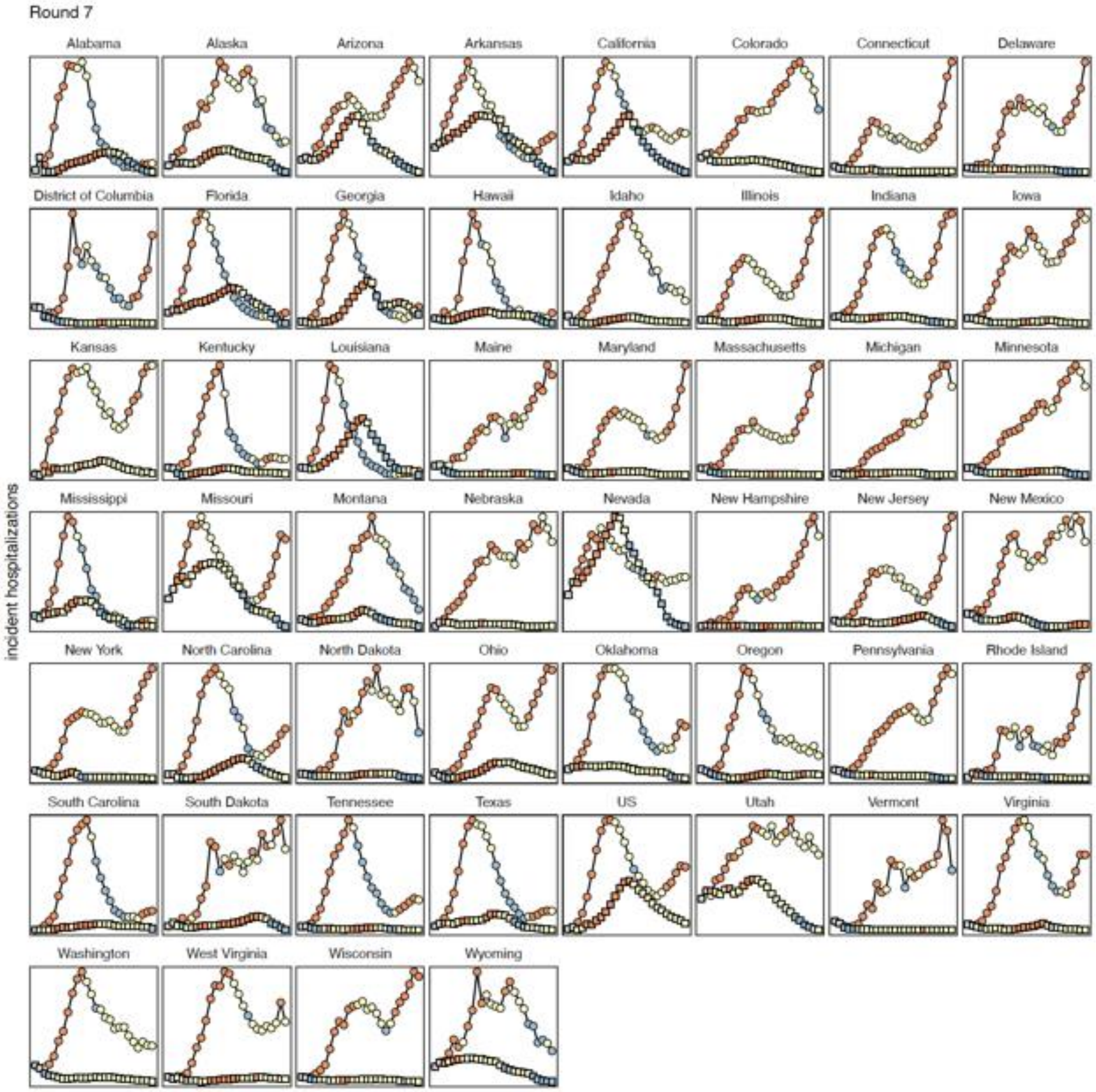


**Figure S30: Classifications of observations and projections as decreasing (blue), flat (yellow), or increasing (orange) for Round 7.** Observed (circles) and projected (squares, SMH ensemble median) incident hospitalizations are shown from 26 Jun 2021 through 18 Dec 2021. Observed incident hospitalizations in the two weeks prior to the start of the projection period (gray) are used to classify the first two weeks and shown for reference. For simplicity, we show the most plausible scenario over the plausible time window (i.e., after truncation). Note, the scale of the y-axis is not consistent across panels.

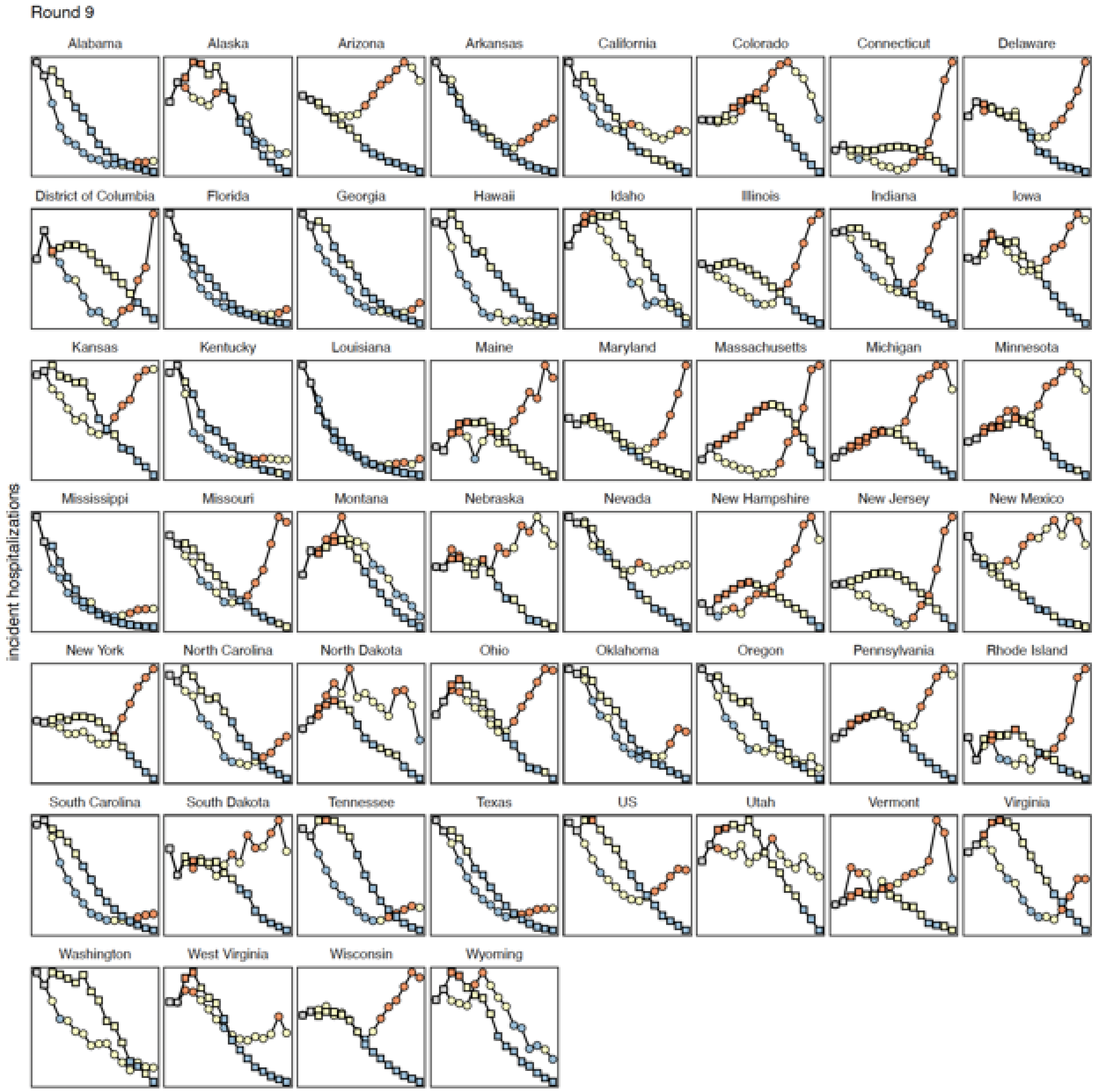


**Figure S31: Classifications of observations and projections as decreasing (blue), flat (yellow), or increasing (orange) for Round 9.** Observed (circles) and projected (squares, SMH ensemble median) incident hospitalizations are shown from 4 Sept 2021 through 18 Dec 2021. Observed incident hospitalizations in the two weeks prior to the start of the projection period (gray) are used to classify the first two weeks and shown for reference. For simplicity, we show the most plausible scenario over the plausible time window (i.e., after truncation). Note, the scale of the y-axis is not consistent across panels.

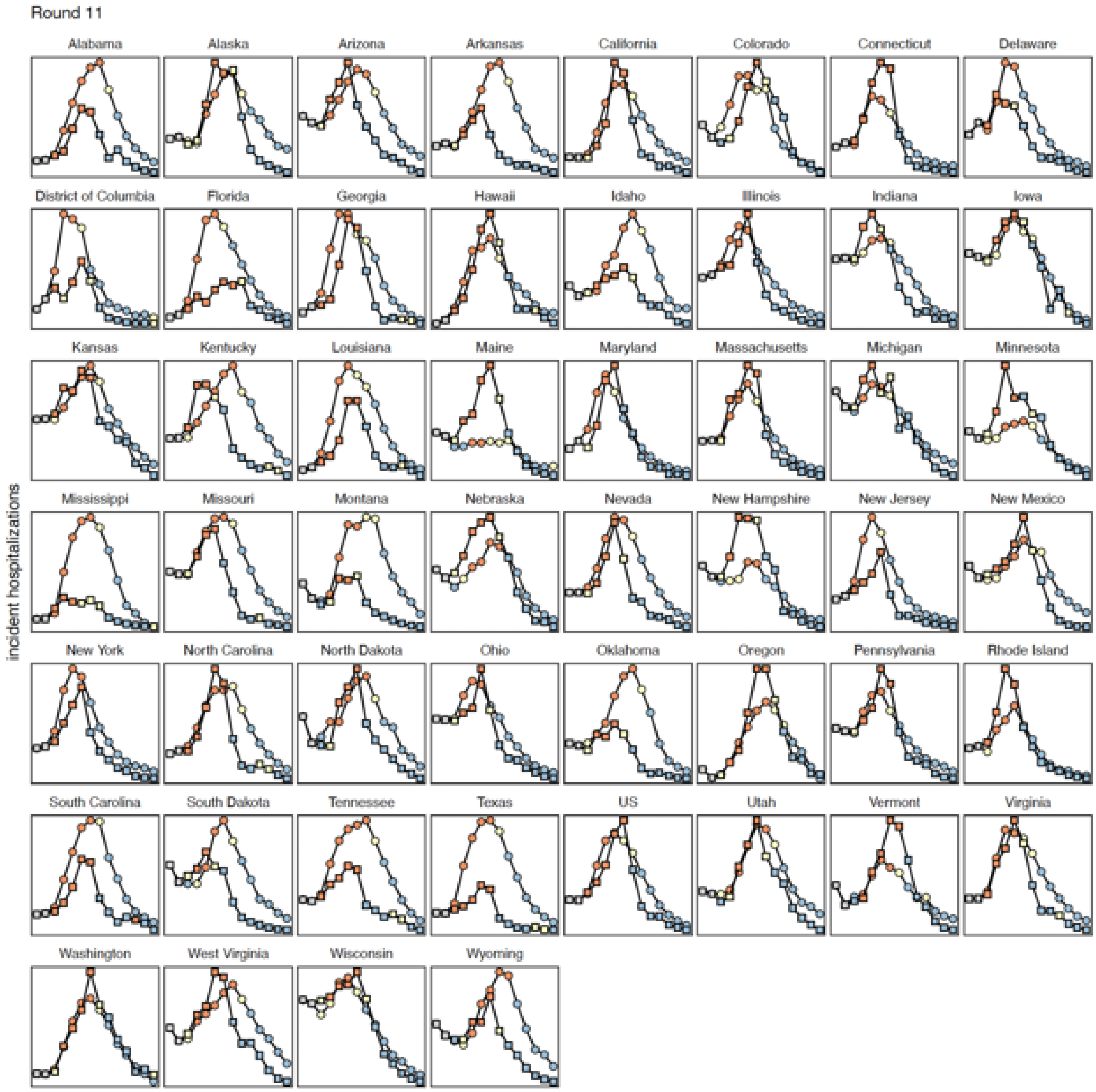


**Figure S32: Classifications of observations and projections as decreasing (blue), flat (yellow), or increasing (orange) for Round 11.** Observed (circles) and projected (squares, SMH ensemble median) incident hospitalizations are shown from 11 Dec 2021 through 12 Mar 2022. Observed incident hospitalizations in the two weeks prior to the start of the projection period (gray) are used to classify the first two weeks and shown for reference. For simplicity, we show the most plausible scenario over the plausible time window (i.e., after truncation). Note, the scale of the y-axis is not consistent across panels.

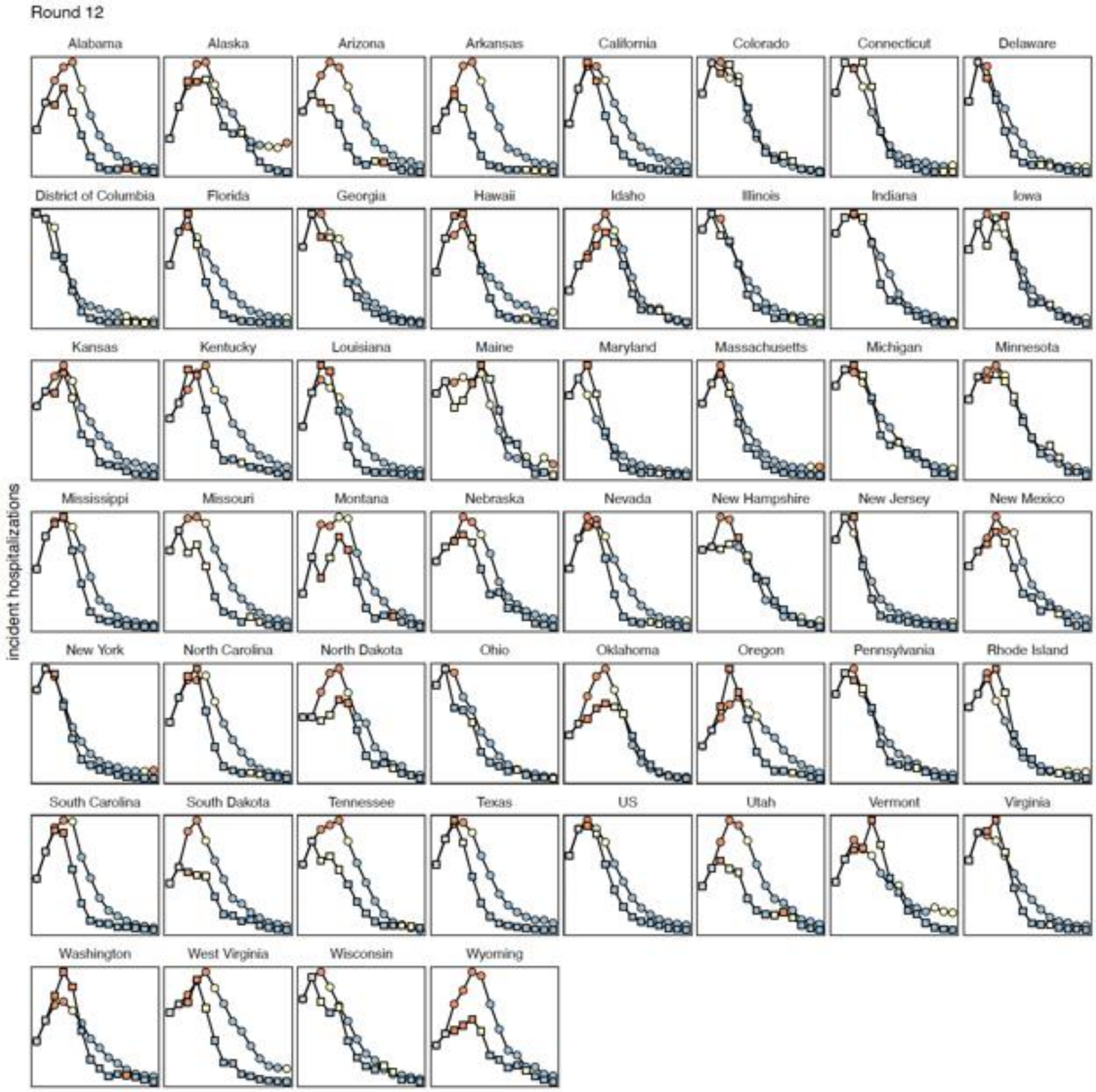


**Figure S33: Classifications of observations and projections as decreasing (blue), flat (yellow), or increasing (orange) for Round 12.** Observed (circles) and projected (squares, SMH ensemble median) incident hospitalizations are shown from 1 Jan 2022 through 2 Apr 2022. Observed incident hospitalizations in the two weeks prior to the start of the projection period (gray) are used to classify the first two weeks and shown for reference. For simplicity, we show the most plausible scenario over the plausible time window (i.e., after truncation). Note, the scale of the y-axis is not consistent across panels.

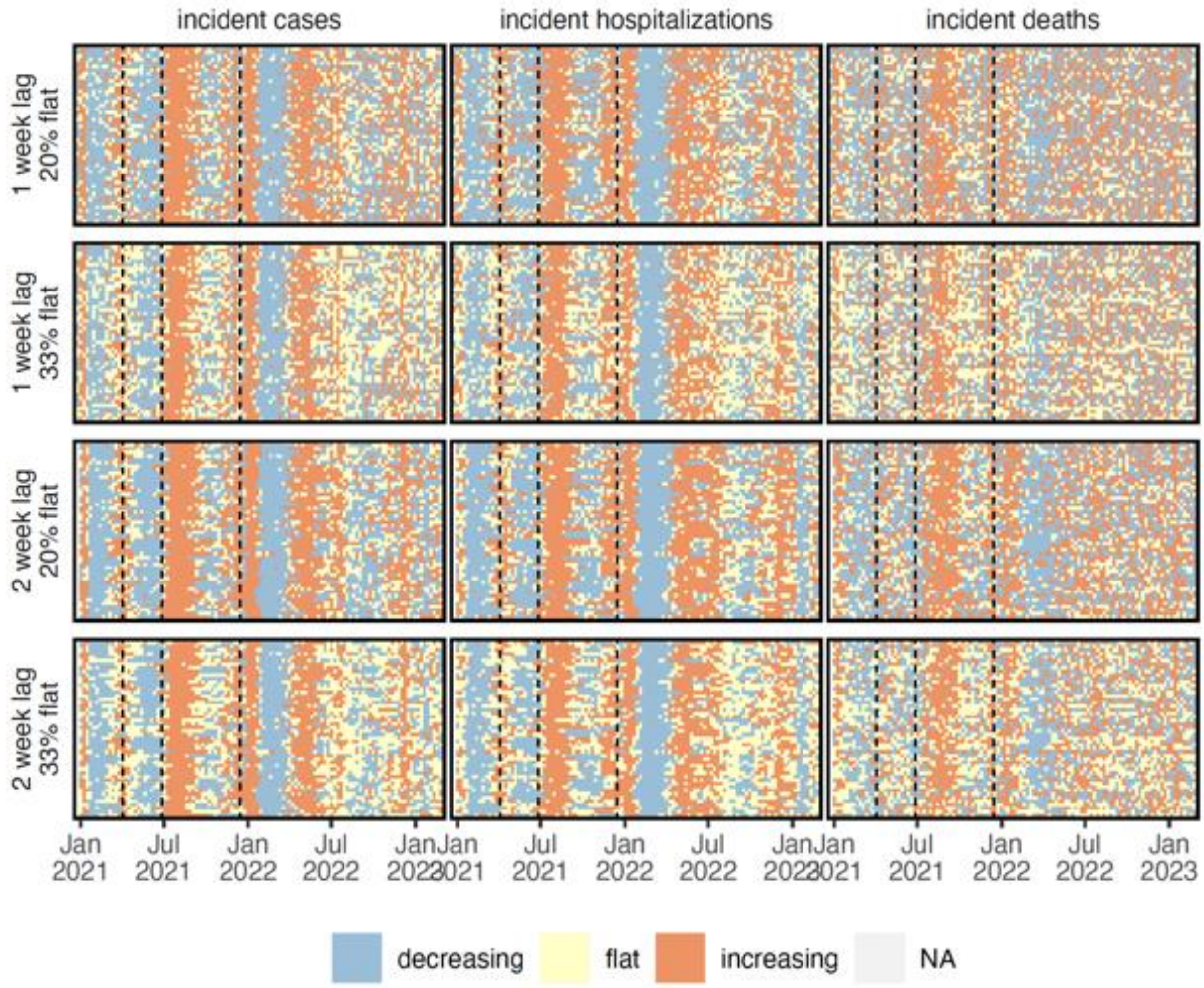


**Figure S34: Classification of observations under varying assumptions.** For all targets (panel columns) and different classification assumptions (panel rows), observations are classified as increasing (orange), flat (yellow), or decreasing (blue) or not available due to data anomalies (gray). Each set of assumptions varies the lag between change weeks, L (i.e., % change = $\log(o_t) - \log(o_{t-L})$) and the percent of observations classified as flat, C (i.e., lower/upper thresholds at quantiles $0.5 \pm \frac{C}{2}$. Within a single panel, each row represents a location (sorted by population size) and each column represents one week. In each panel, the horizontal dashed lines represent the emergence of Alpha, Delta, and Omicron variants respectively.

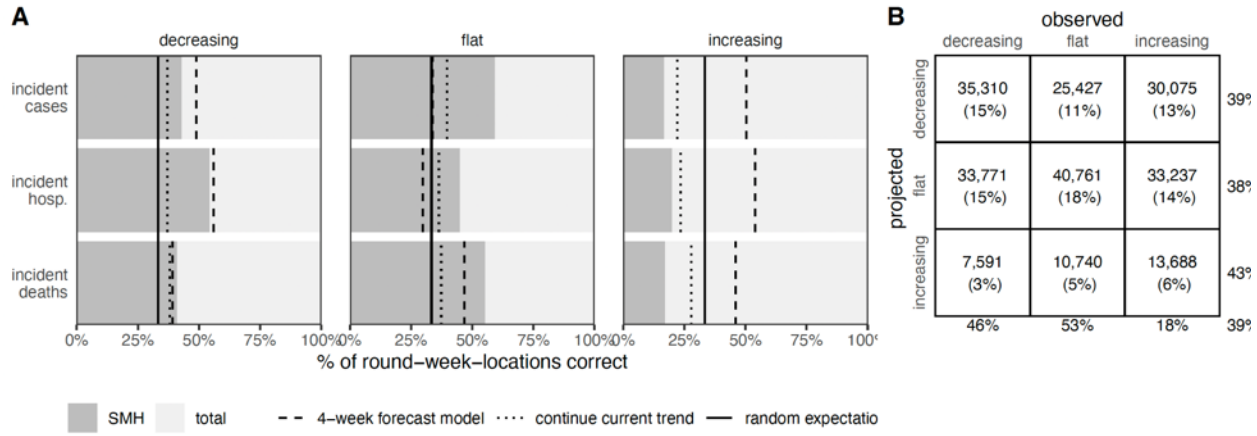


**Figure S35: Percent of projections for which trend of the SMH ensemble median is correct across all rounds, weeks, scenarios, and locations.** (A) For decreasing, flat and increasing observations, percent of observations (across all rounds, scenarios, weeks, locations) of incident cases, hospitalizations and deaths correctly identified by SMH ensemble projection median (gray), the 4-week forecast model (dashed line), a model that continues current trend (dotted), and the expectation if observations are classified randomly (solid). (B) The number (and percentage) of observations that are classified as decreasing, flat, or increasing by the SMH ensemble projection median, across case, hospitalization, and death projections. Totals are calculated across all rounds (meaning that some weeks are included multiple times, and therefore although 33% of observations are in each category, 33% of projections may not fall in each category). Percentages on the outside show the percent correct for a given projected (rows) or observed (columns) classification.

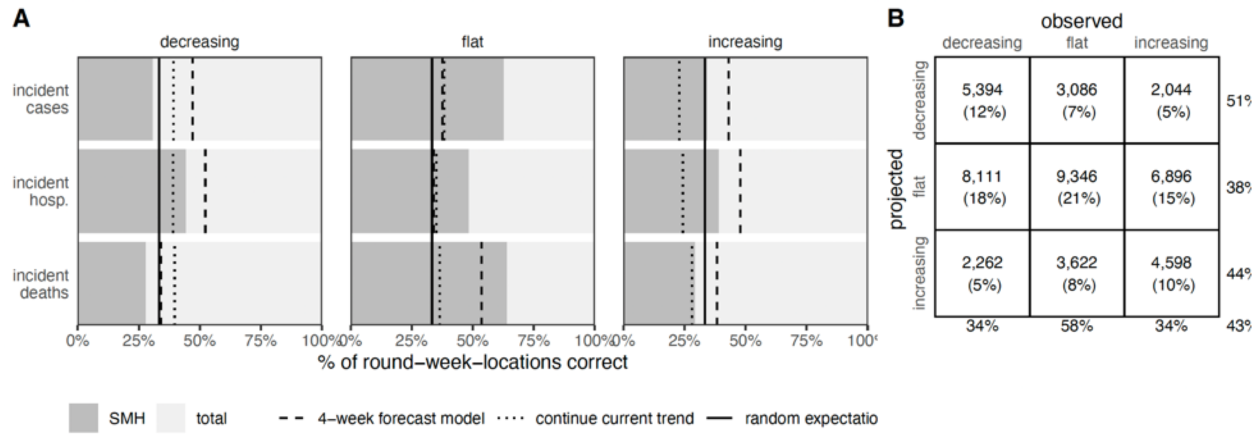


**Figure S36: Percent of projections for which trend of Q75 of the SMH ensemble is correct.** (A) For decreasing, flat and increasing observations, percent of observations (across all rounds and locations for plausible scenarios and weeks) of incident cases, hospitalizations and deaths correctly identified by SMH ensemble projection Q75 (gray), the 4-week forecast model (dashed line), a model that continues current trend (dotted), and the expectation if observations are classified randomly (solid). (B) For decreasing, flat and increasing observations in realistic scenarios and weeks, the number (and percentage) of observations that are classified as decreasing, flat, or increasing by the SMH ensemble projection Q75, across case, hospitalization, and death projections. Totals are calculated across all rounds (meaning that some weeks are included multiple times, and therefore although 33% of observations are in each category, 33% of projections may not fall in each category) and weighted by the plausibility of the scenario and week (for rounds with multiple plausible scenarios, this could introduce decimal totals; we rounded values down in these cases). Percentages on the outside show the percent correct for a given projected (rows) or observed (columns) classification.

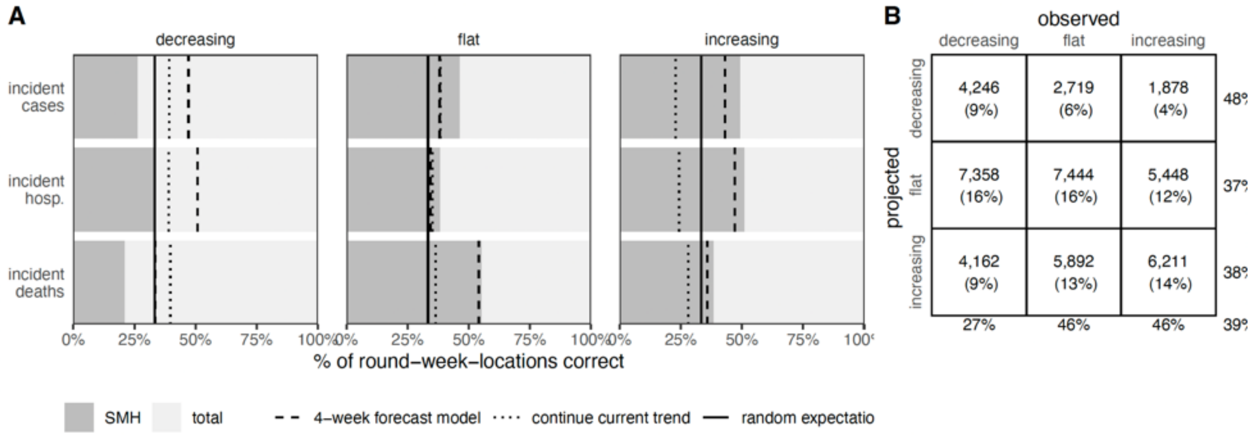


**Figure S37: Percent of projections for which trend of Q97.5 of the SMH ensemble is correct.** (A) For decreasing, flat and increasing observations, percent of observations (across all rounds and locations for plausible scenarios and weeks) of incident cases, hospitalizations and deaths correctly identified by SMH ensemble projection Q97.5 (gray), the highly informed model (dashed line), a model that continues current trend (dotted), and the expectation if observations are classified randomly (solid). (B) For decreasing, flat and increasing observations in realistic scenarios and weeks, the number (and percentage) of observations that are classified as decreasing, flat, or increasing by the SMH ensemble projection Q97.5, across case, hospitalization, and death projections. Totals are calculated across all rounds (meaning that some weeks are included multiple times, and therefore although 33% of observations are in each category, 33% of projections may not fall in each category) and weighted by the plausibility of the scenario and week (for rounds with multiple plausible scenarios, this could introduce decimal totals; we rounded values down in these cases). Percentages on the outside show the percent correct for a given projected (rows) or observed (columns) classification.

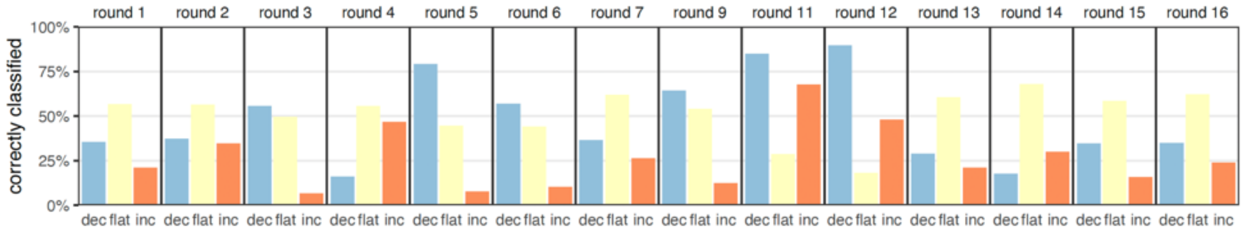


**Figure S38: Summary of correct classifications of SMH ensemble by round.** For observations that were decreasing (blue), flat (yellow), or increasing (orange), the percent of those observations that were correctly anticipated by the SMH ensemble median. Results are summarized across all locations and incident targets for a given round.

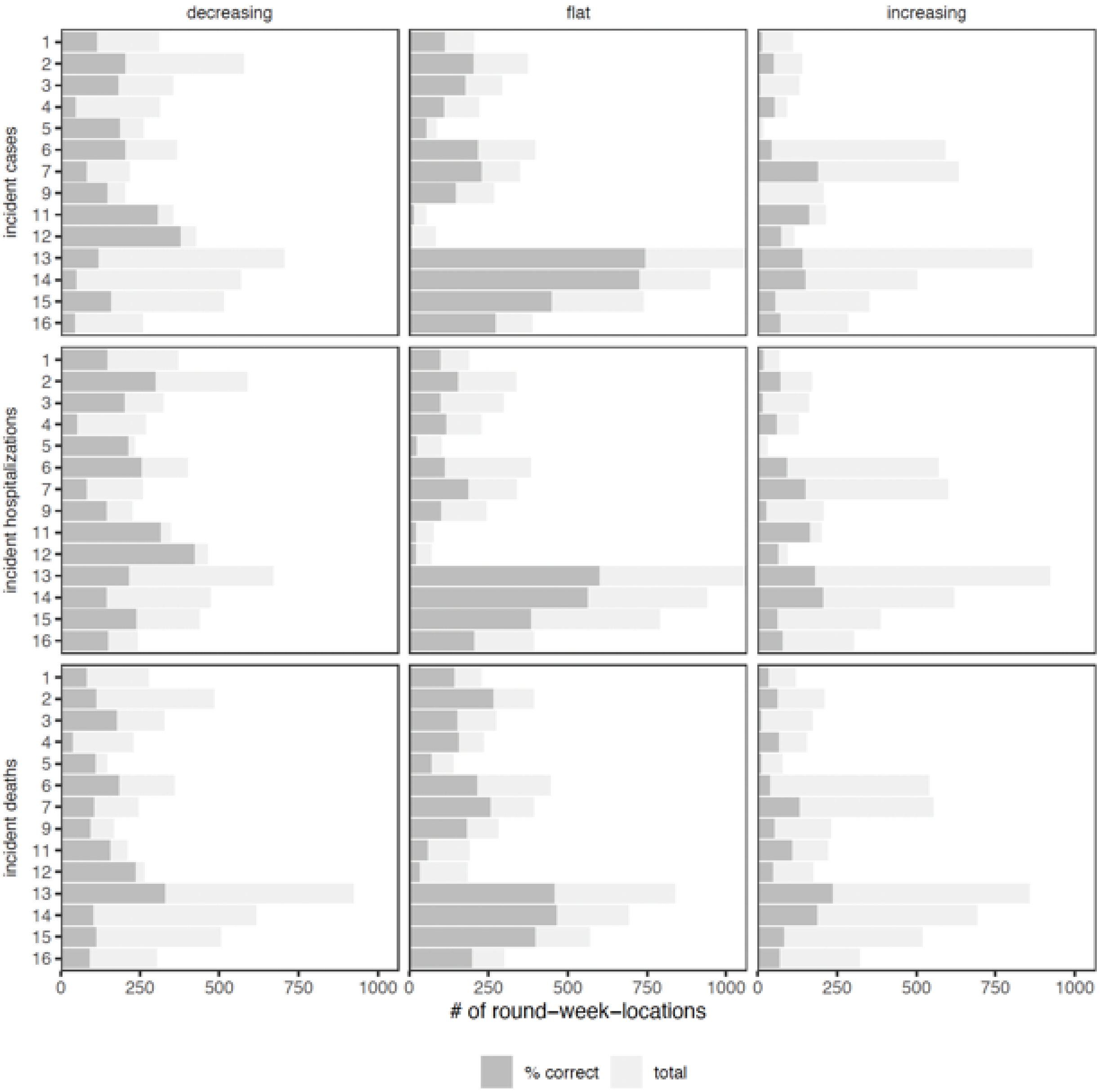


**Figure S39: Number of projections for which trend of SMH ensemble median is correct by round.** For decreasing, flat and increasing observations, number of observations (across all rounds, weeks, locations) of incident cases, hospitalizations and deaths observed (light gray) and correctly identified by SMH ensemble median (dark gray).

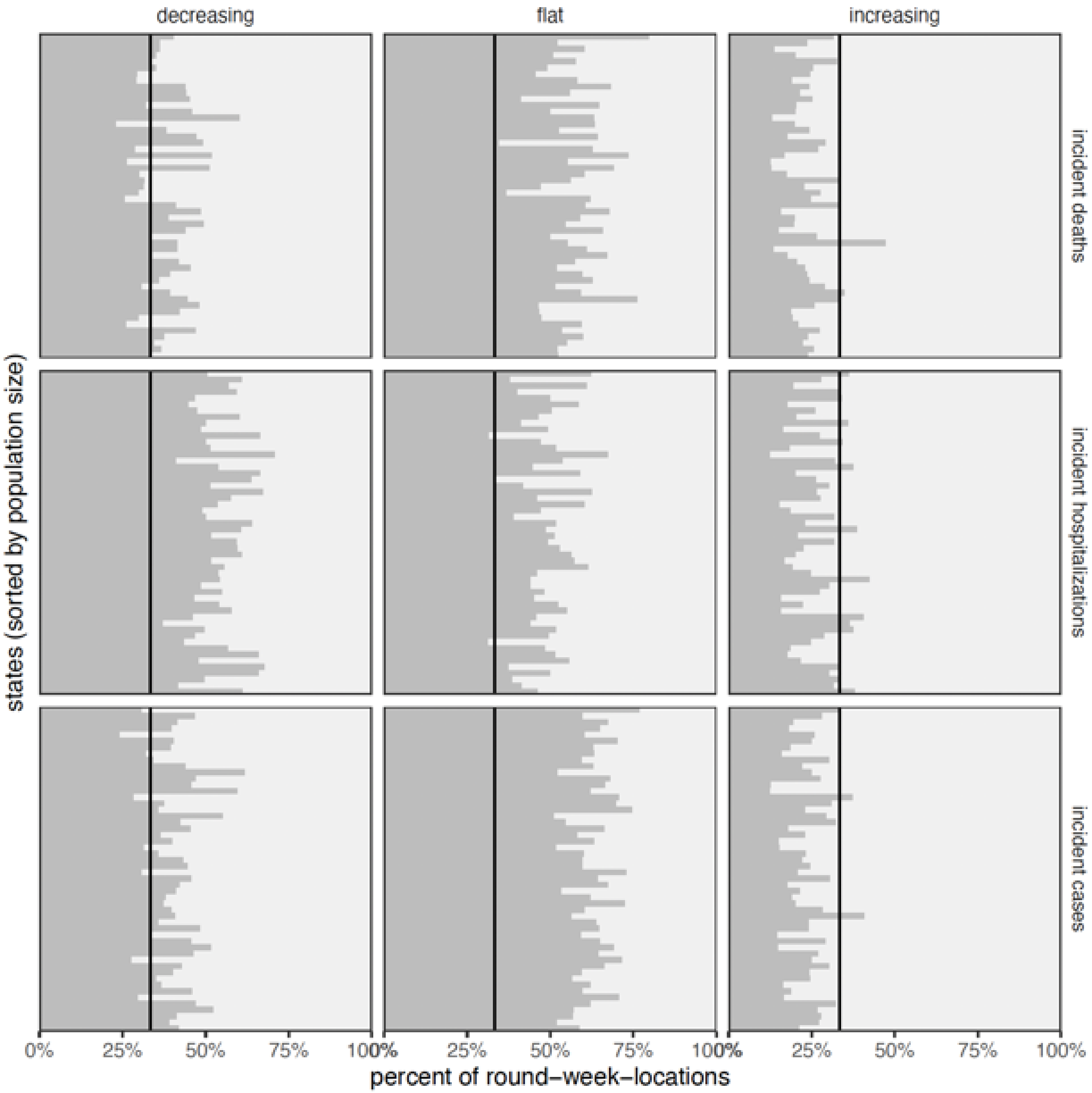


**Figure S40: Number of projections for which trend of SMH ensemble median is correct by location.** For decreasing, flat and increasing observations, number of observations (across all rounds, weeks, locations) of incident cases, hospitalizations and deaths observed (light gray) and correctly identified by SMH ensemble median (dark gray). In each panel, a single row represents one location (i.e., state), with locations with the largest population size at the top of the panel.

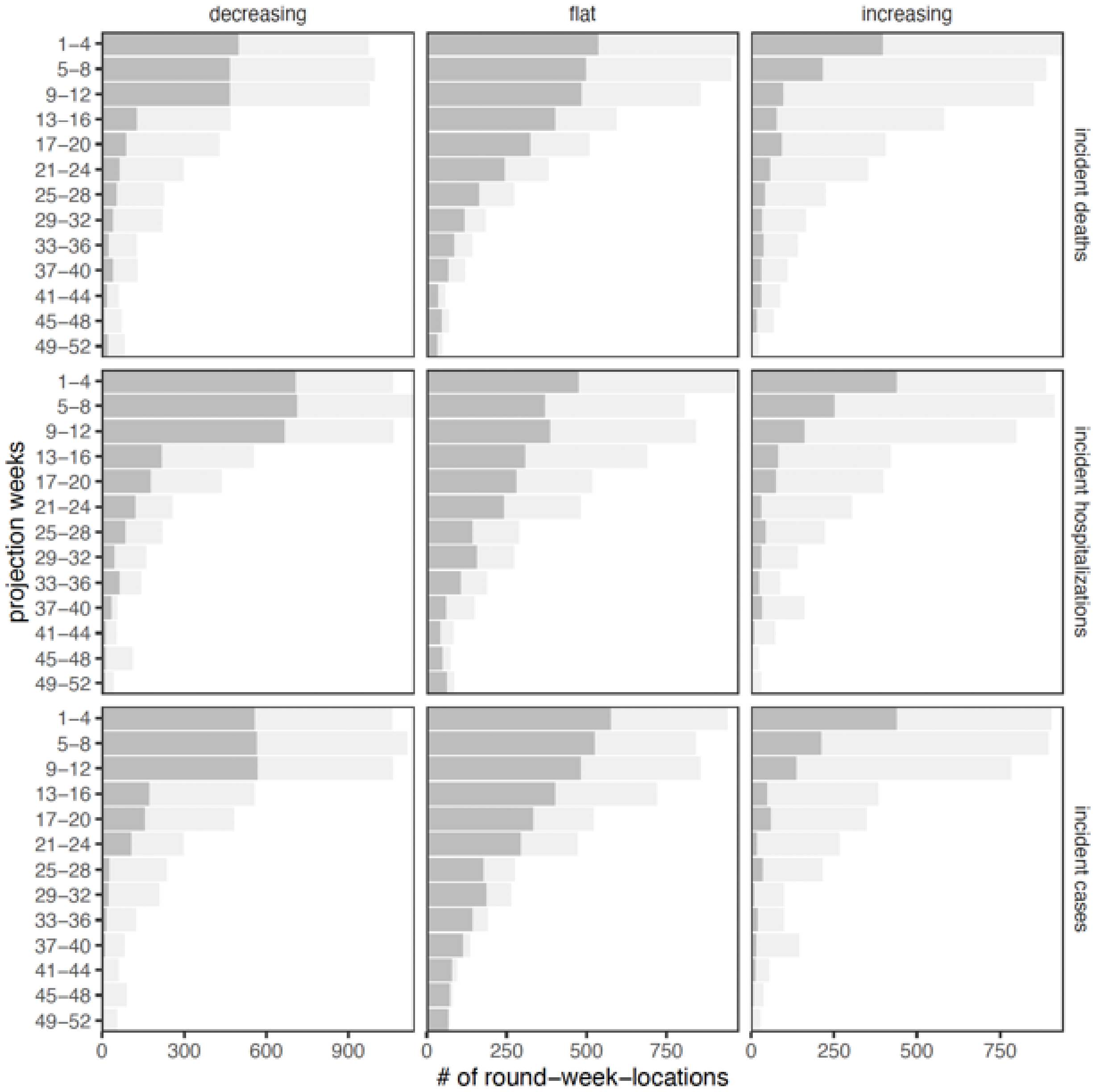


**Figure S41: Number of projections for which trend of SMH ensemble median is correct by projection horizon and target.** For decreasing, flat and increasing observations, number of observations (across all rounds, weeks, locations) of incident cases, hospitalizations and deaths observed (light gray) and correctly identified by SMH ensemble median (dark gray).

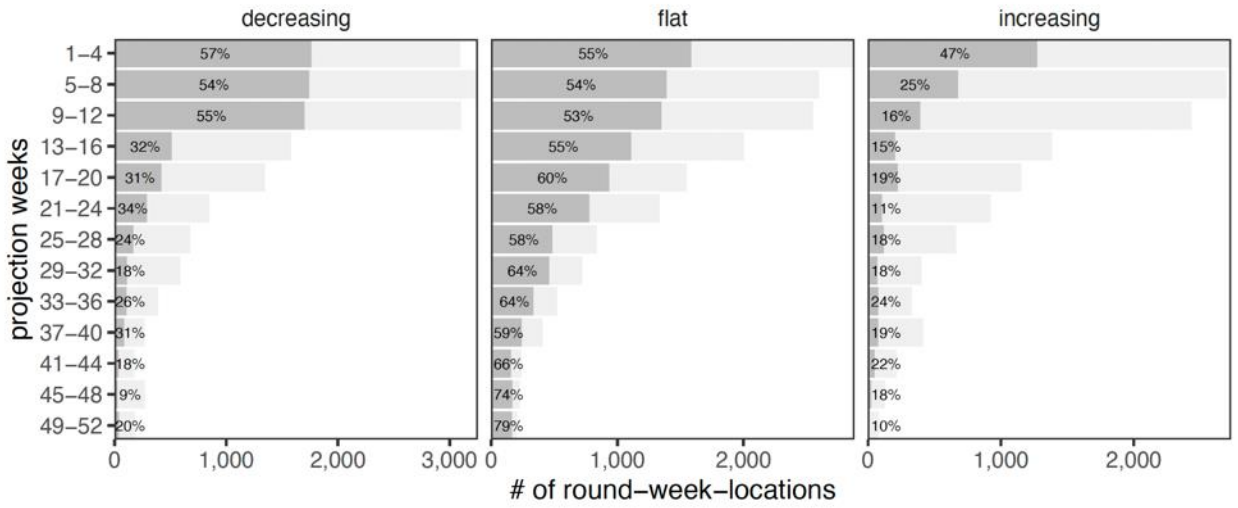


**Figure S42: Number of projections for which trend of SMH ensemble median is correct by projection horizon.** For decreasing, flat and increasing observations, number of observations (across all rounds, weeks, locations) of incident cases, hospitalizations and deaths observed (light gray) and correctly identified by SMH ensemble median (dark gray). Percent correctly identified is shown for each horizon.

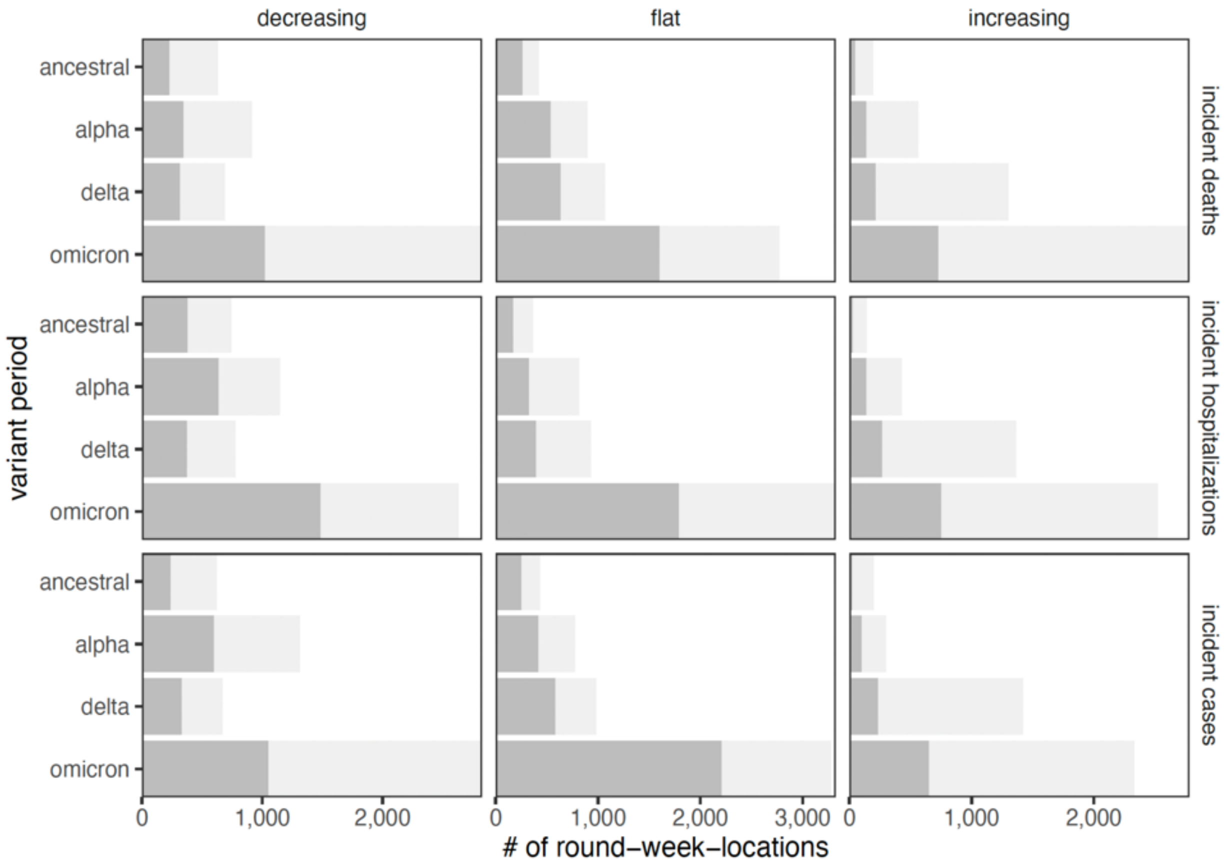

**Figure S43: Number of projections for which trend of SMH ensemble median is correct by variant period.** For decreasing, flat and increasing observations, number of observations (across all rounds, weeks, locations) of incident cases, hospitalizations and deaths observed (light gray) and correctly identified by SMH ensemble median (dark gray). Variant periods are defined on the same dates used for truncation (i.e., when variant reaches 50% prevalence nationally: ancestral, before 30 March 2021; alpha, between 30 March 2021 and 22 June 2021; delta, between 22 June 2021 and 20 December 2021, omicron after 20 December 2021.

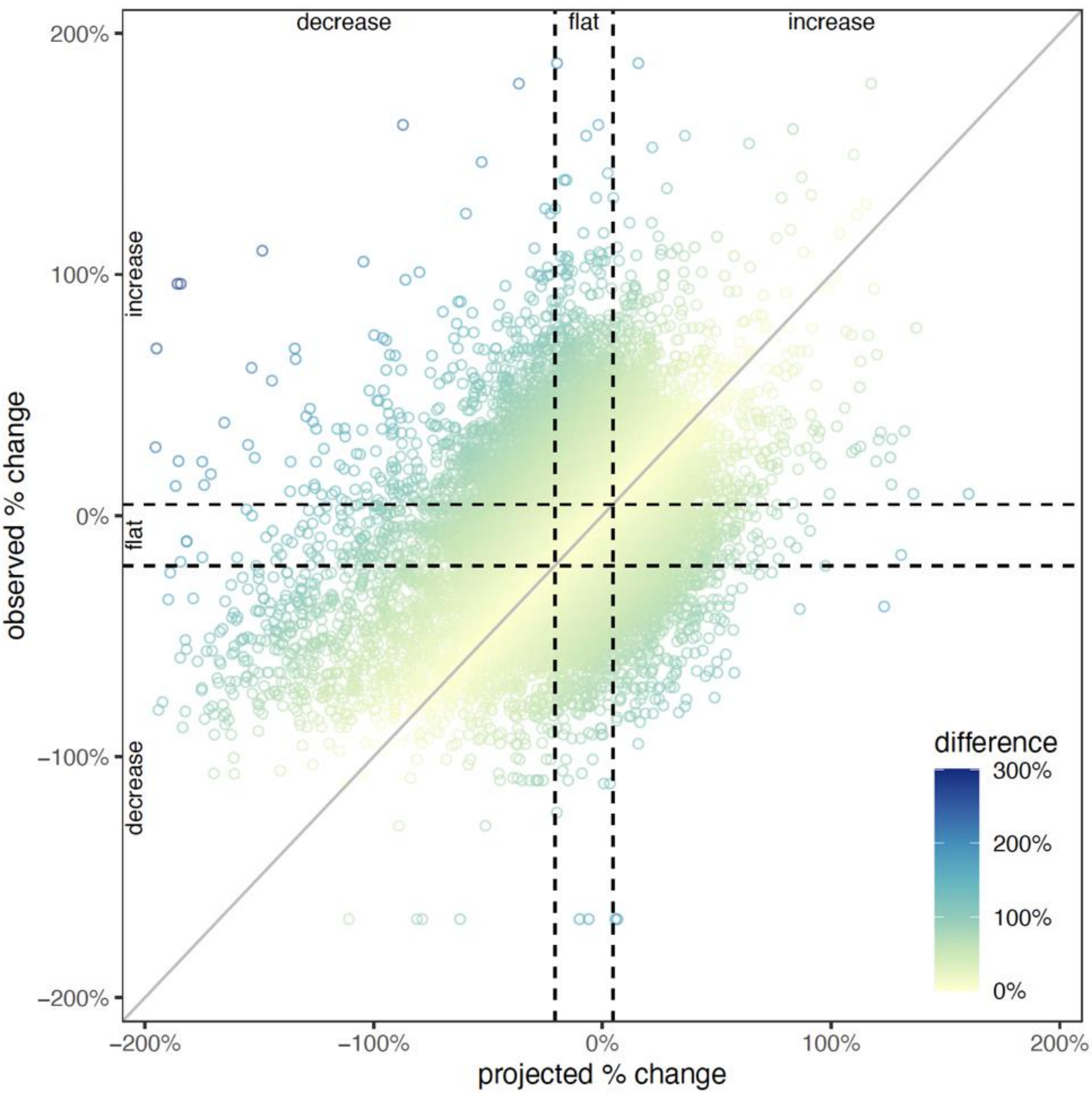


**Figure S44: Projected vs. observed percent (%) change.** The percent change in the SMH ensemble median is compared to the percent change in the observations (both calculated with a 2-week lag). Each point represents one projected round, week, location and incident target, and the color of the point represents the absolute difference between projected and observed percent change. Dashed lines indicate cutoff values for increasing and decreasing classifications that were used, and the gray line represents cases where the projected percent change is exactly what was observed.

### S5. Additional performance results

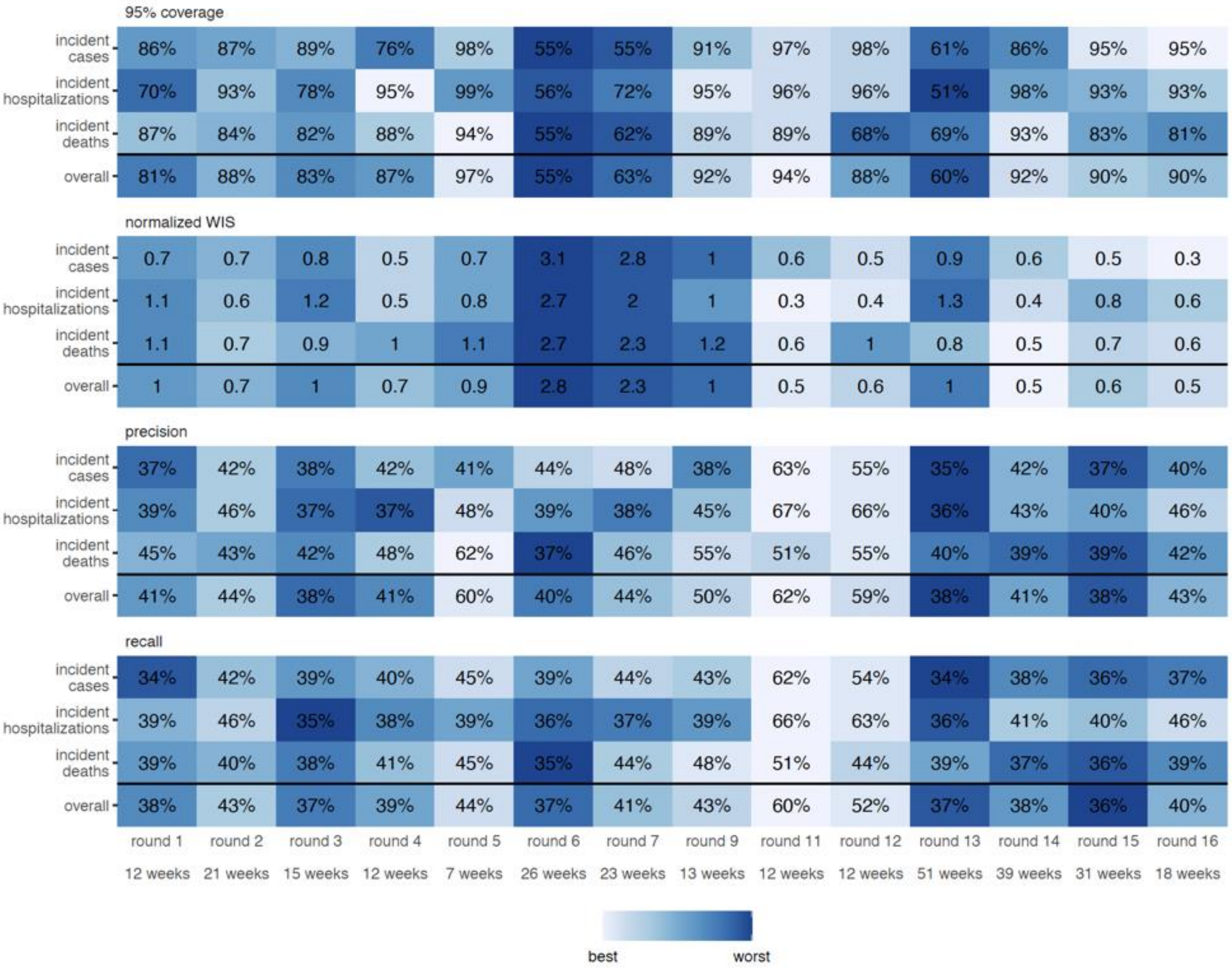


**Figure S45: 95% prediction interval coverage, normalized weighted interval score, precision, and recall by round and target.** Results are calculated for plausible scenario-weeks. Color scales show rank of each round by target and metric: ideal coverage ranked by absolute distance from 95% coverage, WIS ranked smallest to largest, precision and recall: red color scale shows, and blue color scale shows distance from 0. The number of plausible weeks evaluated is listed below each round (x-axis). For these results, precision and recall are averaged across all three classes (increasing, flat, and decreasing).

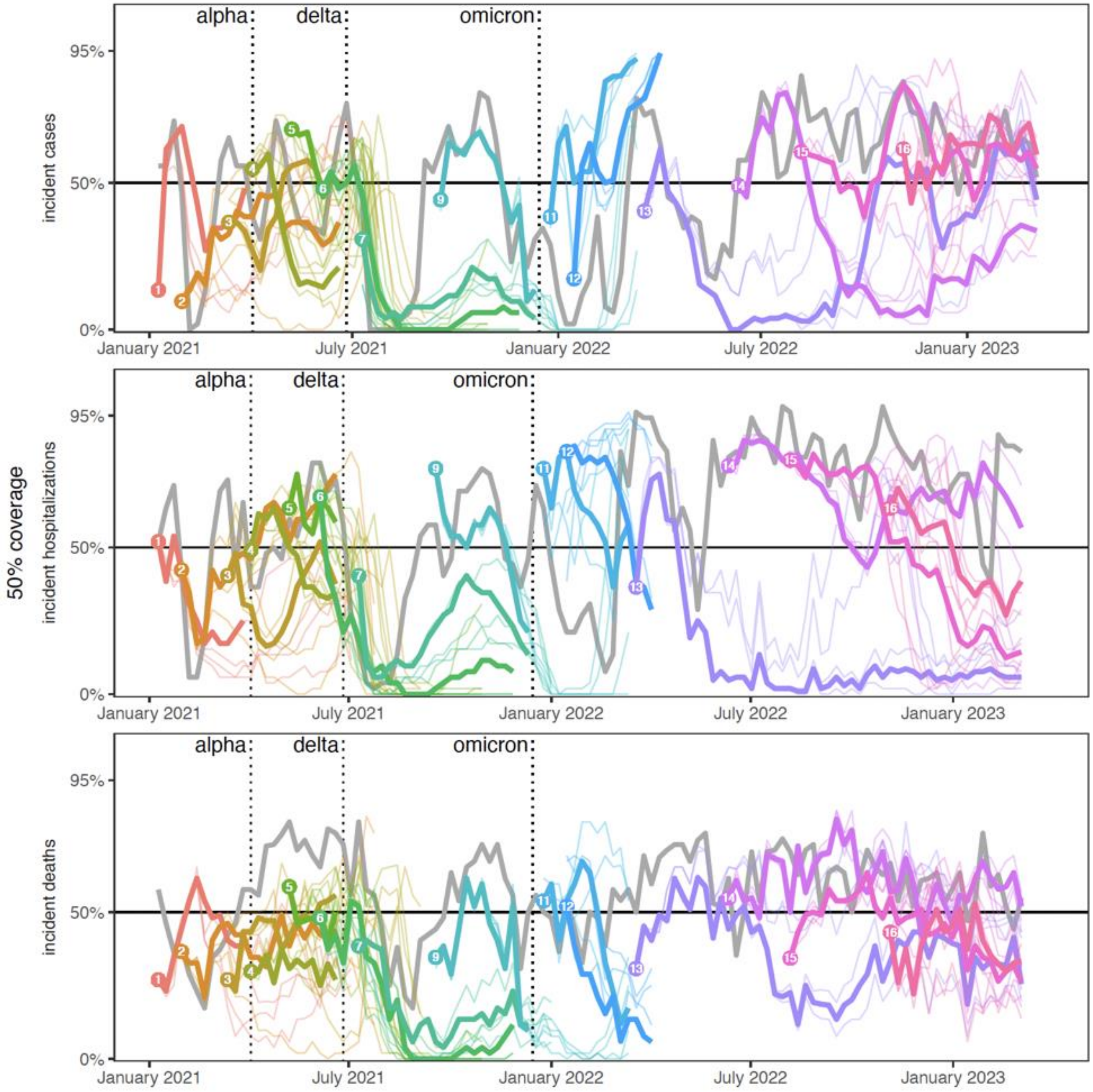


**Figure S46: Coverage of COVID-19 Scenario Modeling Hub (SMH) ensemble 50% projection interval for weekly incident cases, hospitalizations, and deaths.** Coverage is calculated across locations by round and scenario. Each round is represented by a different color and a number at the start of the round. Each scenario is represented by a different line, with realistic scenario-weeks bolded (see Methods). Performance of the 4-wk ahead COVID-19 Forecast Hub ensemble is shown in gray. Ideal coverage of 50% is shown with a horizontal black line, and the emergence dates for Alpha, Delta, and Omicron variants are shown with vertical dotted lines.

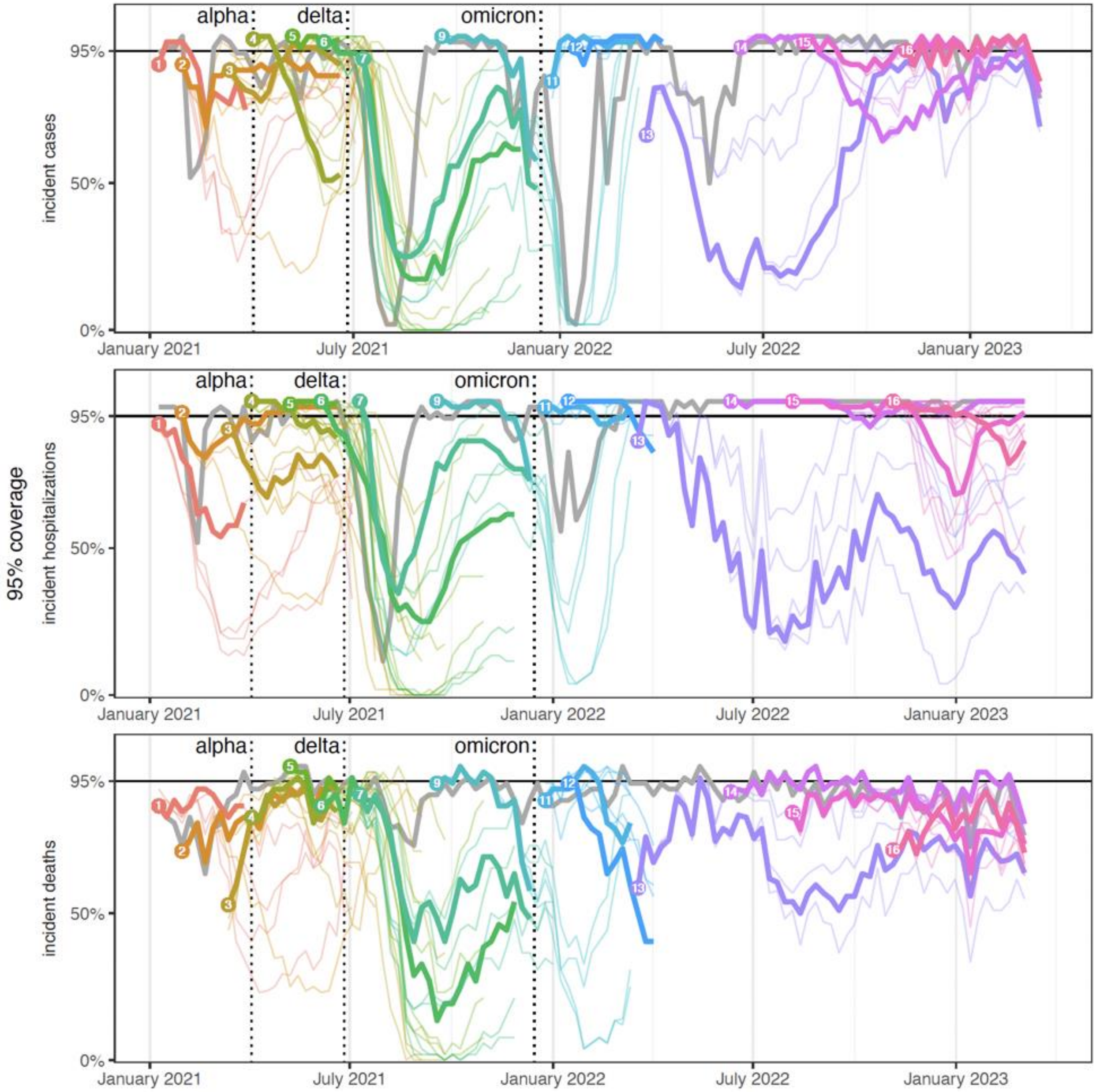


**Figure S47: Coverage of COVID-19 Scenario Modeling Hub (SMH) ensemble 95% projection interval for weekly incident cases, hospitalizations, and deaths.** Coverage is calculated across locations by round and scenario. Each round is represented by a different color and a number at the start of the round. Each scenario is represented by a different line, with realistic scenario-weeks bolded (see Methods). Performance of the 4-wk ahead COVID-19 Forecast Hub ensemble is shown in gray. Ideal coverage of 95% is shown with a horizontal black line, and the emergence dates for Alpha, Delta, and Omicron variants are shown with vertical dotted lines.

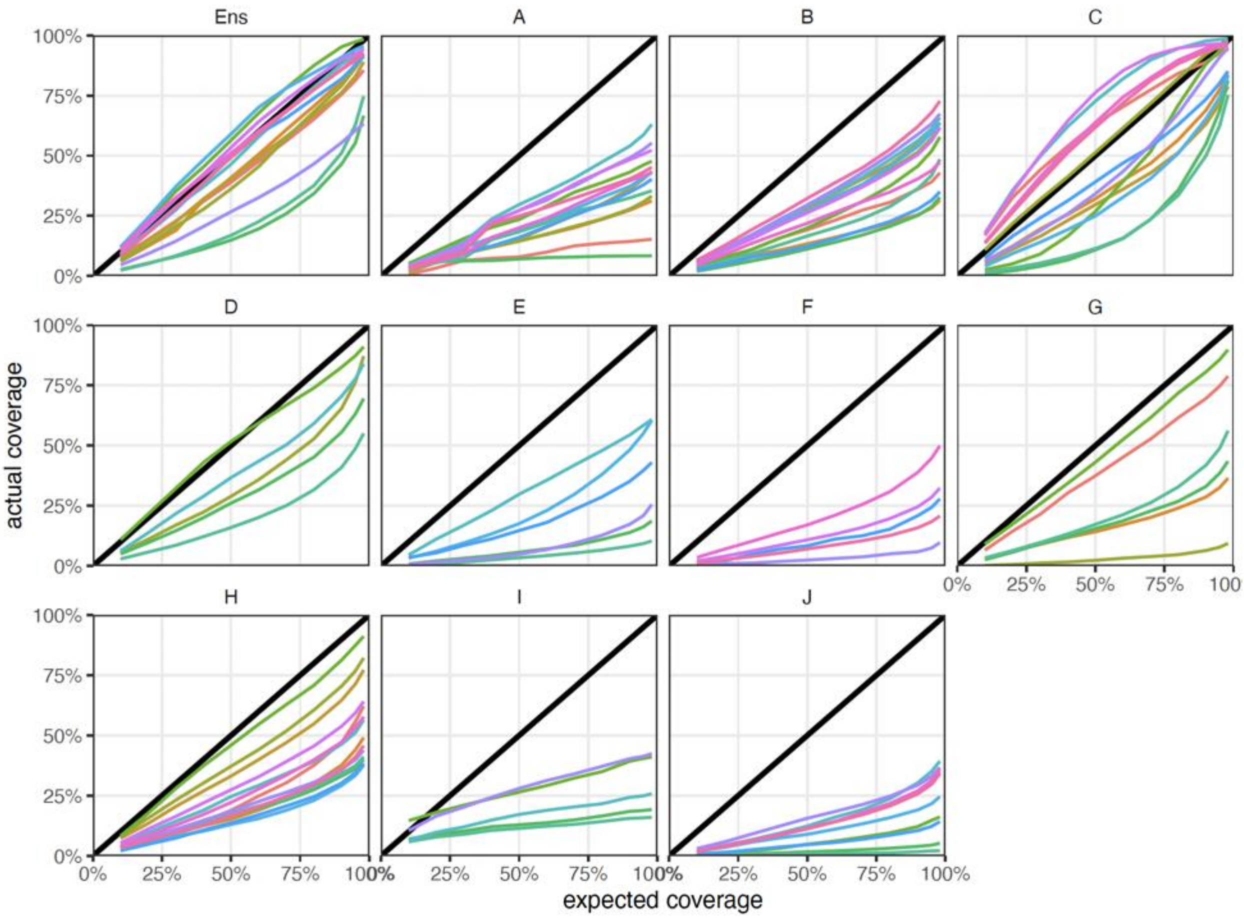


**Figure S48: Coverage results across quantiles, Q-Q plot.** For each SMH round (color), coverage for all available intervals (10%-90%, 95% and 98%) is compared to expected coverage for that interval. Results along the black indicate ideal coverage. Each panel shows results for a different model: the SMH ensemble and individual models in the rounds they contributed national projections.

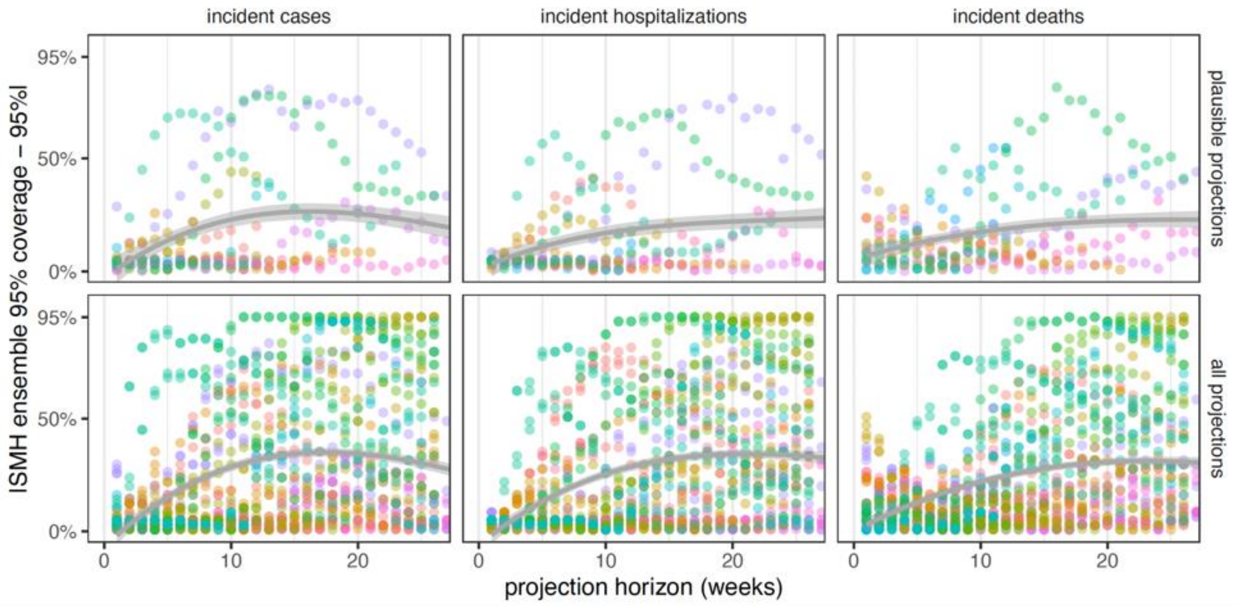


**Figure S49: Performance of 95% projection interval (PI) by projection horizon (in weeks) for incident cases, hospitalizations, and deaths.** Each point represents the difference of PI coverage from ideal (95%). For plausible projections (top), all plausible scenario-weeks are weighted equally, and results for all projections are also shown (bottom) with each scenario represented by a different point (color). Point color represents the SMH round. A simple generalized additive model is fit through the points in each panel. Coverage is only shown up to 26 weeks ahead, because there were not enough rounds with longer horizons to meaningfully compare.

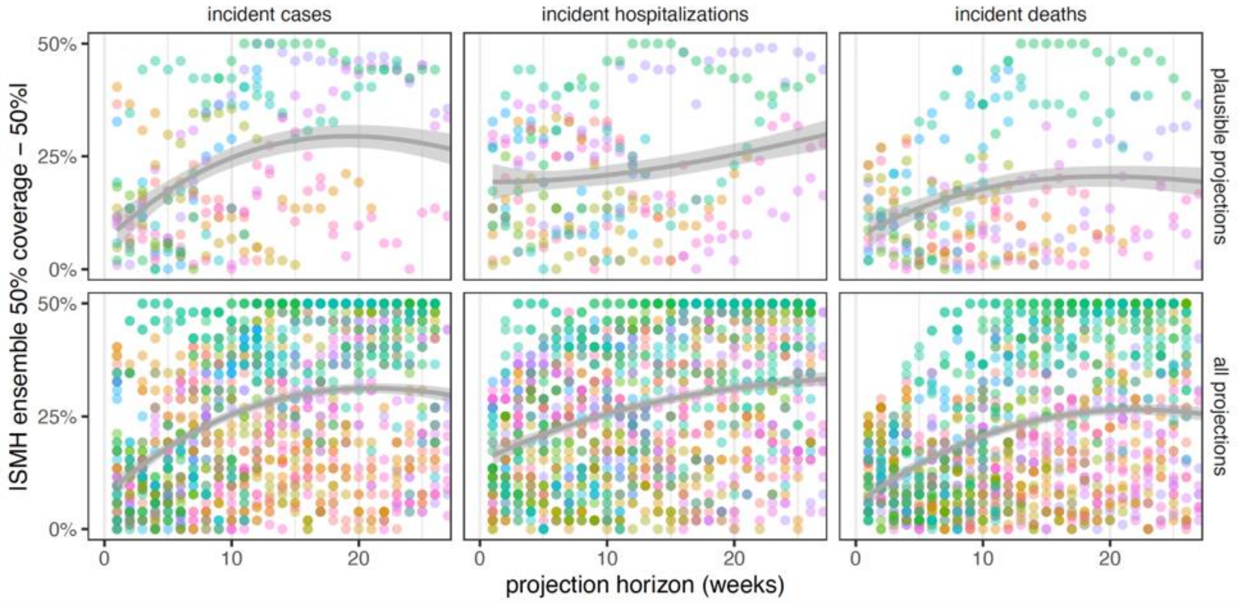


**Figure S50: Performance of 50% projection interval (PI) by projection horizon (in weeks) for incident cases, hospitalizations, and deaths.** Each point represents the difference of PI coverage from ideal (50%). For plausible projections (top), all plausible scenario-weeks are weighted equally, and results for all projections are also shown (bottom) with each scenario represented by a different point (color). Point color represents the SMH round. A simple generalized additive model is fit through the points in each panel. Coverage is only shown up to 26 weeks ahead, because there were not enough rounds with longer horizons to meaningfully compare.

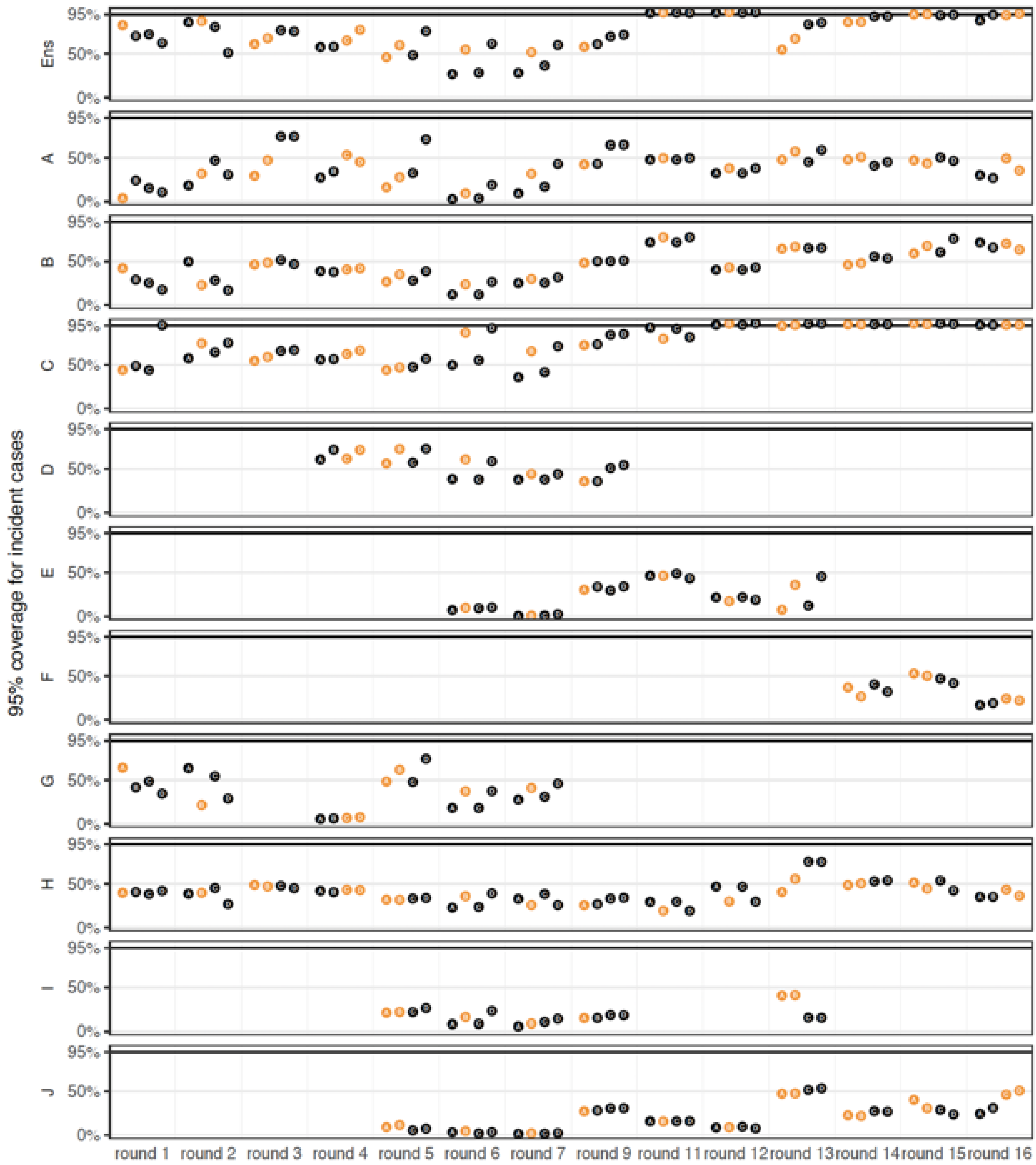


**Figure S51: Coverage of individual models across scenarios for incident cases.** Coverage is calculated across all weeks and locations in a single round for all scenarios (A-D), and most realistic scenarios are shown in orange. Each panel shows results for a single model, with "Ens" representing the SMH ensemble, and A-I representing individual models.

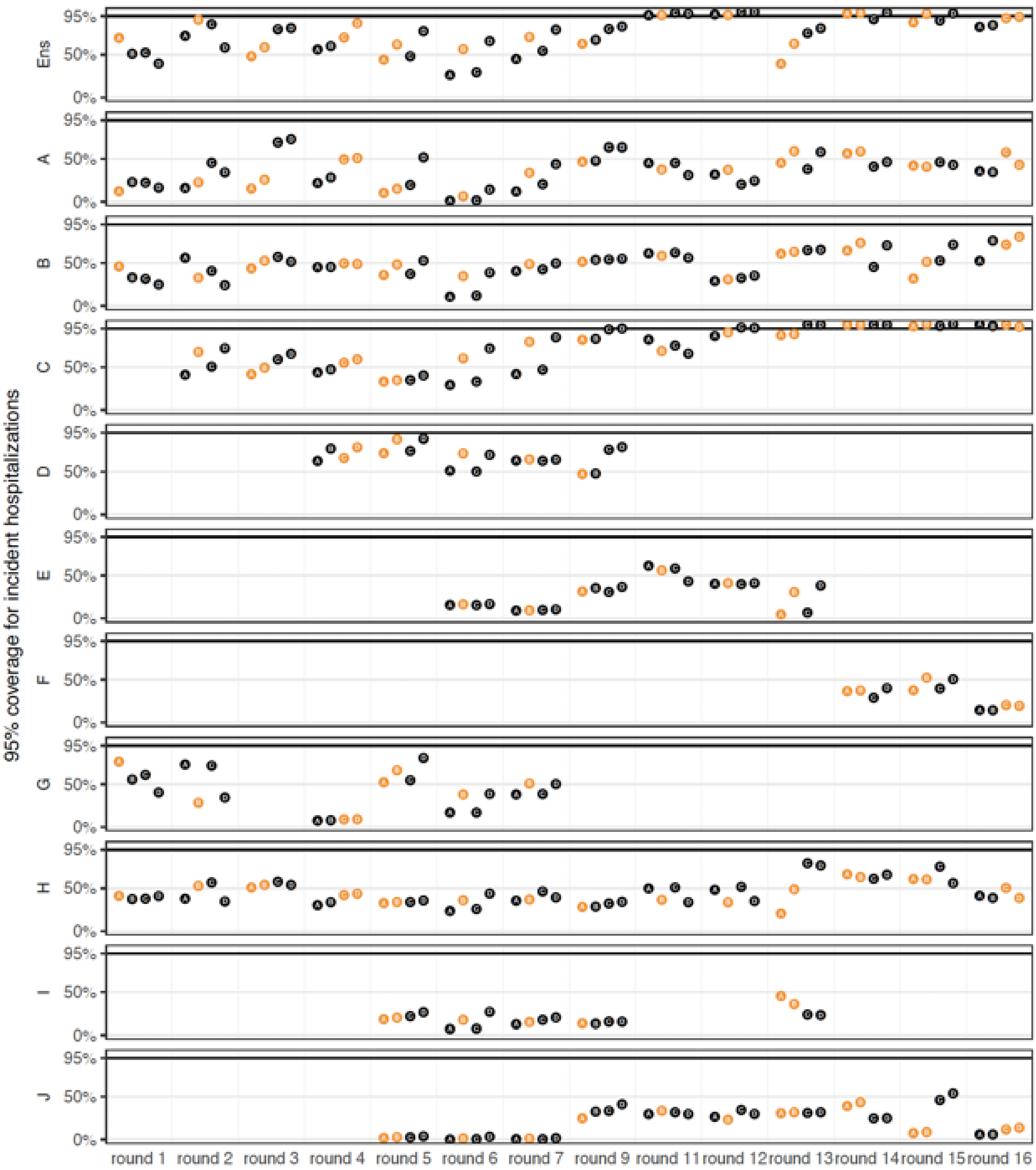


**Figure S52: Coverage of individual models across all scenarios for incident hospitalizations.** Coverage is calculated across all weeks and locations in a single round for all scenarios (A-D), and most realistic scenarios are shown in orange. Each panel shows results for a single model, with “Ens” representing the SMH ensemble, and A-I representing individual models.

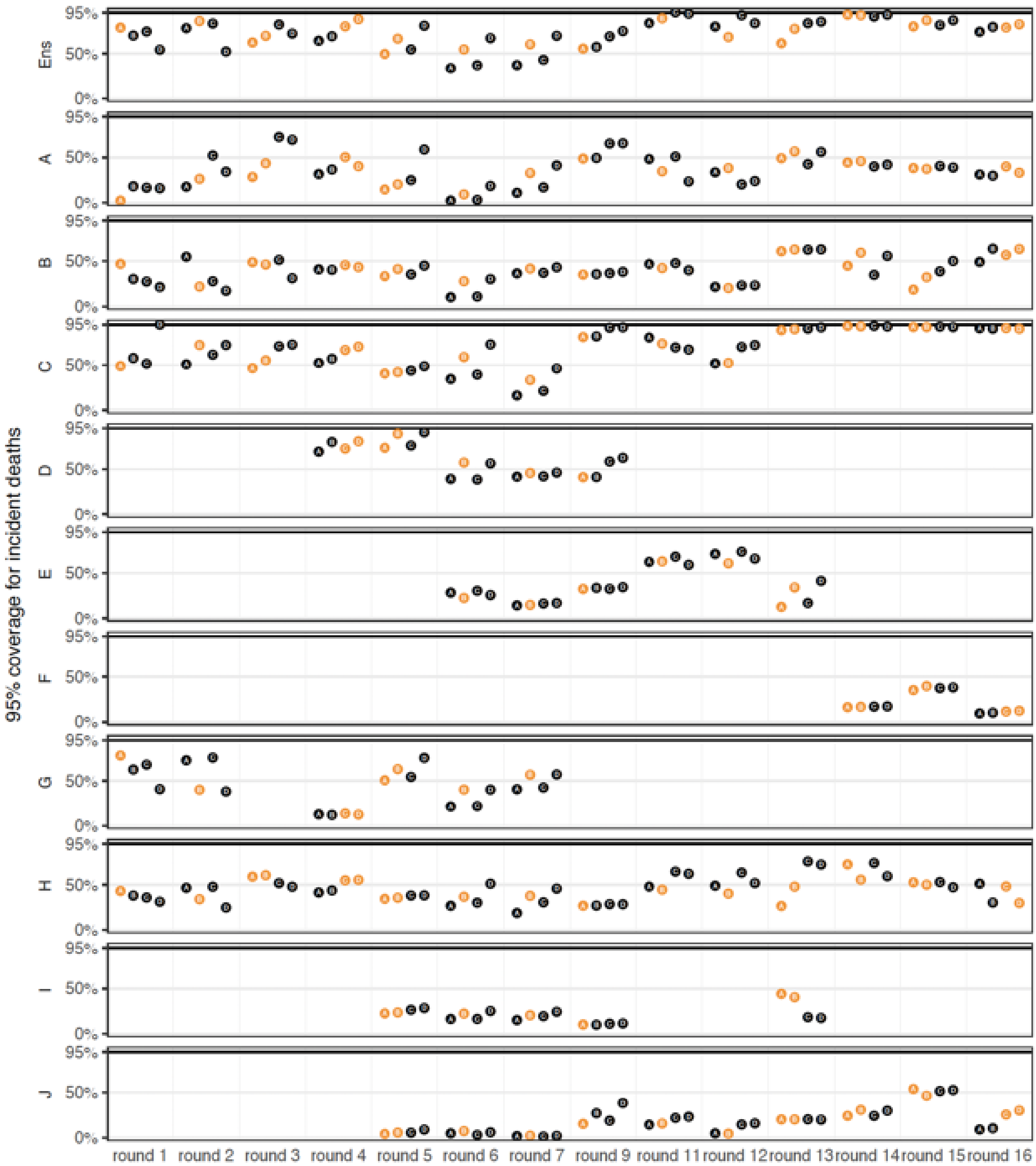


**Figure S53: Coverage of individual models across all scenarios for incident deaths.** Coverage is calculated across all weeks and locations in a single round for all scenarios (A-D), and most realistic scenarios are shown in orange. Each panel shows results for a single model, with “Ens” representing the SMH ensemble, and A-I representing individual models.

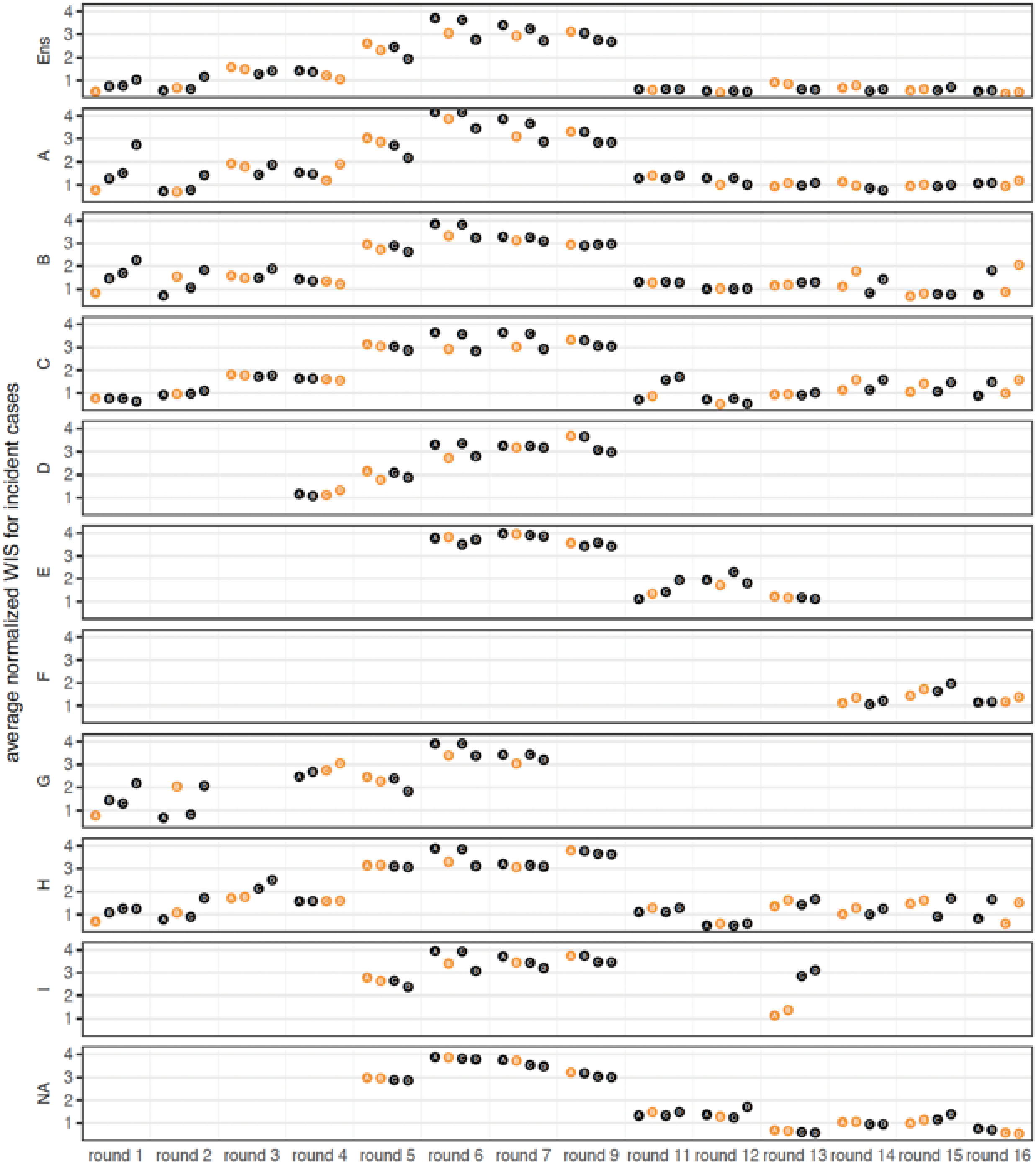


**Figure S54: Average normalized WIS of individual models across all scenarios for incident cases.** Normalized WIS (see Methods) is averaged across all weeks and locations in a single round for all scenarios (A-D), and most realistic scenarios are shown in orange. Each panel shows results for a single model, with “Ens” representing the SMH ensemble, and A-I representing individual models.

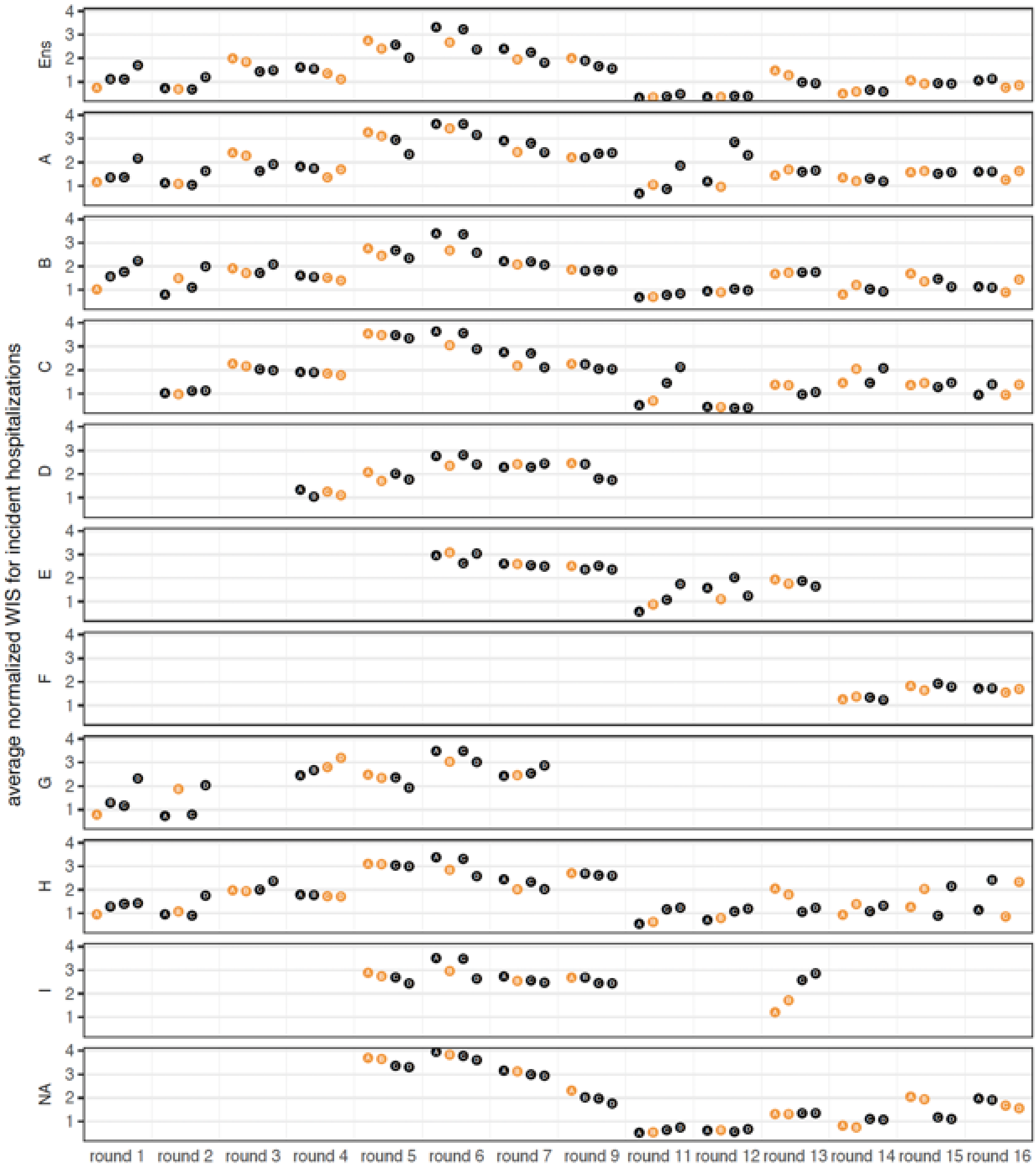


**Figure S55: Average normalized WIS of individual models across all scenarios for incident hospitalizations.** Normalized WIS (see Methods) is averaged across all weeks and locations in a single round for all scenarios (A-D), and most realistic scenarios are shown in orange. Each panel shows results for a single model, with "Ens" representing the SMH ensemble, and A-I representing individual models.

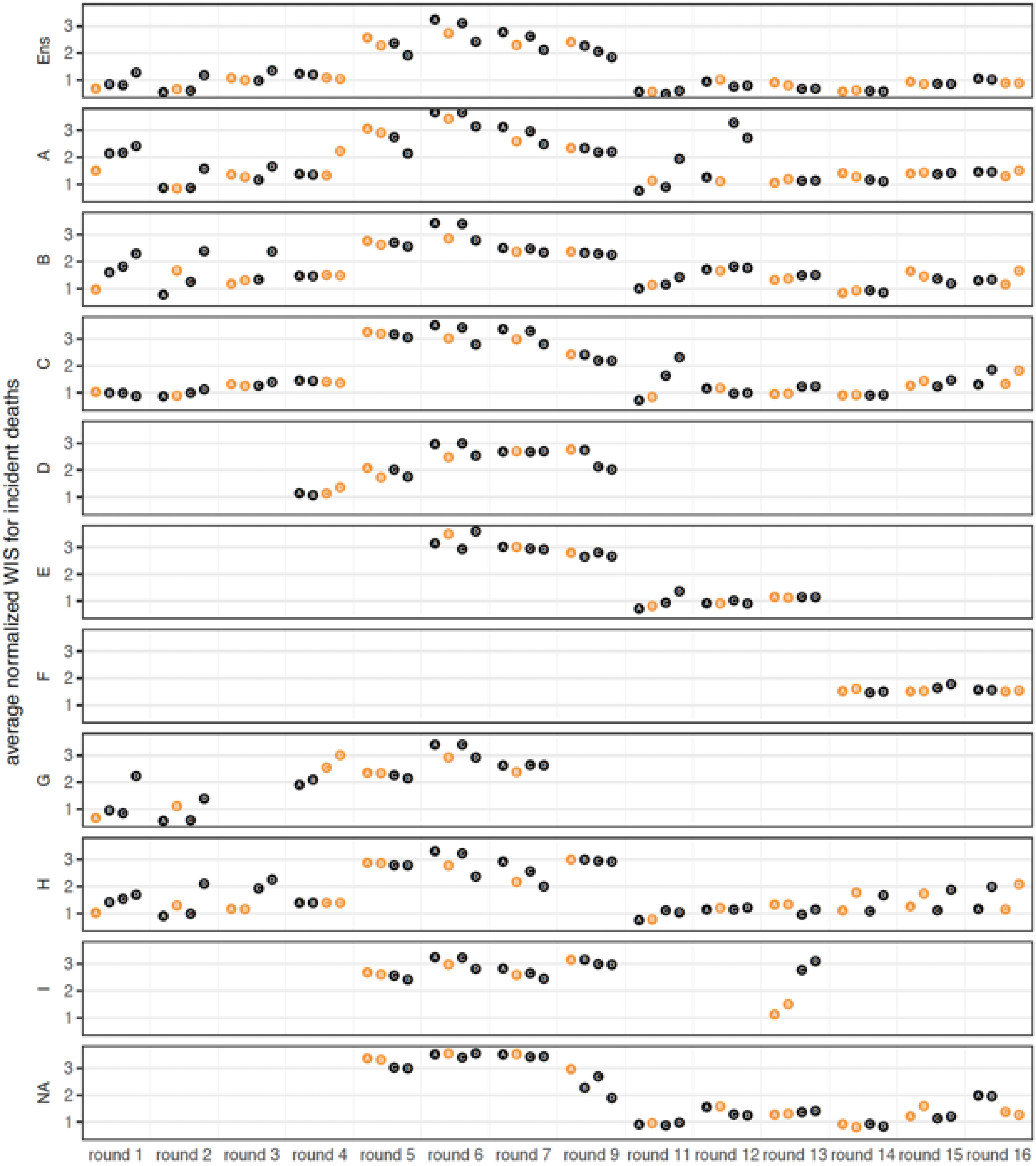


**Figure S56: Average normalized WIS of individual models across all scenarios for incident deaths.** Normalized WIS (see Methods) is averaged across all weeks and locations in a single round for all scenarios (A-D), and most realistic scenarios are shown in orange. Each panel shows results for a single model, with “Ens” representing the SMH ensemble, and A-I representing individual models.

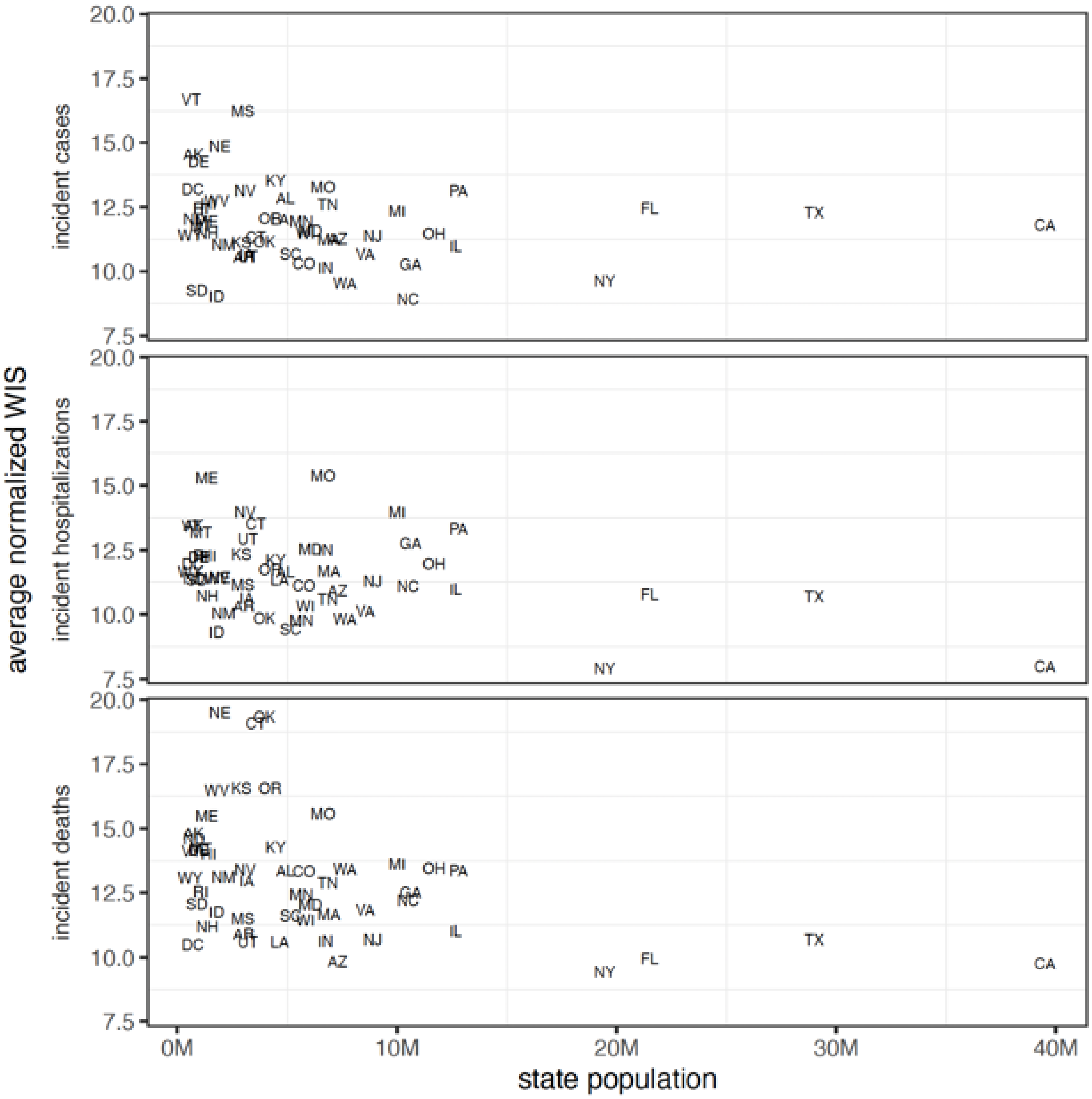


**Figure S57: Average normalized WIS by state population size for incident cases, hospitalizations, and deaths.** Normalized WIS is calculated by relativizing WIS by the standard deviation of WIS across all scenarios and models for a given week, location, target, and round. This yields a scale-free value, and we weighted average normalized WIS across all projection weeks for a given location (and scenario in the case of the SMH ensemble). Here we show results for plausible scenario-weeks.

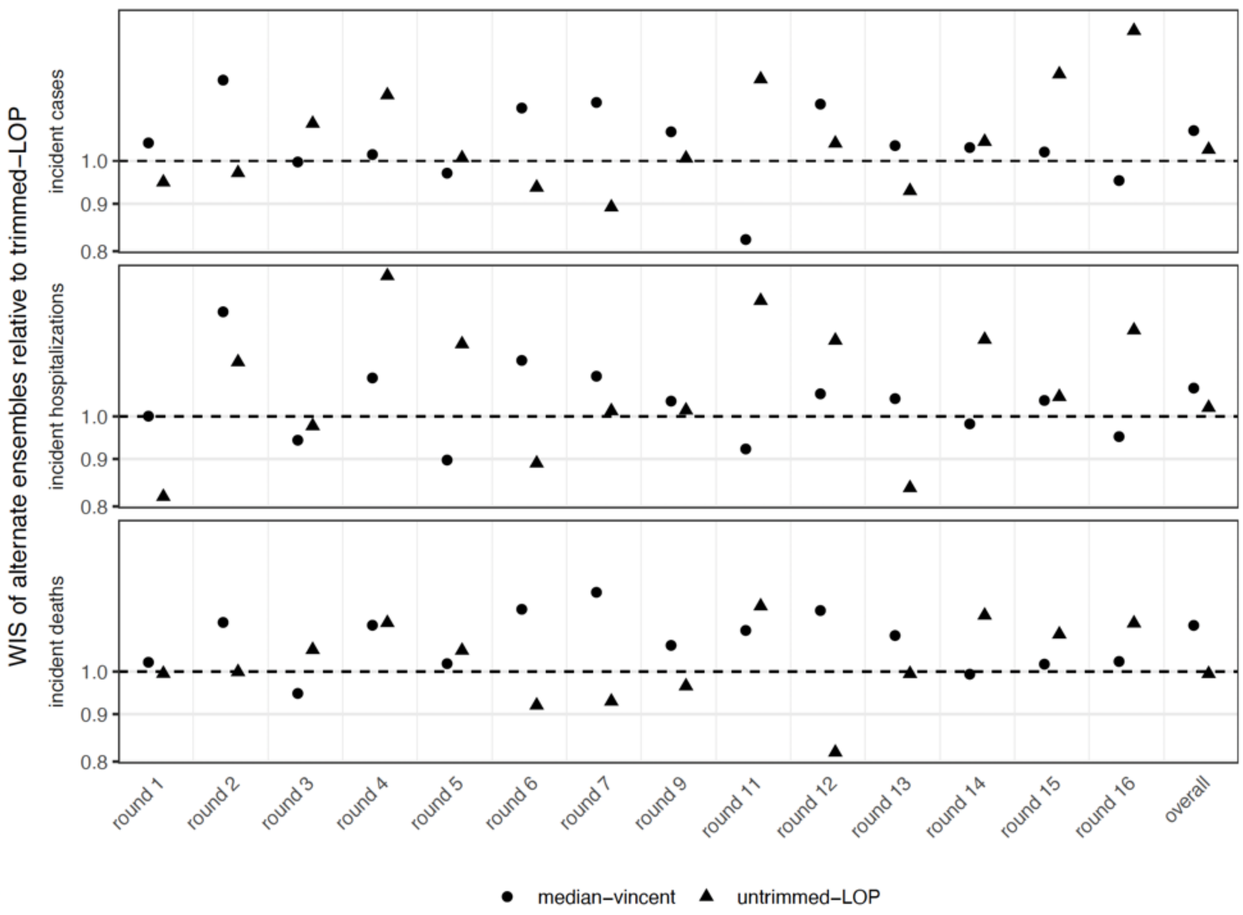


**Figure S58: Weighted interval score (WIS) ratio of alternate ensembles vs. trimmed-linear opinion pool (LOP) ensemble for U.S. COVID-19 Scenario Modeling Hub projections of incident cases, hospitalizations and deaths.** WIS for trimmed-LOP ensemble and each alternative is averaged across all locations and weeks in a given round (considering only realistic scenarios), or across all locations, weeks and rounds, for "overall". The WIS ratio is the average WIS for the alternative ensembles divided by the average WIS for the trimmed-LOP. Values below 1 (dashed horizontal line) indicate performance better than the trimmed-LOP. See Methods for details on each ensembling technique.

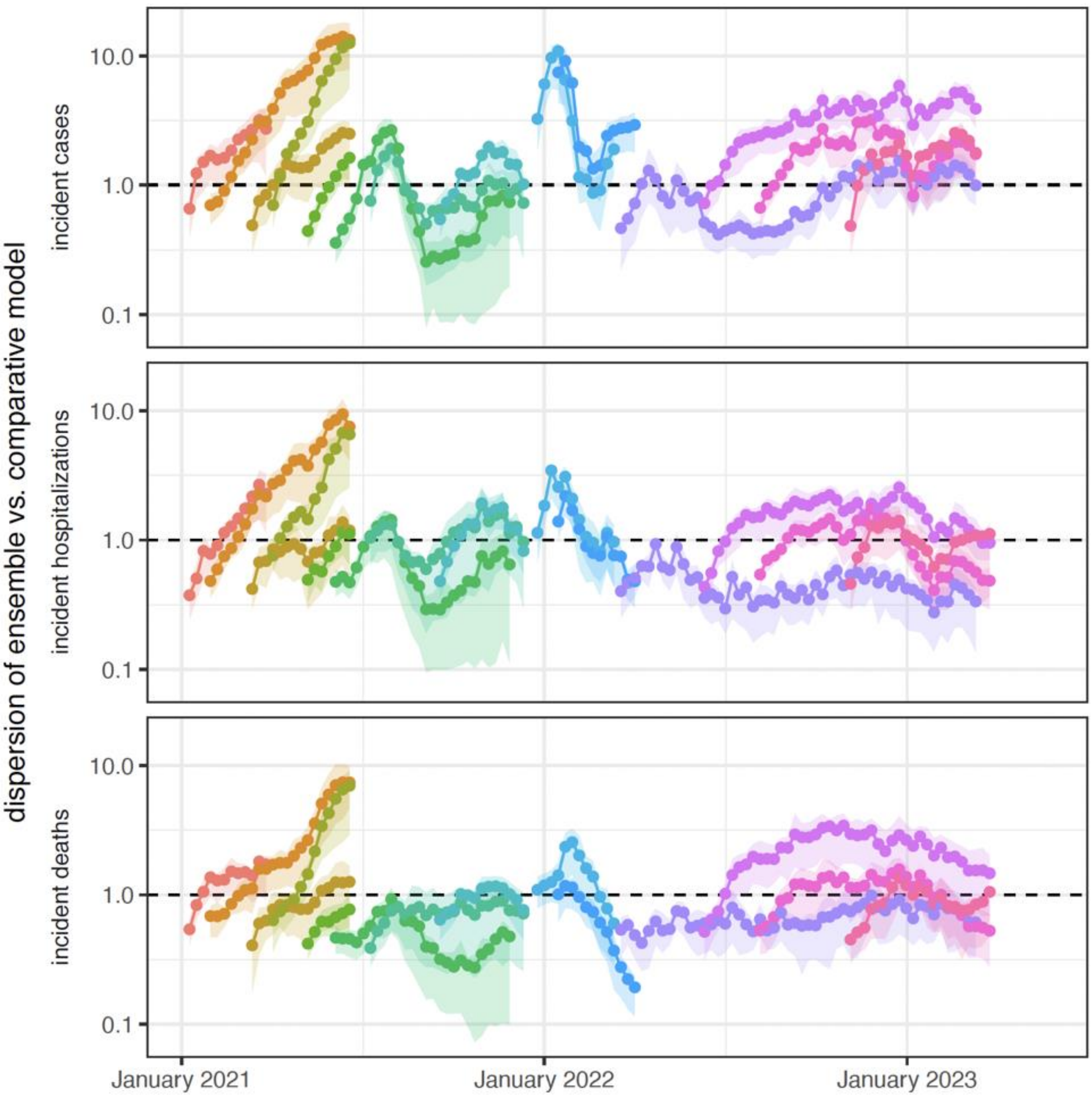


**Figure S59: SMH ensemble versus the 4-week forecast comparative model for incident cases, hospitalizations, and deaths.** For each projection week (indicated by a point) within a given round (round), we calculate dispersion (SMH ensemble)/dispersion(4-week forecast model) for all locations. We plot the mean (line) and IQR (Q25-Q75, ribbon) across locations. As such, values above 1 (dashed line) indicate the SMH ensemble has larger dispersion (i.e., wider projection intervals) than the 4-week forecast comparative model. Dispersion is measured by the dispersion component of the weighted interval score (first term of WIS formula $(u - l)$, see Methods).

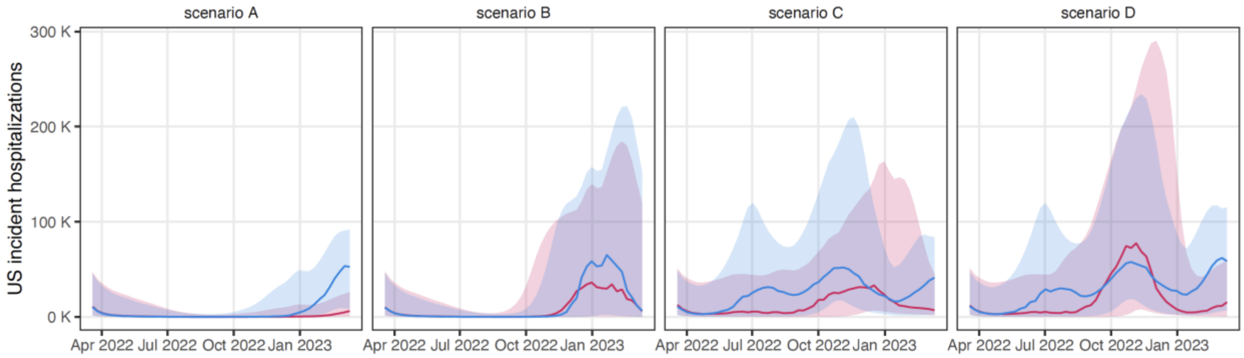


**Figure S60: Round 13 projections of US incident hospitalizations.** Projections are from the same model with two different assumptions about the specifics of how immunity wanes (waning times are assumed to be exponentially distributed for projections in pink, and assumed to be gamma distributed for projections in blue), despite both having the same scenario specified average duration and final protection levels. Line shows median projection, and ribbon shows 95% prediction interval.

Any use of trade, firm, or product names is for descriptive purposes only and does not imply endorsement by the U.S. Government.

10. K. Gangavarapu, A. A. Latif, J. Mullen, M. Alkuzweny, E. Hufbauer, G. Tsueng, E. Haag, M. Zeller, C. M. Aceves, K. Zaiets, M. Cano, J. Zhou, Z. Qian, R. Sattler, N. L. Matteson, J. I. Levy, R. T. Lee, L. Freitas, S. Maurer-Stroh, GISAID core and curation team, M. A. Suchard, C. Wu, A. I. Su, K. G. Andersen, L. D. Hughes, Center for Viral Systems Biology, "Alpha Variant Report," (available at https://outbreak.info/situation-reports/alpha?loc=GBR&loc=USA&loc=USA_US-CA&selected=GBR).

11. Z. Du, C. Liu, C. Wang, L. Xu, M. Xu, L. Wang, Y. Bai, X. Xu, E. H. Y. Lau, P. Wu, B. J. Cowling, Reproduction Numbers of Severe Acute Respiratory Syndrome Coronavirus 2 (SARS-CoV-2) Variants: A Systematic Review and Meta-analysis. *Clinical Infectious Diseases*. **75**, e293–e295 (2022).

12. R. Earnest, R. Uddin, N. Matluk, N. Renzette, S. E. Turbett, K. J. Siddle, C. Loreth, G. Adams, C. H. Tomkins-Tinch, M. E. Petrone, J. E. Rothman, M. I. Breban, R. T. Koch, K. Billig, J. R. Fauver, C. B. F. Vogels, K. Bilguvar, B. De Kumar, M. L. Landry, D. R. Peaper, K. Kelly, G. Omerza, H. Grieser, S. Meak, J. Martha, H. B. Dewey, S. Kales, D. Berenzy, K. Carpenter-Azevedo, E. King, R. C. Huard, V. Novitsky, M. Howison, J. Darpolor, A. Manne, R. Kantor, S. C. Smole, C. M. Brown, T. Fink, A. S. Lang, G. R. Gallagher, V. E. Pitzer, P. C. Sabeti, S. Gabriel, B. L. MacInnis, A. Altajar, A. DeJesus, A. Brito, A. E. Watkins, A. Muyombwe, B. S. Blumenstiel, C. Neal, C. C. Kalinich, C. Liu, C. Loreth, C. Castaldi, C. Pearson, C. Bernard, C. M. Nolet, D. Ferguson, E. Buzby, E. Laszlo, F. L. Reagan, G. Vicente, H. M. Rooke, H. Munger, H. Johnson, I. R. Tikhonova, I. M. Ott, J. Razeq, J. C. Meldrim, J. Brown, J. Wang, J. Vostok, J. P. Beauchamp, J. L. Grimsby, J. Hall, K. S. Messer, K. L. Larkin, K. Vernest, L. C. Madoff, L. M. Green, L. Webber, L. Gagne, M. A. Ulcena, M. C. Ray, M. E. Fisher, M. Barter, M. D. Lee, M. T. DeFelice, M. C. Cipicchio, N. L. Smith, N. J. Lennon, N. A. Fitzgerald, N. Kerantzas, P. Hui, R. Harrington, R. Downing, R. Haye, R. Lynch, S. E. Anderson, S. Hennigan, S. English, S. Cofsky, S. Clancy, S. Mane, S. Ash, S. Baez, S. Fleming, S. Murphy, S. Chaluvadi, T. Alpert, T. Rivard, W. Schulz, Z. M. Mandese, R. Tewhey, M. D. Adams, D. J. Park, J. E. Lemieux, N.

D. Grubaugh, Comparative transmissibility of SARS-CoV-2 variants Delta and Alpha in New England, USA. *Cell Reports Medicine*. **3**, 100583 (2022).
